# Systematic review on chronic non-communicable disease in disaster settings

**DOI:** 10.1101/2020.10.15.20213025

**Authors:** Christine Ngaruiya, Robyn Bernstein, Rebecca Leff, Lydia Wallace, Pooja Agrawal, Anand Selvam, Denise Hersey, Alison Hayward

## Abstract

**Background:** Non-communicable diseases (NCDs) constitute the leading cause of mortality globally. Low and middle-income countries (LMICs) not only experience the largest burden of humanitarian emergencies but are also disproportionately affected by NCDs, yet primary focus on the topic is lagging. We conducted a systematic review on the effect of humanitarian disasters on NCDs in LMICs assessing epidemiology, interventions, and treatment.

**Methods:** A systematic search in MEDLINE, MEDLINE (PubMed, for in-process and non-indexed citations), Social Science Citation Index, and Global Health (EBSCO) for indexed articles published before December 11, 2017 was conducted, and publications reporting on NCDs and humanitarian emergencies in LMICs were included. We extracted and synthesized results using a thematic analysis approach and present the results by disease type. The study is registered at PROSPERO (CRD42018088769).

**Results:** Of the 85 included publications, most reported on observational research studies and almost half (48.9%) reported on studies in the Eastern Mediterranean Region (EMRO), with scant studies reporting on the African and Americas regions. NCDs represented a significant burden for populations affected by humanitarian crises in our findings, despite a dearth of data from particular regions and disease categories. The majority of studies included in our review presented epidemiologic evidence for the burden of disease, while few studies addressed clinical management or intervention delivery. Commonly cited barriers to healthcare access in all phases of disaster and major disease diagnoses studied included: low levels of education, financial difficulties, displacement, illiteracy, lack of access to medications, affordability of treatment and monitoring devices, and centralized healthcare infrastructure for NCDs. Screening and prevention for NCDs in disaster-prone settings was supported. Refugee status was independently identified both as a risk factor for diagnosis with an NCD and conferred worse morbidity.

**Conclusions:** An increased focus on the effects of, and mitigating factors for, NCDs occurring in disaster-afflicted LMICs is needed. While the majority of studies included in our review presented epidemiologic evidence for the burden of disease, research is needed to address contributing factors, interventions, and means of managing disease during humanitarian emergencies in LMICs.

## Background

Non-communicable diseases (NCDs) constitute the leading cause of mortality globally, accounting for 70% of deaths worldwide (1). This percentage is projected to rise in the next fifteen years, with the steepest increase in morbidity and mortality from NCDs projected to occur in Low and Middle-Income Countries (LMICs). The World Health Organization (WHO) projects a 10% rise in mortality in Africa from NCDs in from 2015 to 2030 (2).

The rise in NCDs in LMICs coincides with an increasing burden of humanitarian disasters (3). The International Red Cross defines a disaster as: “a sudden, calamitous event that seriously disrupts the functioning of a community or society and causes human, material, and economic or environmental losses that exceed the community’s or society’s ability to cope using its own resources” (4). The United Nations Office for Disaster Risk Reduction (UNISDR) recorded over 1.35 million people killed by natural hazards between 1997-2017, with disproportionate mortality in LMICs (5). Poverty, rapid urbanization, inadequate infrastructure, and underdeveloped disaster warning and health systems are all contributors to morbidity and mortality in disasters (5, 6). The fatalities in a disaster are most directly related to the preexisting vulnerability of the population it affects (1, 6).

Humanitarian response in emergencies can be divided into four phases: mitigation, preparedness, response, and recovery. Mitigation refers to measures designed to either prevent or reduce the impact of disasters. Preparedness refers to preparation and instruction to strengthen the overall capacity and capability of a country or a community for events that cannot be mitigated. The response phase encompasses the immediate aftermath of a disaster when disaster response plans are implemented. Finally, the recovery phases is comprised of restoration efforts which must occur in parallel with routine operations and activities (7). However, each phase may be prolonged, exemplified by current conflicts in Israel-Palestine, Syria, Yemen, the Democratic Republic of Congo (DRC), and countries of the Lake Chad region which are party to protracted conflicts where the response or active phase has persisted (8). Such conflicts may be international, defined as arising between two or more states, or non-international, in which one or more non-State armed groups are involved, as defined under Article 3 common to the 1949 Geneva Conventions, and may blur the line between relief and development which further necessitates attention to NCDs amongst humanitarian actors (9, 10).

The scale of humanitarian disasters has increased in recent decades for two primary reasons. Firstly, the frequency and ferocity of climate-related disasters are increasing due to climate changes (11). Secondly, the number of refugees, displaced persons, and migrants are at an all-time high due to the unprecedented refugee crises in Syria, Iraq, and the Democratic Republic of Congo (12). According to the UNHCR Global Trends Report, an unprecedented 79.5 million people are estimated to have been displaced from their homes as internally displaced persons (IDPs) or refugees in 2019 - the largest figure ever recorded (13). Disasters may directly exacerbate NCDs through effects such as increased stress levels (14), exposures such as inhalation of substances that trigger worsening of pulmonary disease (15), and exacerbation of underlying disease secondary to limited access to care (16).

Despite the growing burden of humanitarian crises with increasing populations at risk for morbidity and mortality from NCDs, primary focus on the topic is lagging. It is essential to better understand the effect of disasters on NCDs in LMICs as the mortality and morbidity are projected only to increase given climate change and population growth in vulnerable areas (17). In this context, we conducted a systematic review on the effect of humanitarian disasters on NCDs in LMICs assessing epidemiology, interventions, and treatment. To our knowledge, this is the first systematic review of its kind cross-cutting both regions and disease type. Our aims are to guide allocation of resources, future research, and policy development.

## Methods

An experienced medical librarian performed a comprehensive search of multiple databases after consultation with the lead authors and a Medical Subject Heading (MeSH) analysis of key articles provided by the research team. In each database, we used an iterative process to translate and refine the searches. English, Arabic and French language articles were eligible and no date restrictions were applied. The formal search strategies used relevant controlled vocabulary terms and synonymous free text words and phrases to capture the concepts of noncommunicable, chronic and noninfectious diseases, and different types of humanitarian emergencies including natural disasters, armed conflicts, terrorism, and failed states (see Appendix). The databases searched were MEDLINE (OvidSP 1946-August Week 2 2015), MEDLINE (PubMed, for in-process and non-indexed citations), Social Science Citation Index, and Global Health (EBSCO). We included studies conducted in LMICs investigating noncommunicable diseases in the context of humanitarian emergencies. Studies conducted in high income countries (HICs) and review articles were excluded. No other restrictions on study type were applied. The original searches were run August 10, 2015 and were rerun on December 11, 2017. The full strategy for PubMed is available in the appendix. The study is registered at PROSPERO (CRD42018088769).

Retrieved references were pooled in EndNote and de-duplicated to 4,430 citations. Two separate screeners independently evaluated the titles, abstracts and full text of the eligible articles, with vetting by a third reviewer. The flowchart per PRISMA is presented in **Figure 1**.

## Results

We retrieved a total of 4,430 references. 4,342 studies were excluded by title or abstract, and 158 articles were read in full. Out of the studies screened, 85 were included in the final thematic analysis. We present the results by disease type. For disease type, we have five categories, which consist of the lead four NCDs in order of burden (18): cardiovascular disease (CVD), cancer, chronic respiratory disease, diabetes, and a section on other NCDs (defined as those not fitting into the lead four categories).

### Cardiovascular disease

Cardiovascular disease was the most highly studied NCD after diabetes, and 29 studies addressed this (see Table 1). Syrian refugees were the most commonly studied population among studies addressing CVD (19-22). Prevalence of disease was high, as demonstrated by Sibai et al. in a community-based cross-sectional study of residents of Beirut, Lebanon with circulatory diseases accounting for nearly 60% of diagnoses, and ischemic heart disease was the leading diagnosis (20). They also demonstrated that strokes had the second highest case fatality rate (54%), which was second only to sepsis (60%).

**Table 1:**
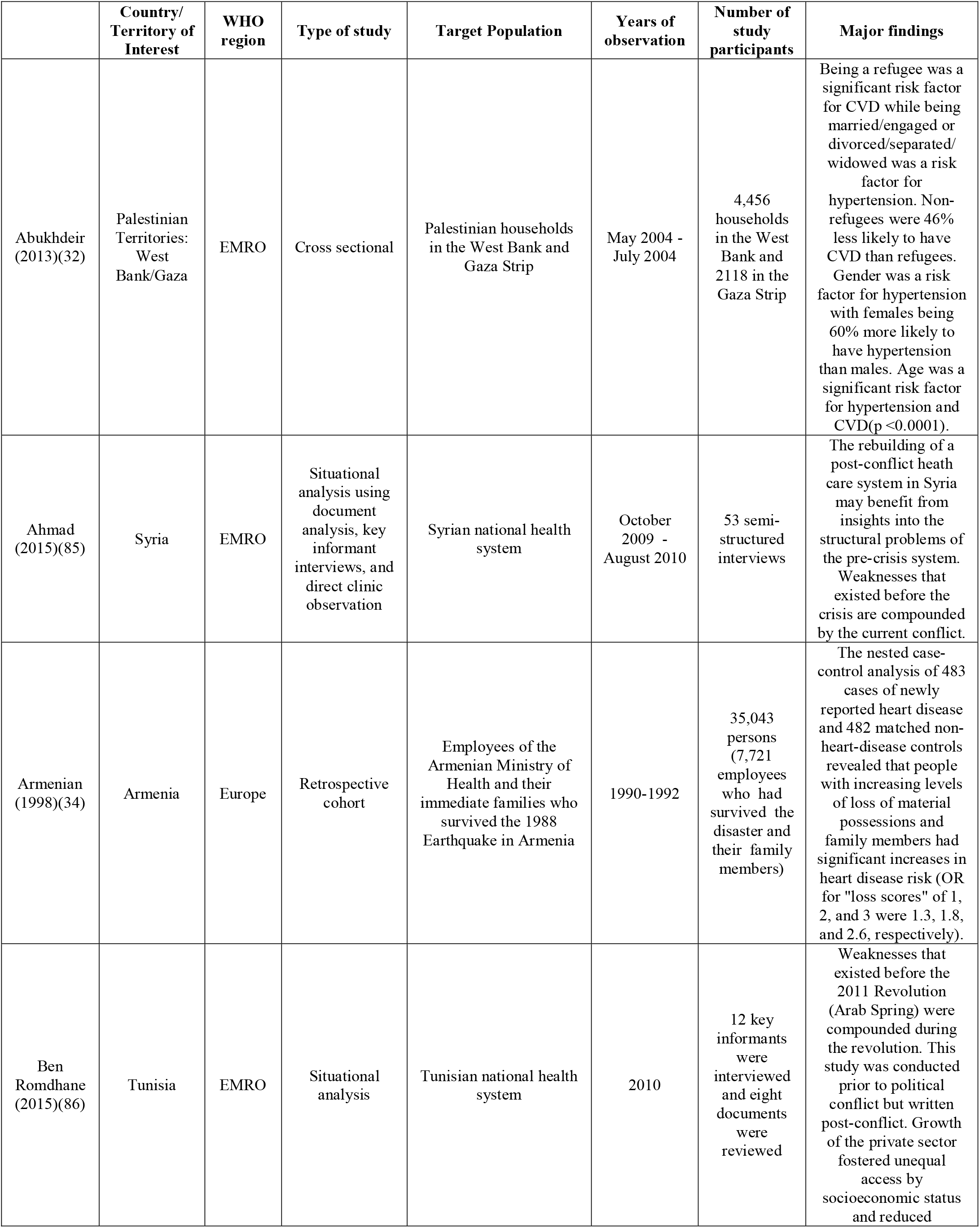
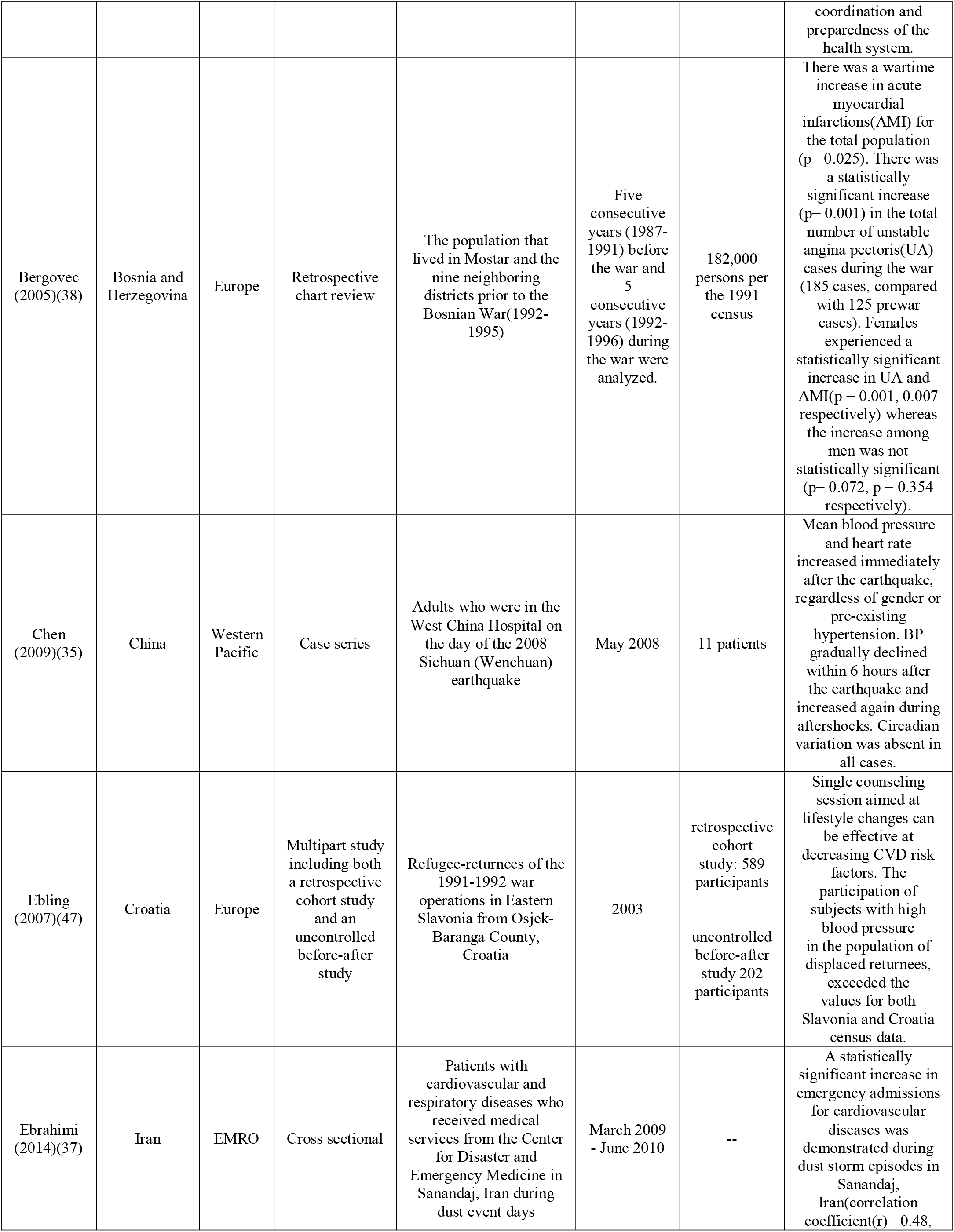
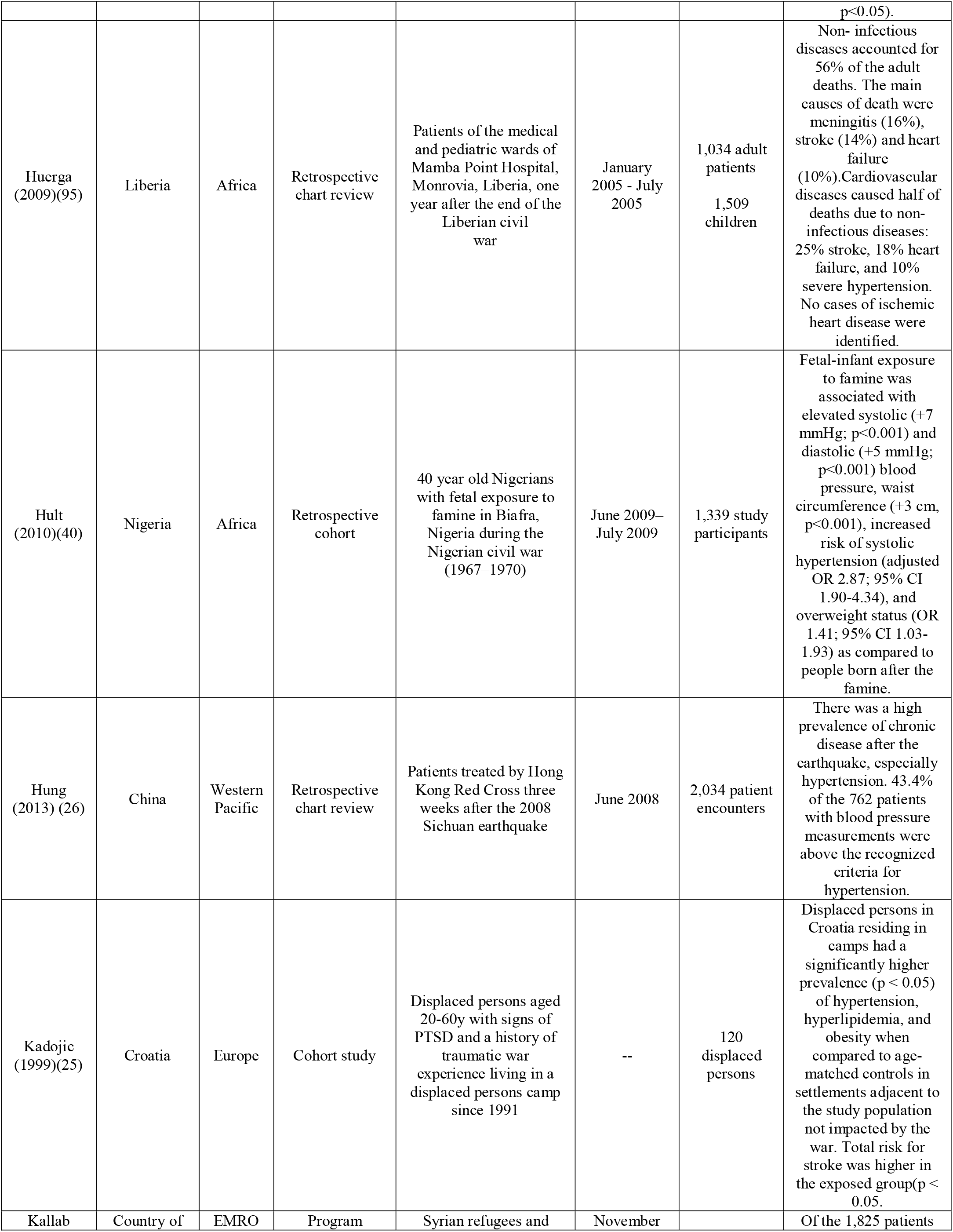
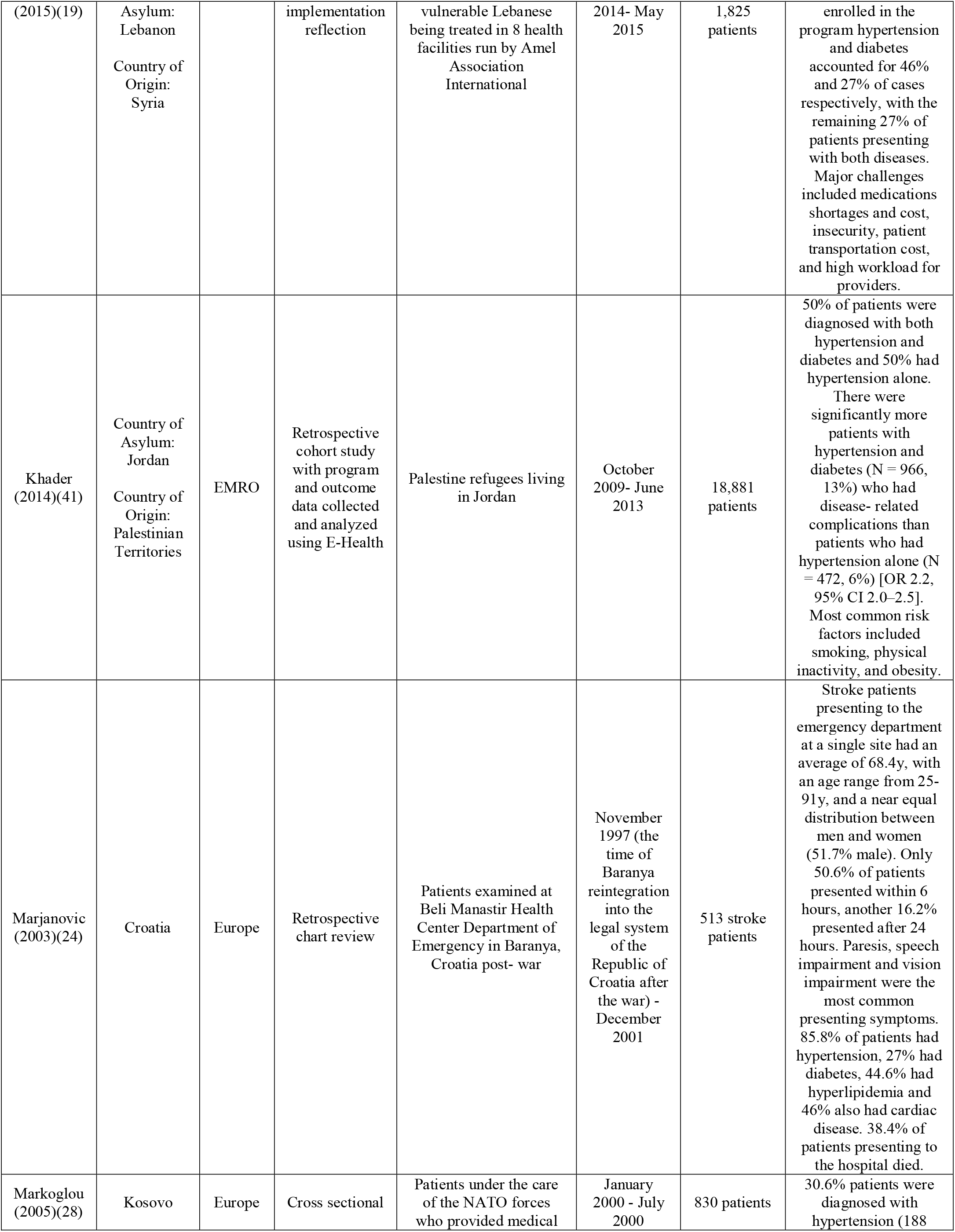
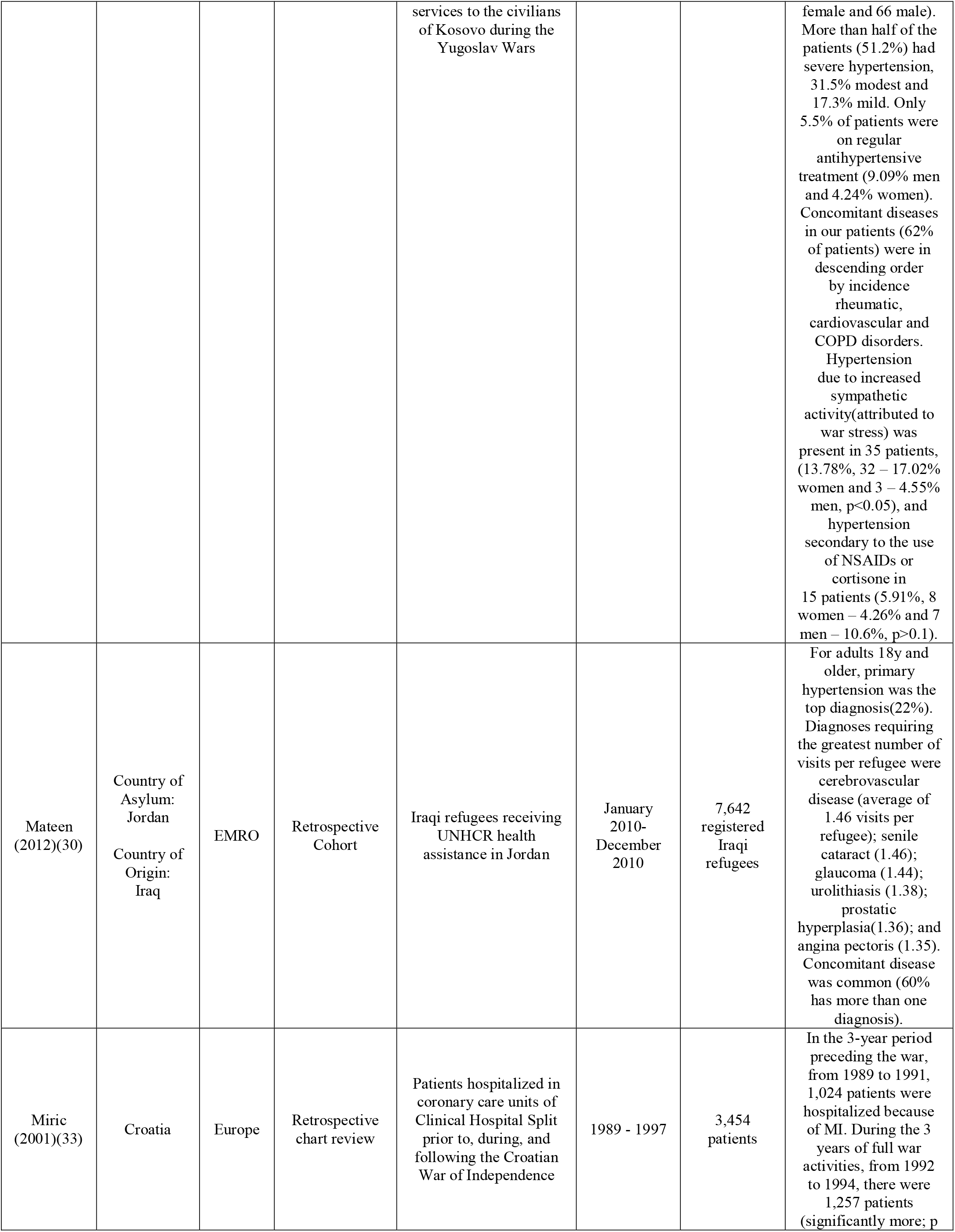
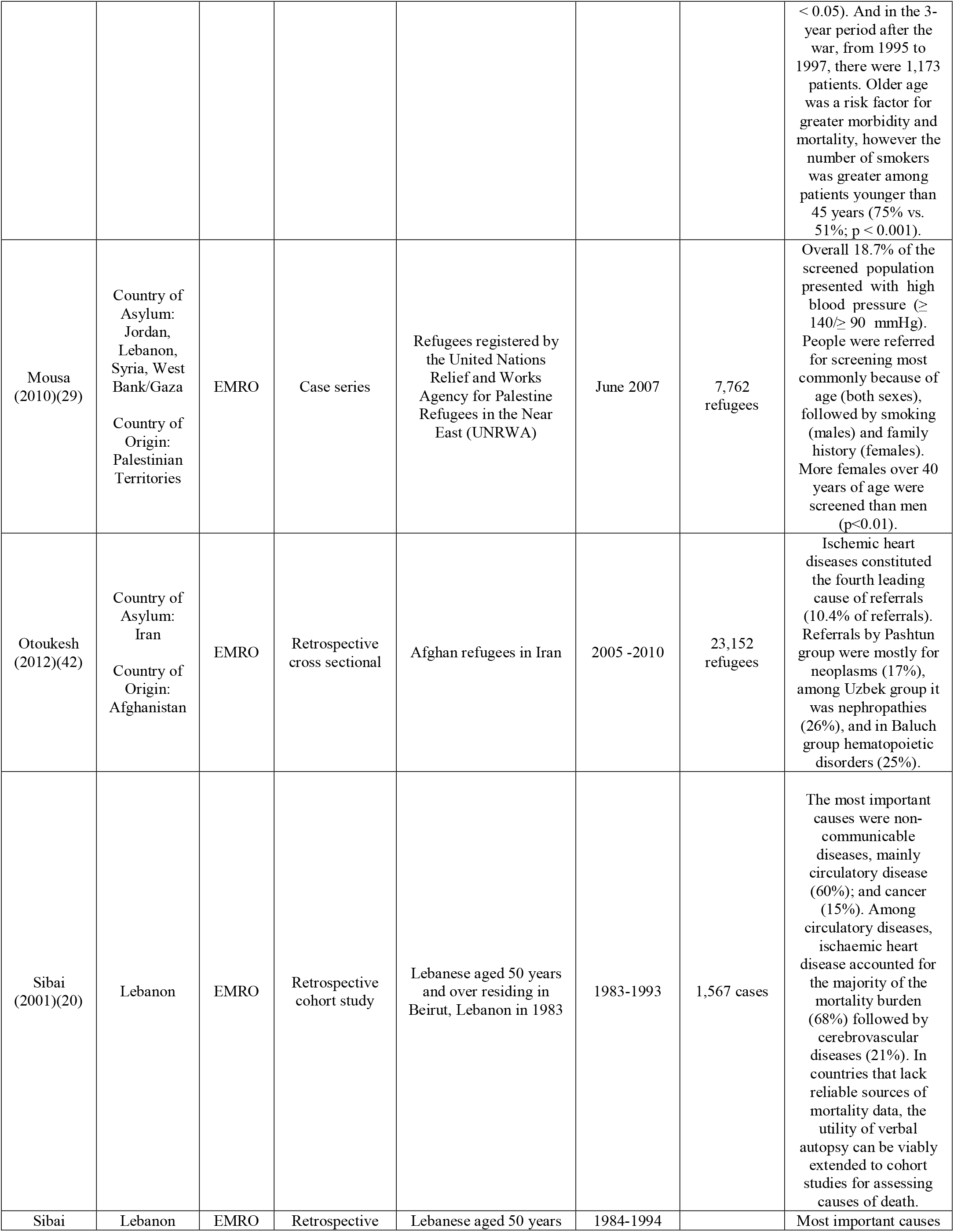
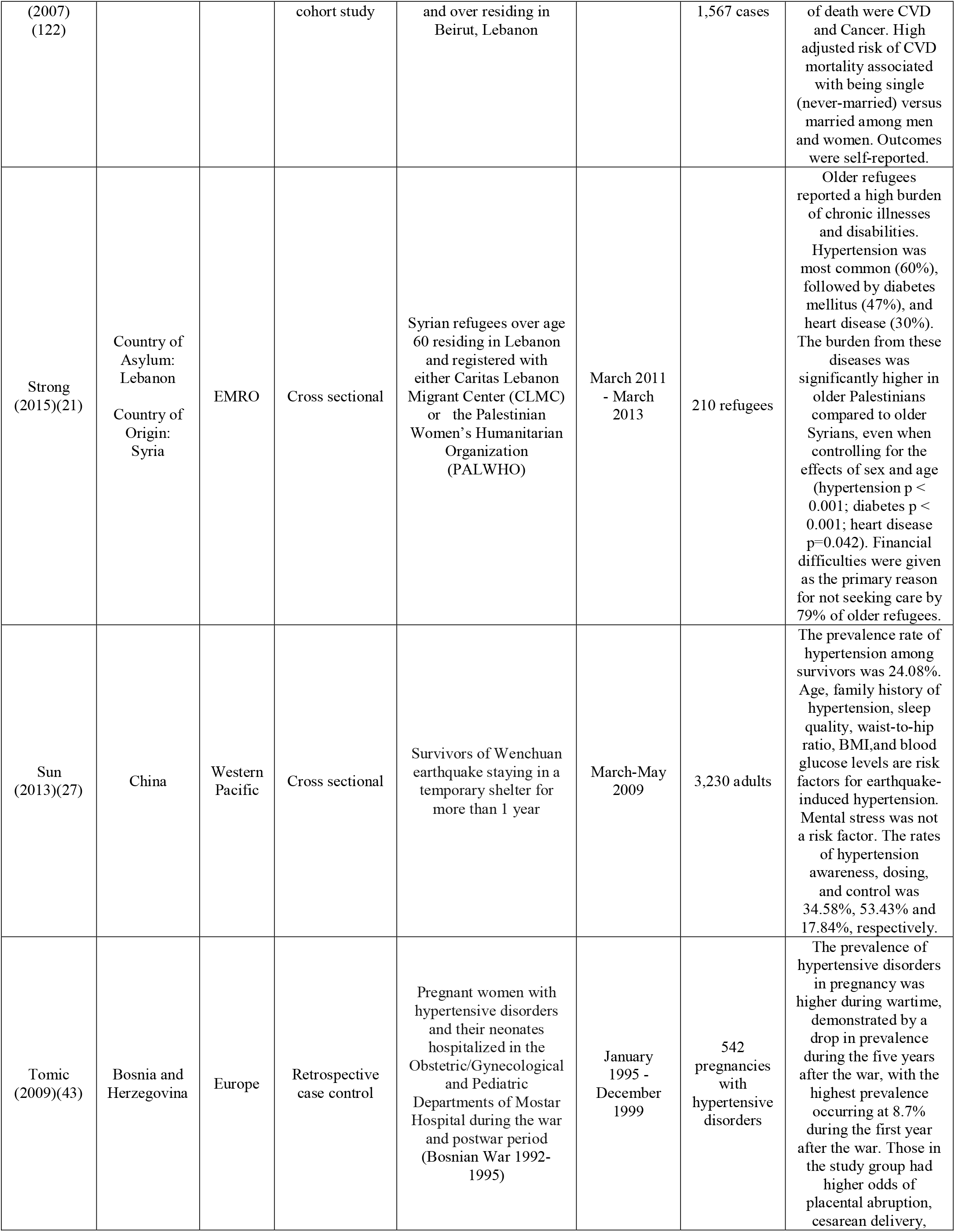
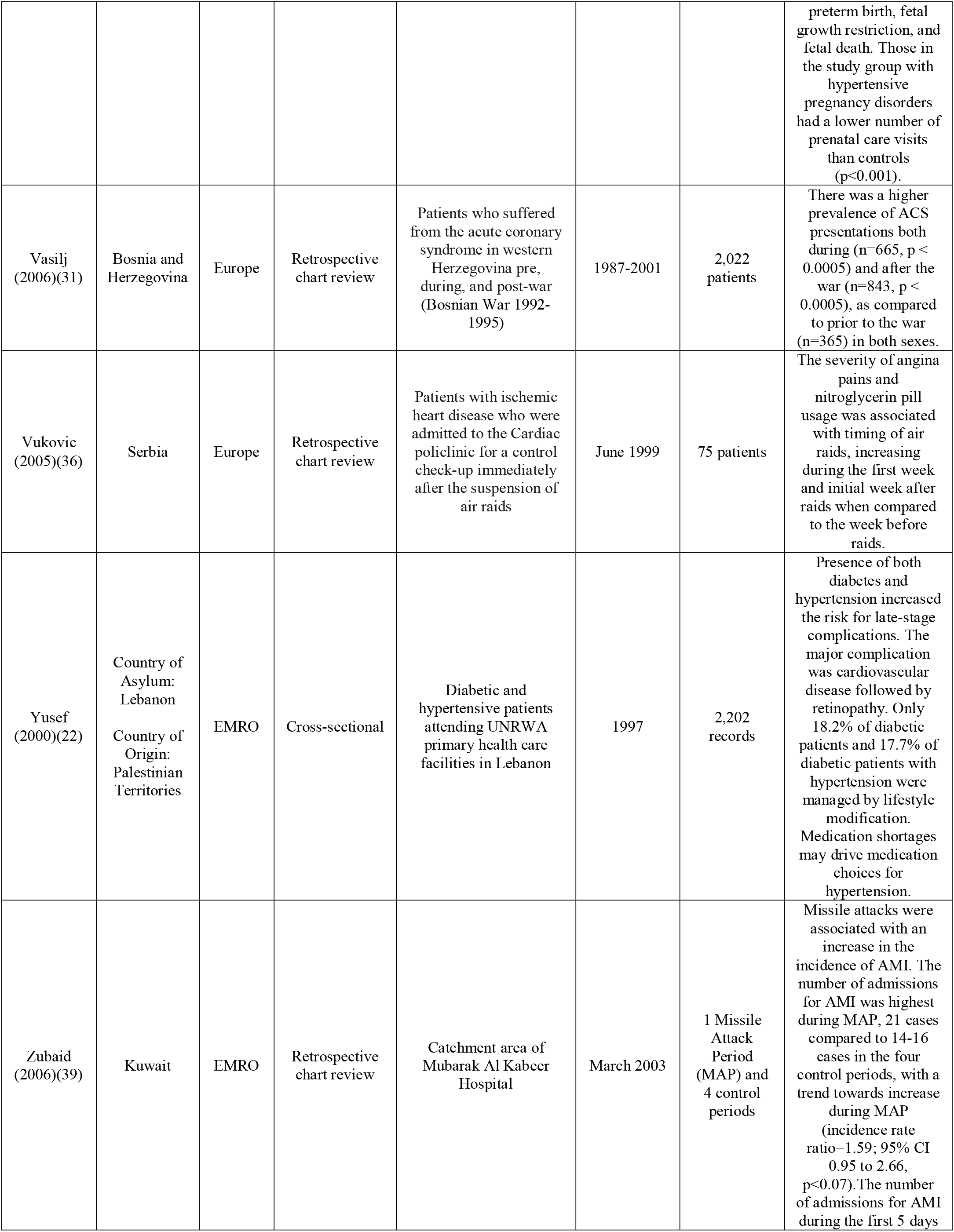
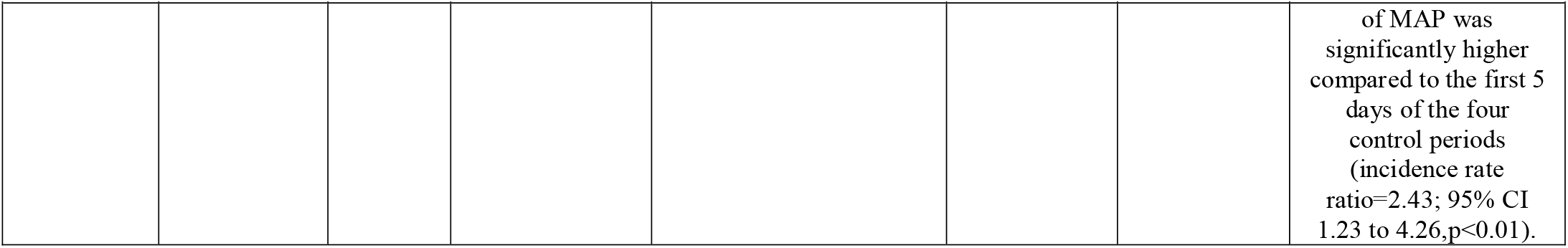
Characteristics of included publications by disease type: Cardiovascular Disease

Most studies assessed cardiovascular disease risk factors, or intermediate risk factors, as opposed to actual diseases such as heart attack or stroke. Intermediate risk factors as defined by the WHO are: raised blood pressure, raised glucose levels, abnormal blood lipids (particularly low density lipoprotein – LDL cholesterol), overweight (body mass index(BMI) ≥25 kg/m2), and obesity (body mass index ≥30 kg/m2) (23). Of note, only two studies primarily addressed strokes (24, 25).

Hypertension remains the lead CVD risk factor (25-27) (19, 21, 28, 29), and reason for presentation for care among refugees as demonstrated by data on Iraqi refugees in Jordan, where for adults 18 and older, primary hypertension was the top diagnosis (30). However, blood pressure control remains a problem, as demonstrated among victims staying in temporary shelter more than 1 year after a 2008 earthquake in the Sichuan province of China, where only half of those diagnosed had medications (53.4%) (27) and less than one in five (17.8%) demonstrated control.

In a study by Marjanovic et al, conducted two years after the 1991-97 Croatia War in the Baranya region of Croatia, they found 513 stroke cases in a single-site emergency department study(24). This was one of only two studies we found that addressed strokes, providing evidence of the downstream effects of intermediate risk factors of high prevalence like hypertension(24, 25). The patients had an average age of 68.4y, with an age range from 25-91y, and a near equal distribution of the cases between men and women (51.7% male). Only 50.6% of patients presented within 6 hours, another 16.2% presented after 24 hours (24), paresis, speech impairment and vision impairment were the most common presenting symptoms. 38.4% died in hospital. 85.8% of patients had hypertension, 27% had diabetes, 44.6% had hyperlipidemia and 46% also had cardiac disease.

The effect of being exposed to a disaster as a primary contributor to CVD also emerged as a trend (31-37). In a Croatian study assessing the patterns of presentations for acute myocardial infarction (AMI) in 3,454 patients, they found a 23% increase (1,254 vs 1,024 hospitalized patients) as compared to the 3-year period preceding the war (1989-1991), and a 15% increase (1,173 hospitalized patients) as compared to the 3-year period following the war (1995-1998) (33). The incidence of hypertension and heart disease is also higher in those with death or injury in their family, with disease occurring most commonly within the first six months after disaster (34). In another study on residents affected by the Bosnian war, they assessed incidence of AMI and unstable angina (UA) 5 years prior to, during, and 5 years after the war (38). The overall incidence of both AMI (n=428 vs 365, p=0.025) and UA (n=185 vs 125, p=0.001) was found to be higher during the war as compared to the period prior. In a Kuwait missile attack, Zubaid et al found that the incidence rate of AMI hospital presentations more than doubled (incidence rate ratio = 2.43; 95% CI: 1.23 – 4.26, p < 0.01) for one year after the event (39). Another study assessing the effects of dust storms in western Iran, showed there was an increase in cardiovascular events with a 1.35% increase in incidence of events for every 100 μg/m3 increase in the PM10 concentration (particulate matter greater than 10μm) (p<0.05) (37). Finally, a retrospective cohort study assessing the effects of famine during the Biafran war (1967-1970) demonstrated association between undernutrition and the presence of hypertension, glucose intolerance, and overweight in Nigerian adults affected (40).

Furthermore, refugee status was associated with higher prevalence of CVD as compared to non-refugee counterparts in several studies. Abukhdeir et al demonstrated a lower prevalence of CVD among those reporting non-refugee status in a representative sample of Palestinian households within the West Bank and the Gaza Strip (OR 0.539, p<0.001), as compared to their refugee counterparts (32). Yusef et al highlight an alarming predominance of late presentations for CVD, and other NCD risk factors, at United Nations Relief and Works Agency(UNRWA) primary health care facilities in Lebanon with 42% of respondents having at least one complication (such as retinopathy, nephropathy, and neuropathy) (22). Similarly, Kadojic demonstrated that displaced persons in Croatia residing in camps had higher prevalence of hypertension, hyperlipidemia and obesity when compared to age-matched controls in settlements not impacted by the war (25).

Regarding CVD risk factors along gender lines, generally men tend to have a higher prevalence of hypertension as compared to women, and associated CVD (myocardial infarction, congestive heart failure, and stroke) (20, 41, 42). However, in several studies we found a trend of disproportionate prevalence and worse outcomes for women for a variety of CVD outcomes (22, 26, 29, 41, 43). Gender differences are described in a separate review.

Only one study addressed management of disease. This was a descriptive analysis by Yusef et al, showing that among refugees accessing care at UNRWA facilities in Lebanon, only 3% were on first-line anti-hypertensive therapy, up to 14.2% were on third line treatment, and 10% reported lifestyle modifications (22). Another study discussed a complex intervention that included capacity-building of staff, provision of key diagnostic tools such as blood pressure cuffs, stethoscopes and glucometers), and advocacy on providing NCD care. The intervention took place in Lebanon (19), and they implemented screening for DM and hypertension in those 40y and older attending any of the clinics (five health centers and three mobile units), with the potential for referral to a specialist, such as cardiology, in case of need. This and the scant other interventions found in our study (19, 44-48, 123) are further addressed in a separate publication.

### Cancer

Multiple studies demonstrated that cancer and oncological emergencies affect populations in conflict (see Table 2). Of the articles, there was a predominant geographic focus on the EMRO region. In Lebanon, Sibai et al (20) observed that cancer was second only to cardiac disease as a cause of death.

**Table 2:** Characteristics of included publications by disease type: Cancer

|  | Country/<br>Territory of<br>Interest | WHO<br>region | Type of study | Target Population | Years of<br>observation | Number of<br>study<br>participants | Major findings |
| --- | --- | --- | --- | --- | --- | --- | --- |
| Huynh<br>(2004)(53) | Vietnam | Western<br>Pacific | Case control | Vietnamese<br>women<br>hospitalized with<br>cervical cancer | June 1996 -<br>September<br>1996 | 145 women in<br>southern<br>Vietnam and<br>80 women in<br>northern<br>Vietnam | The development of<br>invasive cervical<br>cancer was<br>significantly<br>associated with<br>military service by<br>husbands during the<br>Second Indochinese<br>War and with parity<br>status. Geographic<br>and temporal<br>variation in cervical<br>cancer rates among<br>Vietnamese women<br>was associated with<br>the movement of<br>soldiers. |
| Khan<br>(1997)(55) | Country of<br>Asylum: Pakistan<br>Country of Origin:<br>Afghanistan | EMRO | Cross<br>sectional | Patients from<br>North West<br>Pakistan and<br>Afghan refugees<br>attending the<br>Institute of<br>Radiotherapy<br>and Nuclear<br>Medicine,<br>Peshwar | 1990 - 1994 | 13,359<br>patients<br>2988 were<br>Afghan<br>refugees<br>10,371 were<br>adults from<br>North West of<br>Pakistan | In male Afghan<br>refugees, esophageal<br>cancer represented<br>16.6% of the cases,<br>compared to only<br>4.6% of the cases in<br>Pakistani residents.<br>Both Pakistani and<br>Afghani refugee<br>women experienced<br>breast cancer as the<br>most common cancer. |
| Li (2012)(50) | China | Western<br>Pacific | Retrospective<br>cohort | Birth cohorts<br>who were<br>exposed to the<br>1959–1961<br>Chinese famine | 1970-2009 | Population of<br>Zhaoyuan<br>county<br>during the<br>1970–1974<br>death survey<br>and 2,830,866<br>during the<br>2005–2009<br>death survey. | The Zhaoyuan<br>population, which<br>experienced long-term nutritional<br>deficiencies from<br>childhood to<br>adolescence, had<br>increased risk for<br>stomach cancer 15 to<br>20 years after the<br>1959–1961 Chinese<br>famine. The birth<br>cohorts who were<br>exposed to famine or<br>experienced<br>malnutrition had<br>higher stomach<br>cancer mortality rates<br>in later life than the birth cohorts not exposed to malnutrition |
| Marom (2014)(45) | Philippines | Western Pacific | Case series | Patients presenting with head and neck (H&N) tumors to a field hospital in the 'sub-acute' period following a typhoon. | November 2013 | 1844 adult patients examined, 85 (5%) presented with H&N tumors | In a relief mission, despite the lack of clinical and pathological staging and questionable continuity of care, surgical interventions can be considered for therapeutic, palliative and diagnostic purposes |
| McKenzie (2015) (48) | Country of Asylum: Jordan<br>Country of Origin: Iraq, Syria | EMRO | Retrospective cohort | UNHCR registered refugees (Iraqi/Syrian) in Jordan | 2012 - 2013 | 223 refugees | Brain tumors accounted for 13% (n=29) of neuropsychiatric applications, and was the most expensive neuropsychiatric diagnosis overall and per applicant. The ECC denied six applications for reasons of eligibility, cost, and/or prognosis. Of the 20 approved applications, 15% (n=3) were approved for less than the requested amount, receiving on average 39% of requested funds. |
| Milojkovic (2005) (51) | Croatia | Europe | Retrospective cohort | Patients with corpus uteri and cervix uteri cancer and ovarian cancer treated in the Clinical Hospital Osijek | 1984 -2002 | 1455 patients treated for gynecological cancer were analyzed. | Gynecologic cancer incidence according to age shows an increase tendency of cervical cancer in younger women in the post war period. The incidence of corpus cancer and ovary has not changed in the observed periods |
| Otoukesh (2012)(42) | Country of Asylum: Iran<br>Country of Origin: Afghanistan | EMRO | Cross sectional | Afghan refugees in Iran | 2005–2010 | 23,152 refugees | Neoplasms represented 17% of referrals among Pashtun group |
| Shamseddine (2004)(49) | Lebanon | EMRO | Ecological study | Lebanese cancer patients following the 1975 -1990 Lebanese Civil War | 1998 | 4388 cases | Among males, the most frequently reported cancer was bladder (18.5%), followed by prostate (14.2%), and lung cancer (14.1%). In sharp contrast to countries worldwide, bladder cancer was notably high, in particular among males. Among females, breast cancer alone constituted around one third of the total cancer caseload in the country. This was followed by colon cancer (5.8%), and cancer of the corpus uteri (4.8%). The predominance of smoking related cancers highlights the importance of primary preventive strategies aimed at reducing smoking prevalence in Lebanon. |
| Sibai<br>(2001)(20) | Lebanon | EMRO | Retrospective cohort | Retrospective cohort study Lebanese aged 50 years and over residing in Beirut, Lebanon in 1983-1993 during the Lebanese Civil War | 1983-1993 | 1567 cases | In both sexes, the leading causes of death were non-communicable, mainly circulatory diseases (60%) and cancer (15%). |
| Telarovic<br>(2006)(52) | Croatia | Europe | Cross sectional | Patients with CNS tumors admitted to the Department of Neurology of Pula General Hospital, Croatia during wartime | January 1986-December 2000 | 364 patients | There was a statistically significant increase of incidence rate ratios (IRR) of CNS tumors in war period versus the periods before and after war. Higher proportion of metastatic tumors than expected per the authors literature review. Authors relate to stress and PTSD |

Cancer represented 15% of all causes of deaths in their retrospective cohort study of 1,567 Lebanese aged 50 years and over residing in Beirut during the Lebanese Civil War (1975-1990). This was followed post-war by Shamseddine et al (49) who identified an overall crude incidence rate for all cancers combined of 141.4 per 100,000 among males and 126.8 among females, a sharp contrast to earlier estimates made in 1966, of 102.8 and 104.1, respectively (49). Of note, few studies addressed refugees, Internally Displaced Persons (IDPs) or noncombatants, in particular (20, 49). We identified no articles relating to cancer prevalence among refugees in Africa, Asia, or the Americas. No studies addressed palliative care for oncology patients in the disaster setting.

Multiple studies indicate a high prevalence of modifiable cancer risk factors (49-52) in conflict-affected populations that could be targets for future intervention such as Human papillomavirus (HPV) vaccination, anti-tobacco smoking campaigns, and access to adequate nutrient-rich food. Cervical cancer, in particular, was identified as being related to or affected by war (51, 53). For example, in the study by Huynh et al (53), they demonstrate that southern Vietnamese women whose husbands served in the armed forces experienced a more than 160%-290% increase in cervical cancer risk, relative to women whose husbands had not served in the armed forces. The authors attribute the association between male combat activity and cervical cancer as men become reservoirs of high risk subtypes of HPV which cause cervical cancer, acquired during wartime movement patterns (53, 54).

We also found a variety of tobacco-related cancers, which highlights the importance of tobacco cessation. Shamseddine et al (49), found, in reviewing 4,388 new cancer cases in post-civil war Lebanon, that lung cancer was the third most prevalent cancer type. In addition, they highlight that bladder cancer incidence rates are disproportionately higher in Lebanon than in the region, and globally. Breast cancer was listed by multiple studies as the most significant cancer burden amongst women in conflict affected LMICs - including studies relating to Lebanon (49), Afghanistan (55), and Pakistan (55) - demonstrating a clear need amongst refugees for accessible mammography. Tobacco associated cancers were noted as prominent in multiple conflict affected nations and as amenable to prevention efforts through anti-smoking campaigns (49, 55).

Li et al (50) identified a relationship between sustained malnutrition in early life and an increased risk of stomach cancer mortality in later life for survivors of the 1959-1961 Chinese famine. Birth cohorts of Zhaoyuan County, China who were exposed to famine or experienced malnutrition had higher stomach cancer mortality rates 15 to 20 years post-famine as compared to birth cohorts not exposed to malnutrition (50). Proposed mechanisms by the authors for this relationship include a correlation between nutritional deficiency and H. Pylori infection, consumption of foods associated with development of gastric carcinoma in times of famine such as salted meat containing N-nitrosamines or nitrite, vitamin deficiencies, and heavy alcohol use (50).

Relating specifically to refugees, Otoukesh et al (42), provided cancer prevalence data for refugees in a 2012 retrospective cross-sectional study of Afghani refugees residing in Iran. Using demographic and medical data collected between 2005 and 2010 from referrals to the United Nations High Commissioner for Refugees (UNHCR) offices in Iran for Afghani refugees, they found that neoplasms represented 13.3% of all referrals second only to ophthalmic diseases. Likewise, McKenzie et al (48) found that amongst UNHCR registered Iraqi and Syrian refugees in Jordan, brain tumors accounted for 13% of all neuropsychiatric applications. Furthermore, Khan et al found a divergence in the epidemiology of cancer diagnosis from the host population when compared to refugees, with esophageal cancer representing 16.6% of oncological cases amongst male Afghan refugees compared to only 4.6% of cases amongst Pakistani residents(55), and further evidence shows a difference in breakdown by ethnicity exemplified by Pashtun refugees who experienced a disproportionate frequency of referrals for oncologic disease(17%) amongst Afghani refugees residing in Iran despite receiving only two percent of all referrals (42).

Further studies identified challenges specific to refugee populations or subgroups of refugee populations (42, 45, 51, 53, 55). Marom et al (45) described clinical and ethical dilemmas in patients with head and neck cancers presenting to a joint Israeli-Filipino field hospital during the subacute period following a 2013 typhoon in the Philippines. They highlight the importance of awareness of cancer epidemiology in the target country prior to deployment. In this case, it guided the Israeli team’s clinical management such as prioritizing physical examination for cervical nodal metastases based on known prevalence of regional lymph node involvement at presentation in 70% of Filipinos with head and neck cancers (45).

Cost of care as a barrier for refugees with cancers was studied by McKenzie et al (48) who aimed to assess the prevalence and cost of neuropsychiatric disorders among Syrian and Iraqi refugees requiring advanced specialty care in Jordan. The UNHCR funds tertiary level medical care for refugees based on the cost and acuity of required care by means of application to an Exceptional Care Committee (ECC). In reviewing refugee applications for tertiary care to the ECC, McKenzie et al (48) found that brain tumors represented the most expensive neuropsychiatric diagnosis overall ($181,815 USD, $7,905 USD/ applicant). Other referral diagnoses were stroke, psychiatric diagnoses, trauma, infectious diseases, multiple sclerosis, neurodevelopmental abnormalities, and epilepsy.

### Chronic Respiratory Disease

Of the fourteen articles that addressed chronic respiratory disease, six were related to war, and most addressed health hazards faced by refugees or victims of chemical weaponry (see Table 3). The geographic focus of most of these studies was the Middle East, with six studies from Iran alone.

**Table 3:**
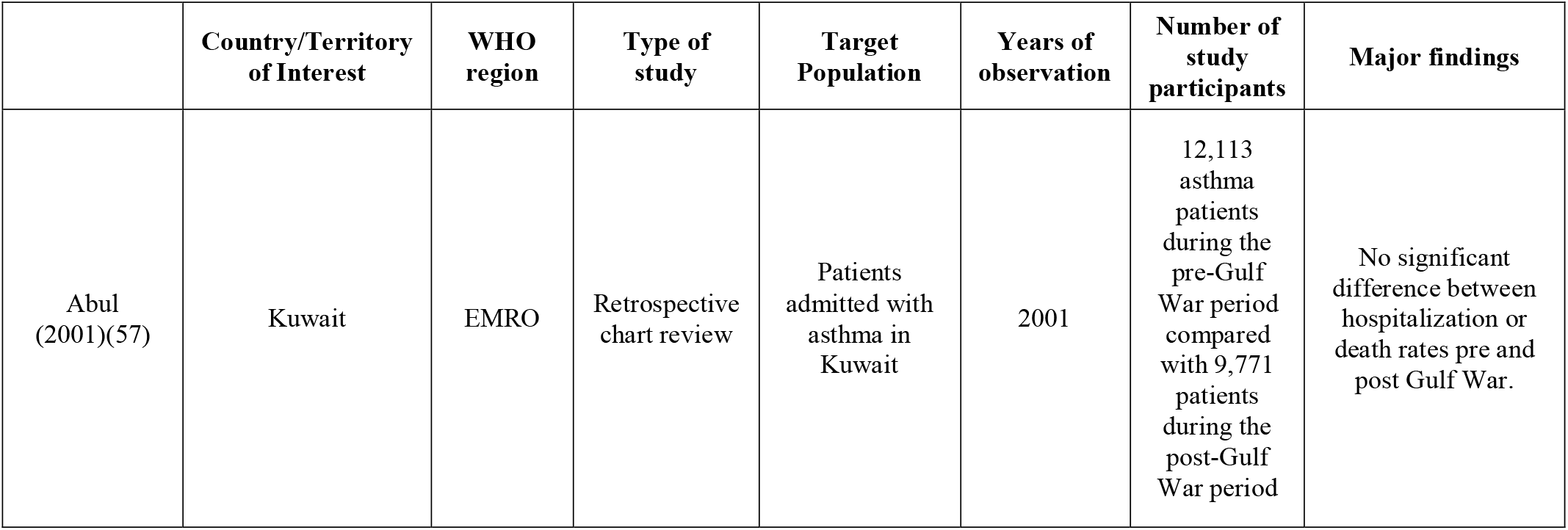
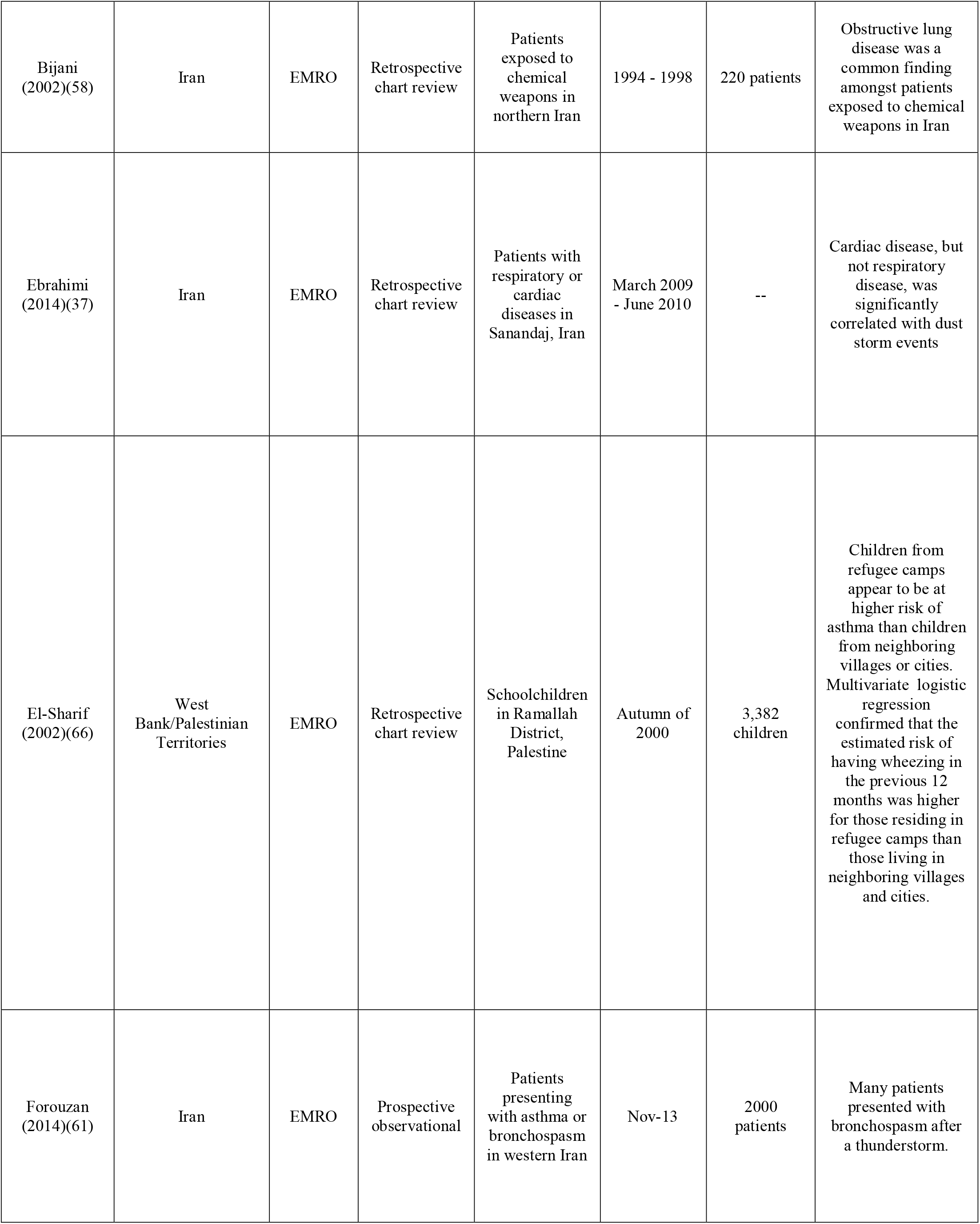
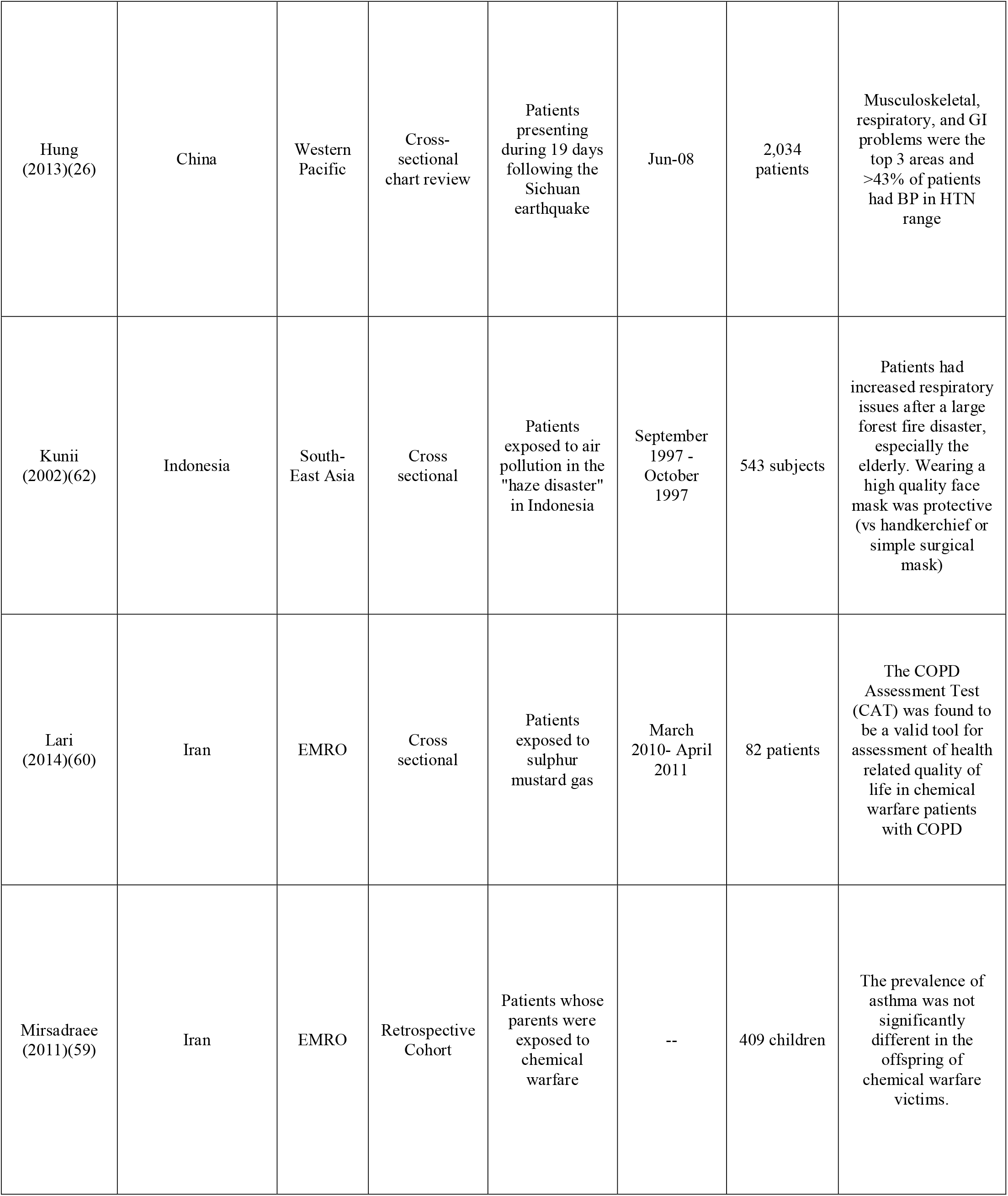
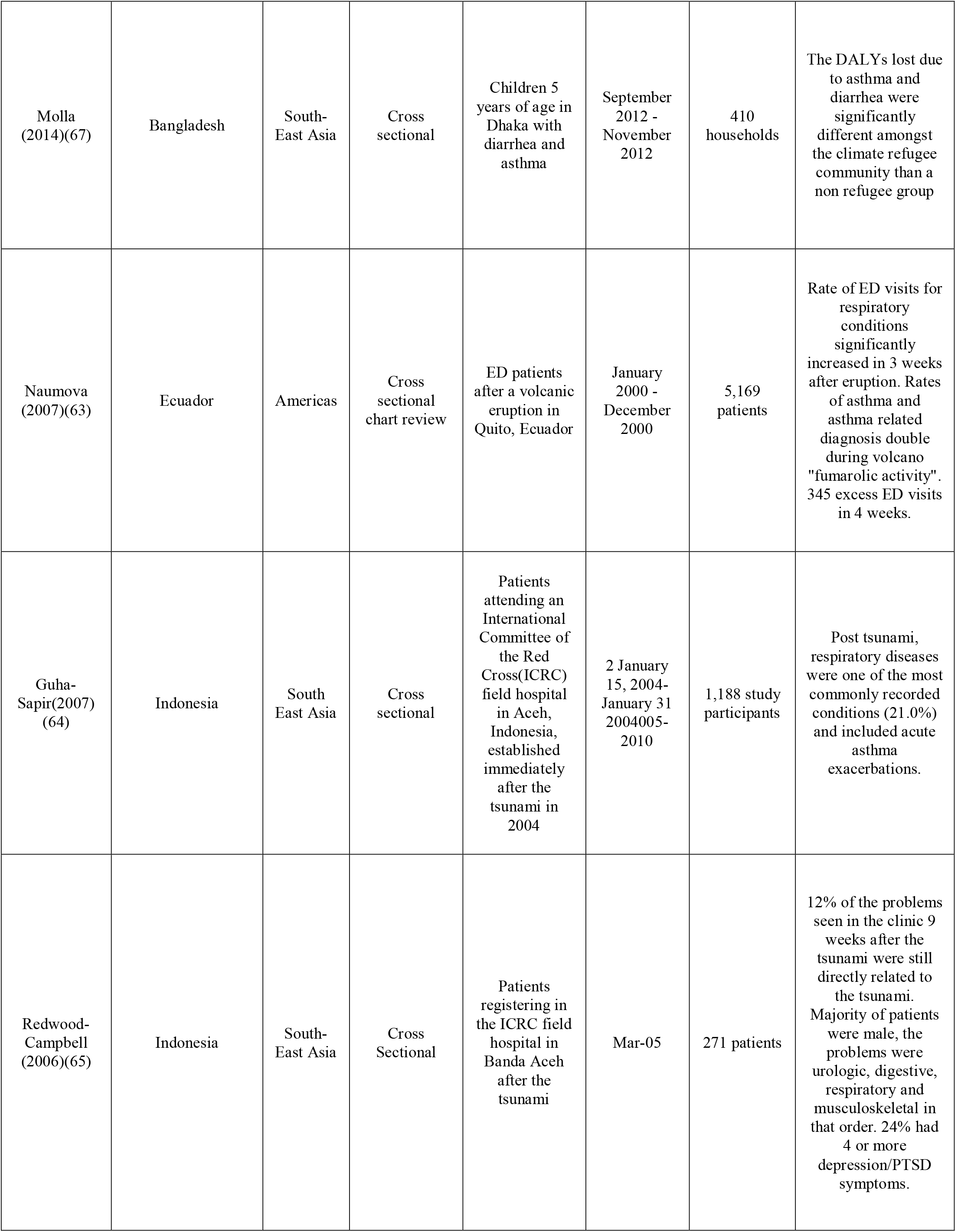
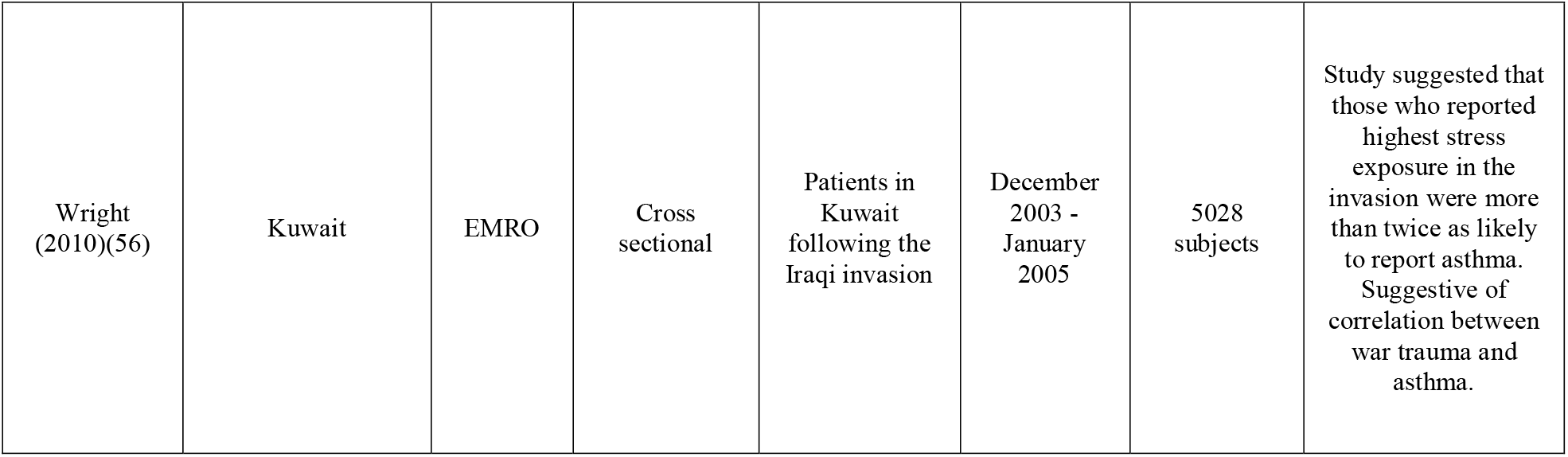
Characteristics of included publications by disease type: Chronic Respiratory Disease

Two studies conducted in Kuwaiti patients affected by the Gulf War demonstrated the association between war trauma and increased in incidence of asthma exacerbations. However, despite the increase in frequency, there was no change in severity of exacerbations. One study found increasing levels of self-reported stress exposure were correlated with reports of asthma (56). In contrast, a chart review on patients admitted with asthma in Kuwait found no difference in admission or mortality rates from asthma when comparing the pre-war and post-war periods (57).

Chemical agents used during warfare, such as sulfur mustard gas, confer an additional risk for chronic respiratory disease (58). In one study assessing incidence of asthma among children of individuals exposed to chemical warfare, a similar incidence of disease was found to that of individuals born to parents with asthma (59). The comparable incidence is concerning for chemical warfare as an independent contributor to the development of asthma. Additionally, a cross-sectional study of a Chronic Obstructive Pulmonary Disease (COPD) cohort demonstrated increased morbidity of patients exposed to sulfur mustard gas also conducted in Iran, and validated use of the COPD Assessment Tool (CAT) for quality of life in this population (60).

The effect of storms on respiratory illness was also studied (37, 61). The only prospective observational study within our review on chronic respiratory disease was on this topic, evaluating asthma exacerbations and bronchospasm associated with thunderstorms in southwestern part of Iran, Ahvaz (61). Two thousand patients who presented with these complaints within three weeks of a thunderstorm were surveyed. This represented an abnormal surge in such complaints for emergency departments there. 30% of patients reported developing their symptoms on the day of the thunderstorm, although only 2% presented within 24 hours. At 3 weeks follow-up, more than two thirds were still using medications, with beta-agonists being the most likely prescriptions, and corticosteroids following. More than half (51.7%) had no prior history of respiratory disease or complaints of shortness of breath. A retrospective chart review similarly looked at respiratory illness and evaluated correlation with dust storms (37). In contrast, this study concluded that cardiac (P <0.05), but not respiratory, disease was associated with occurrence of dust storms.

Beyond storms, a variety of studies looked at the health effects of different types of natural disasters via chart review of patients who presented after the disaster. A large forest fire in Indonesia caused a “haze disaster” in 1997 resulting in increased respiratory complaints. Among 543 respondents, while only 7.4% had a history of chronic respiratory illness (asthma), 98.7% presented with respiratory complaints. 49.2% of all respondents reported symptoms which disturbed their daily life (62). In Ecuador, researchers looked at pediatric emergency department visits and found that there was an increase in frequency of visits associated with volcanic eruptions. Visits for asthma and asthma-related conditions doubled (RR 1.97, 95% CI 1.19, 3.24) during the three weeks following volcanic activity (63). Among NCD presentations to an International Committee of the Red Cross (ICRC) Hospital in Banda Aceh, Indonesia post-tsunami respiratory diseases were one of the most commonly recorded conditions (21%), which included acute asthma exacerbations (64). Similarly, Redwood-Campbell et al (65) cited respiratory complaints as constituting 12% of presentations in the outpatient/ emergency department at the same Indonesian ICRC facility, with asthma making up 29% of those cases.

Studies looking at populations in refugee camps were epidemiologic in nature. In the Palestinian West Bank, children from refugee camps were at higher risk of asthma than children from neighboring villages or cities (66). Having a history of wheezing was reported for 22.1% of children in refugee camps versus 16.5% in cities, and 15.5% in villages. Overall, 8.8% (n=298) of children reported wheezing in the previous year, with a 17.1% lifetime prevalence of wheezing. Similarly, in the slums of Dhaka, Bangladesh, children under 5 who were part of a “climate refugee” community were studied and compared to a non-refugee group. Asthma caused a 1069-fold higher number of disability adjusted life years (DALYs) lost in the group displaced due to climate change in comparison to non-affected populations (67).

### Diabetes

We found that studies addressing diabetes were predominantly conducted in the EMRO Region (see Table 4). Specifically, 20 studies were conducted in the Eastern Mediterranean Region, two studies were conducted in the Caucasus region, three studies occurred in Sub-Saharan Africa, six studies occurred in Asia including South and Southeast Asia, and two studies were conducted in Eastern Europe.

**Table 4:**
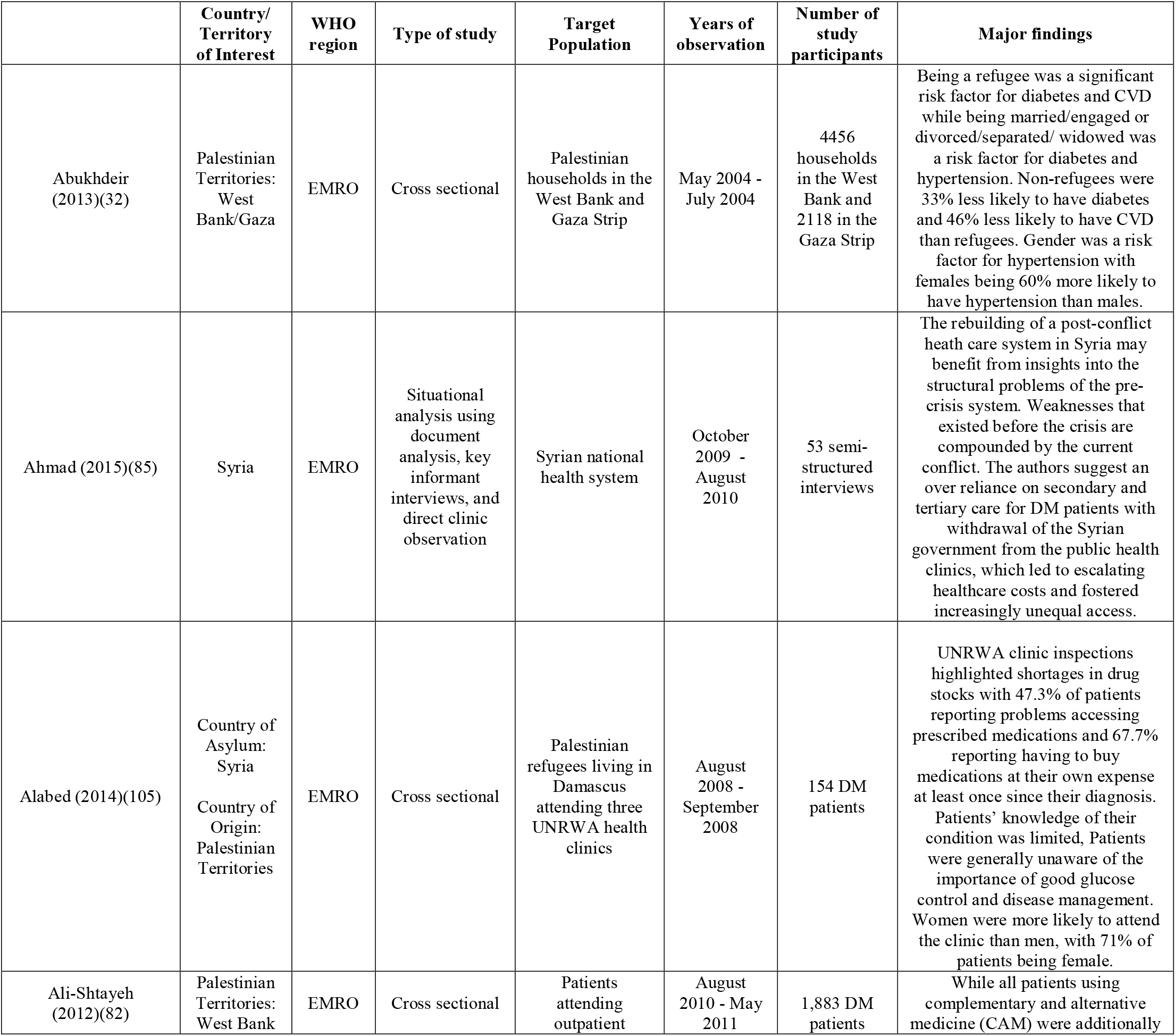
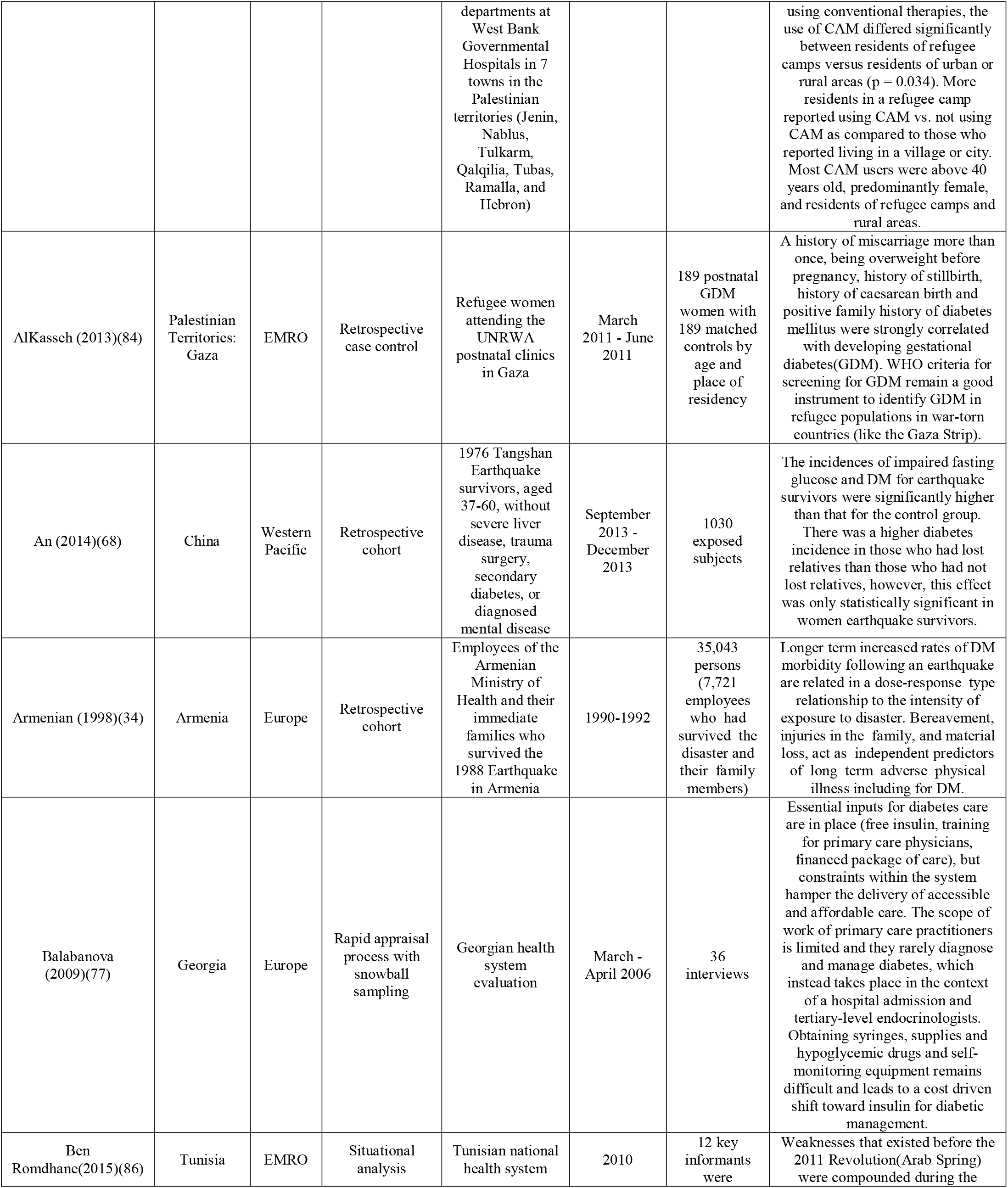
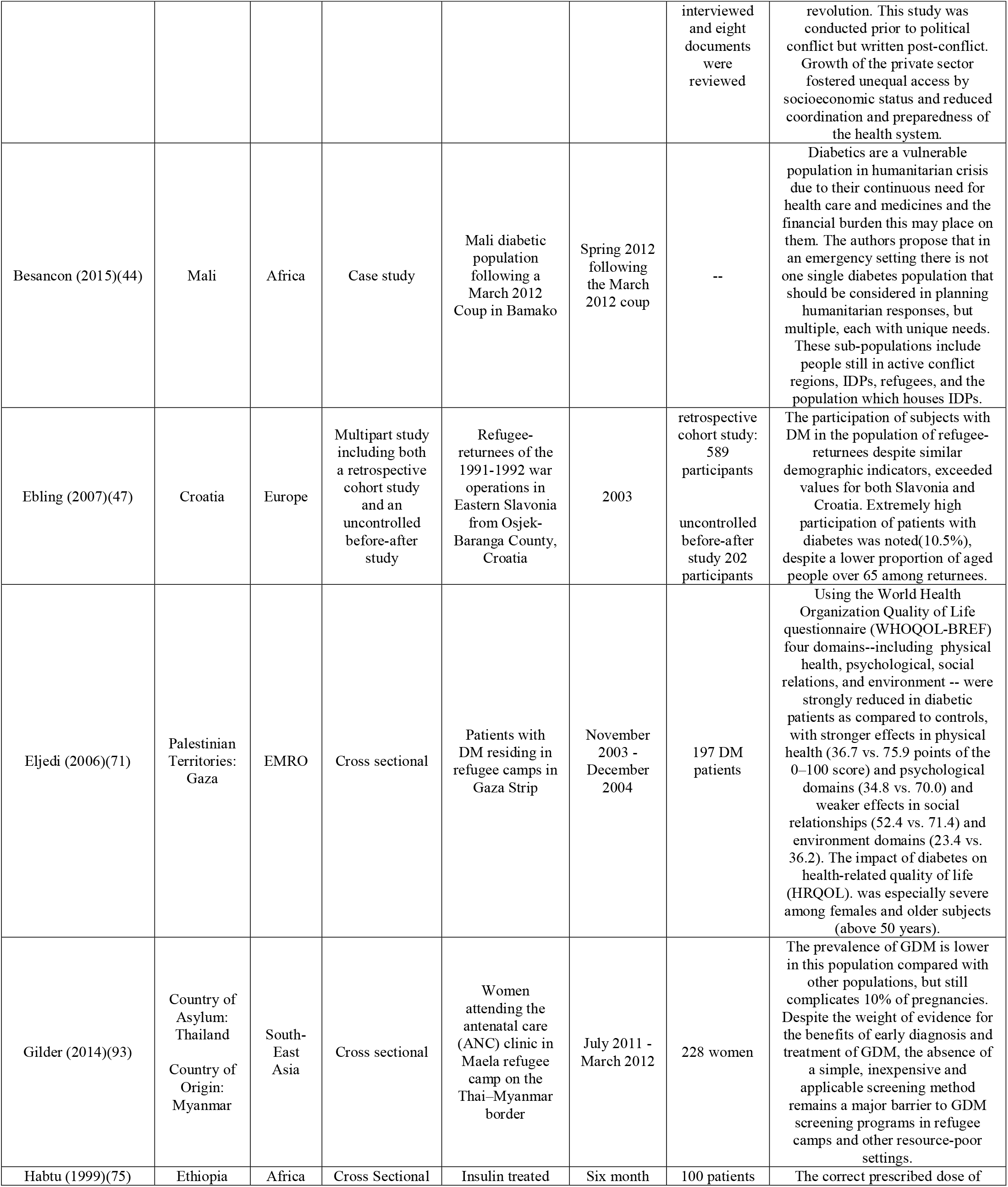
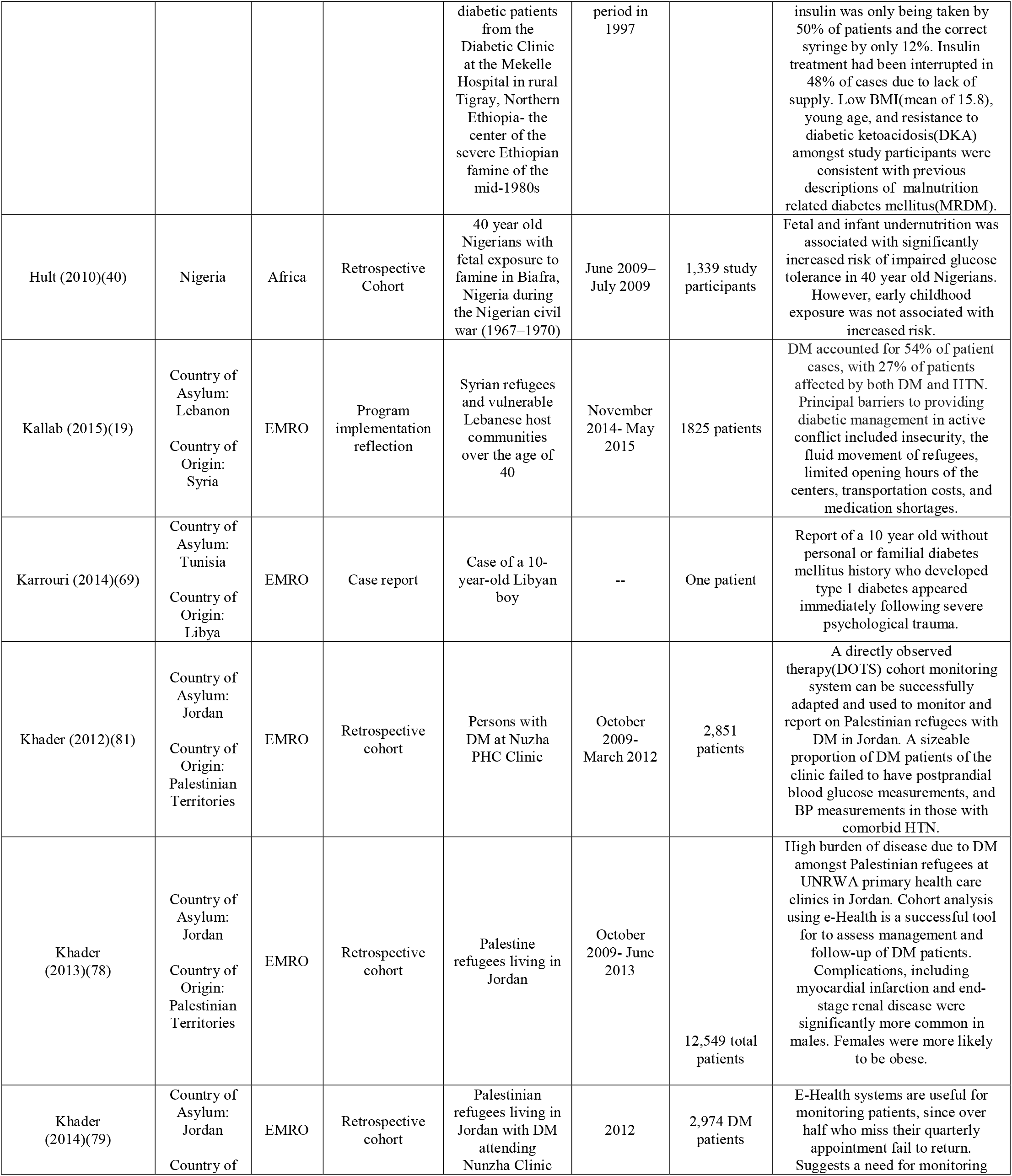
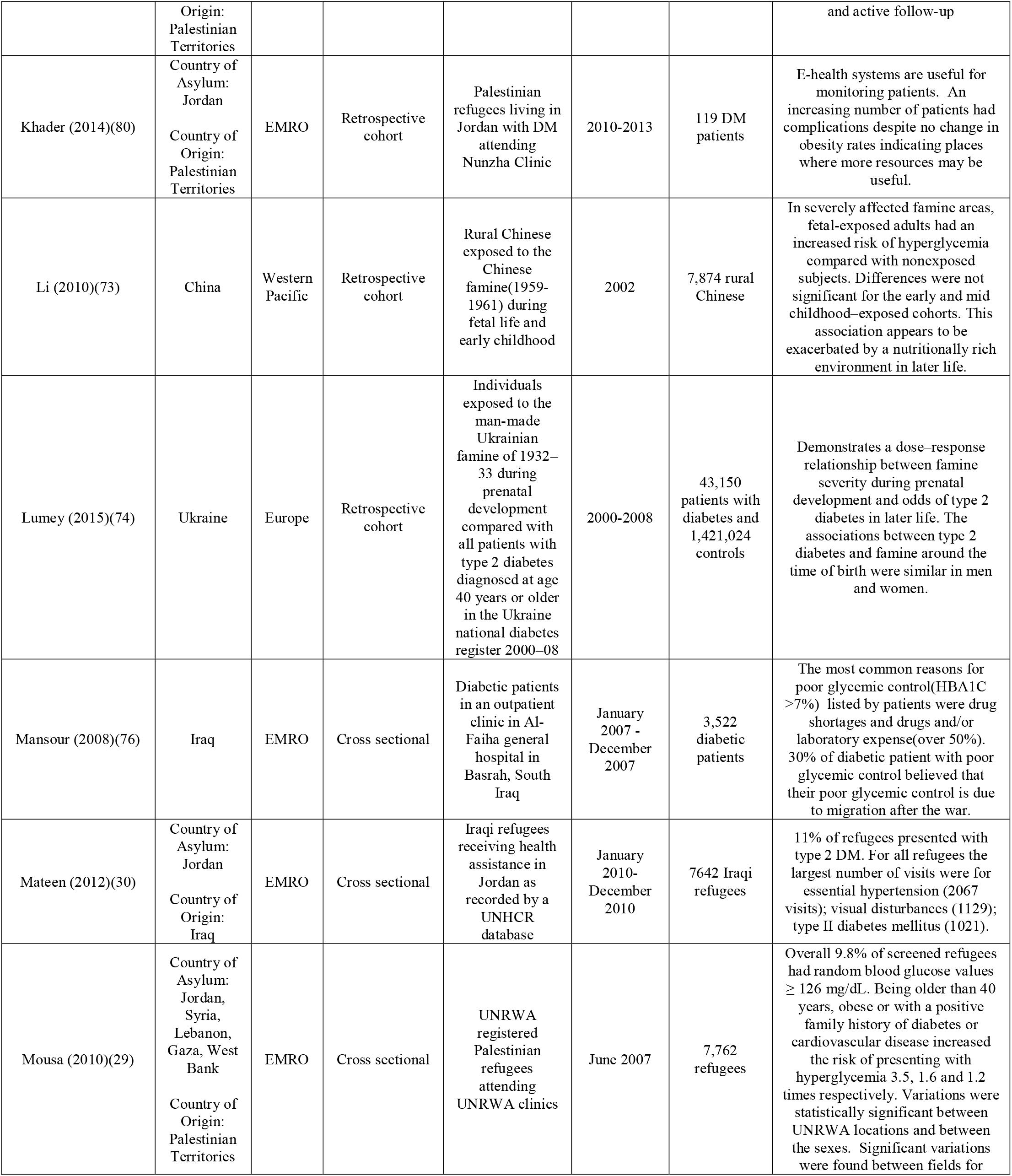
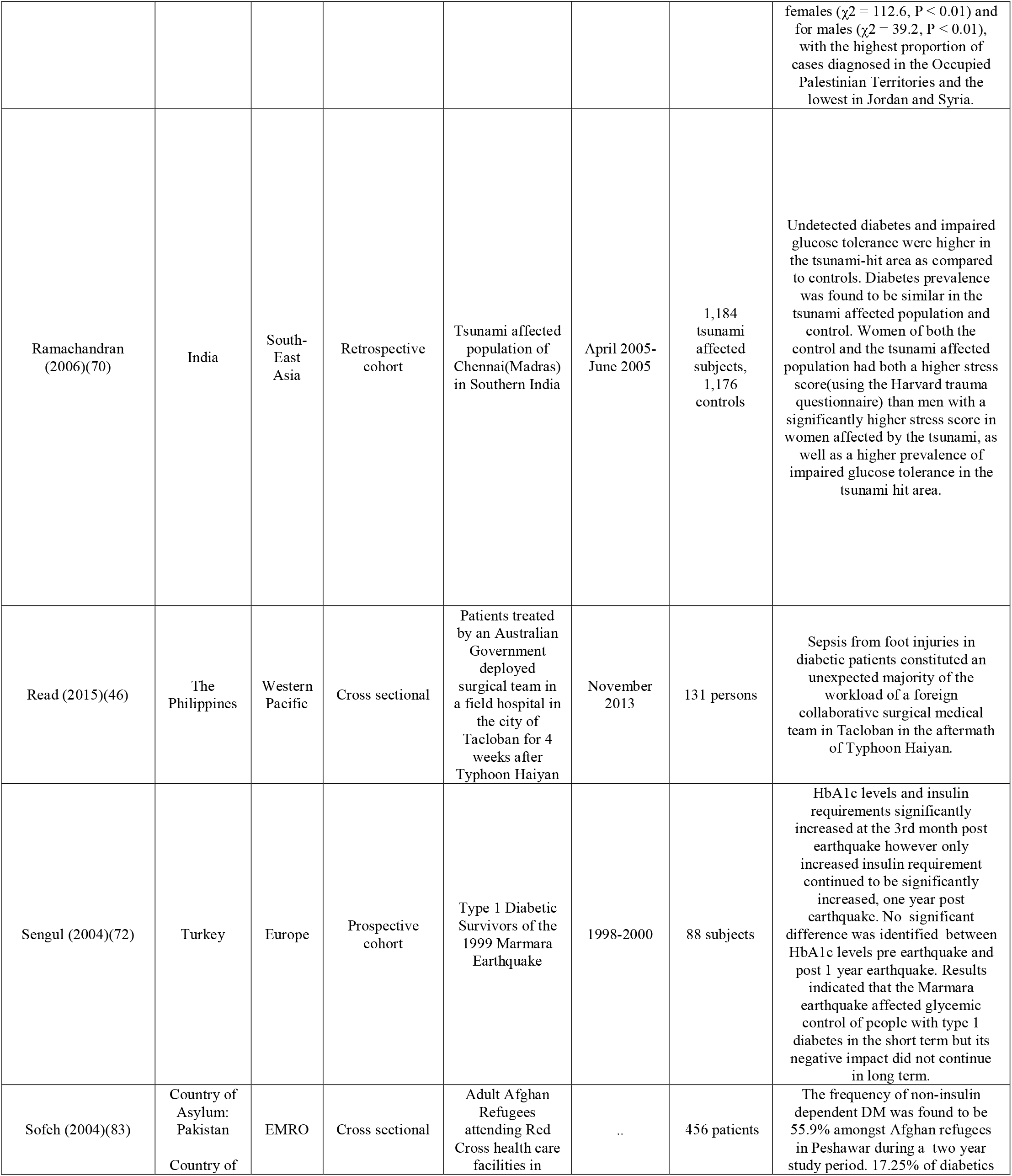
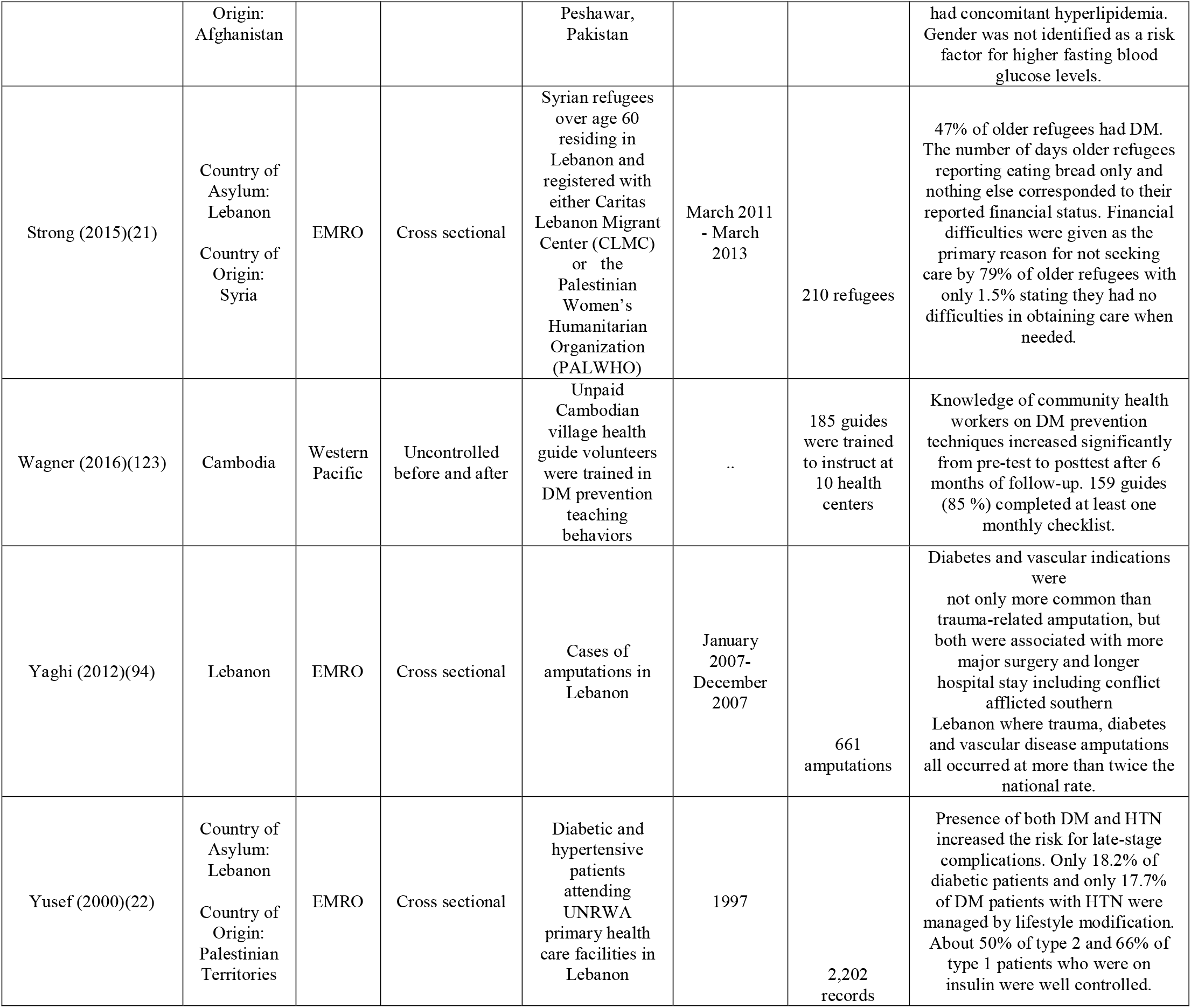
Characteristics of included publications: Diabetes Mellitus

|  | Country/<br>Territory<br>of Interest | WHO<br>region | Type of study | Target<br>Population | Years of<br>observation | Number of<br>study<br>participants | Major findings |
| --- | --- | --- | --- | --- | --- | --- | --- |
| Abukhdeir<br>(2013)(32) | Palestinian<br>Territories:<br>West<br>Bank/Gaza | EMRO | Cross sectional | Palestinian<br>households in the<br>West Bank and<br>Gaza Strip | May 2004 -<br>July 2004 | 4456<br>households<br>in the West<br>Bank and<br>2118 in the<br>Gaza Strip | Being a refugee was a significant<br>risk factor for diabetes and CVD<br>while being married/engaged or<br>divorced/separated/ widowed was<br>a risk factor for diabetes and<br>hypertension. Non-refugees were<br>33% less likely to have diabetes<br>and 46% less likely to have CVD<br>than refugees. Gender was a risk<br>factor for hypertension with<br>females being 60% more likely to<br>have hypertension than males. |
| Ahmad (2015)(85) | Syria | EMRO | Situational<br>analysis using<br>document<br>analysis, key<br>informant<br>interviews, and<br>direct clinic<br>observation | Syrian national<br>health system | October<br>2009 -<br>August<br>2010 | 53 semi-structured<br>interviews | The rebuilding of a post-conflict<br>health care system in Syria may<br>benefit from insights into the<br>structural problems of the pre-crisis system. Weaknesses that<br>existed before the crisis are<br>compounded by the current<br>conflict. The authors suggest an<br>over reliance on secondary and<br>tertiary care for DM patients with<br>withdrawal of the Syrian<br>government from the public health<br>clinics, which led to escalating<br>healthcare costs and fostered<br>increasingly unequal access. |
| Alabed (2014)(105) | Country of<br>Asylum:<br>Syria<br>Country of<br>Origin:<br>Palestinian<br>Territories | EMRO | Cross sectional | Palestinian<br>refugees living in<br>Damascus<br>attending three<br>UNRWA health<br>clinics | August<br>2008 -<br>September<br>2008 | 154 DM<br>patients | UNRWA clinic inspections<br>highlighted shortages in drug<br>stocks with 47.3% of patients<br>reporting problems accessing<br>prescribed medications and 67.7%<br>reporting having to buy<br>medications at their own expense<br>at least once since their diagnosis.<br>Patients' knowledge of their<br>condition was limited, Patients<br>were generally unaware of the<br>importance of good glucose<br>control and disease management.<br>Women were more likely to attend<br>the clinic than men, with 71% of<br>patients being female. |
| Ali-Shtayeh<br>(2012)(82) | Palestinian<br>Territories:<br>West Bank | EMRO | Cross sectional | Patients<br>attending<br>outpatient departments at West Bank Governmental Hospitals in 7 towns in the Palestinian territories (Jenin, Nablus, Tulkarm, Qalqilia, Tubas, Ramalla, and Hebron) | August<br>2010 - May<br>2011 | 1,883 DM<br>patients | While all patients using<br>complementary and alternative<br>medicine (CAM) were additionally using conventional therapies, the use of CAM differed significantly between residents of refugee camps versus residents of urban or rural areas ( $p = 0.034$ ). More residents in a refugee camp reported using CAM vs. not using CAM as compared to those who reported living in a village or city. Most CAM users were above 40 years old, predominantly female, and residents of refugee camps and rural areas. |
| AlKasseh (2013)(84) | Palestinian Territories: Gaza | EMRO | Retrospective case control | Refugee women attending the UNRWA postnatal clinics in Gaza | March 2011 - June 2011 | 189 postnatal GDM women with 189 matched controls by age and place of residency | A history of miscarriage more than once, being overweight before pregnancy, history of stillbirth, history of caesarean birth and positive family history of diabetes mellitus were strongly correlated with developing gestational diabetes(GDM). WHO criteria for screening for GDM remain a good instrument to identify GDM in refugee populations in war-torn countries (like the Gaza Strip). |
| An (2014)(68) | China | Western Pacific | Retrospective cohort | 1976 Tangshan Earthquake survivors, aged 37-60, without severe liver disease, trauma surgery, secondary diabetes, or diagnosed mental disease | September 2013 - December 2013 | 1030 exposed subjects | The incidences of impaired fasting glucose and DM for earthquake survivors were significantly higher than that for the control group. There was a higher diabetes incidence in those who had lost relatives than those who had not lost relatives, however, this effect was only statistically significant in women earthquake survivors. |
| Armenian (1998)(34) | Armenia | Europe | Retrospective cohort | Employees of the Armenian Ministry of Health and their immediate families who survived the 1988 Earthquake in Armenia | 1990-1992 | 35,043 persons (7,721 employees who had survived the disaster and their family members) | Longer term increased rates of DM morbidity following an earthquake are related in a dose-response type relationship to the intensity of exposure to disaster. Bereavement, injuries in the family, and material loss, act as independent predictors of long term adverse physical illness including for DM. |
| Balabanova (2009)(77) | Georgia | Europe | Rapid appraisal process with snowball sampling | Georgian health system evaluation | March - April 2006 | 36 interviews | Essential inputs for diabetes care are in place (free insulin, training for primary care physicians, financed package of care), but constraints within the system hamper the delivery of accessible and affordable care. The scope of work of primary care practitioners is limited and they rarely diagnose and manage diabetes, which instead takes place in the context of a hospital admission and tertiary-level endocrinologists. Obtaining syringes, supplies and hypoglycemic drugs and self-monitoring equipment remains difficult and leads to a cost driven shift toward insulin for diabetic management. |
| Ben Romdhane(2015)(86) | Tunisia | EMRO | Situational analysis | Tunisian national health system | 2010 | 12 key informants were interviewed and eight documents were reviewed | Weaknesses that existed before the 2011 Revolution(Arab Spring) were compounded during the revolution. This study was conducted prior to political conflict but written post-conflict. Growth of the private sector fostered unequal access by socioeconomic status and reduced coordination and preparedness of the health system. |
| Besancon (2015)(44) | Mali | Africa | Case study | Mali diabetic population following a March 2012 Coup in Bamako | Spring 2012 following the March 2012 coup | -- | Diabetics are a vulnerable population in humanitarian crisis due to their continuous need for health care and medicines and the financial burden this may place on them. The authors propose that in an emergency setting there is not one single diabetes population that should be considered in planning humanitarian responses, but multiple, each with unique needs. These sub-populations include people still in active conflict regions, IDPs, refugees, and the population which houses IDPs. |
| Ebling (2007)(47) | Croatia | Europe | Multipart study including both a retrospective cohort study and an uncontrolled before-after study | Refugee-returnees of the 1991-1992 war operations in Eastern Slavonia from Osjek-Baranga County, Croatia | 2003 | retrospective cohort study: 589 participants<br>uncontrolled before-after study 202 participants | The participation of subjects with DM in the population of refugee-returnees despite similar demographic indicators, exceeded values for both Slavonia and Croatia. Extremely high participation of patients with diabetes was noted(10.5%), despite a lower proportion of aged people over 65 among returnees. |
| Eljedi (2006)(71) | Palestinian Territories: Gaza | EMRO | Cross sectional | Patients with DM residing in refugee camps in Gaza Strip | November 2003 - December 2004 | 197 DM patients | Using the World Health Organization Quality of Life questionnaire (WHOQOL-BREF) four domains--including physical health, psychological, social relations, and environment -- were strongly reduced in diabetic patients as compared to controls, with stronger effects in physical health (36.7 vs. 75.9 points of the 0–100 score) and psychological domains (34.8 vs. 70.0) and weaker effects in social relationships (52.4 vs. 71.4) and environment domains (23.4 vs. 36.2). The impact of diabetes on health-related quality of life (HRQOL). was especially severe among females and older subjects (above 50 years). |
| Gilder (2014)(93) | Country of Asylum: Thailand<br>Country of Origin: Myanmar | South-East Asia | Cross sectional | Women attending the antenatal care (ANC) clinic in Maela refugee camp on the Thai–Myanmar border | July 2011 - March 2012 | 228 women | The prevalence of GDM is lower in this population compared with other populations, but still complicates 10% of pregnancies. Despite the weight of evidence for the benefits of early diagnosis and treatment of GDM, the absence of a simple, inexpensive and applicable screening method remains a major barrier to GDM screening programs in refugee camps and other resource-poor settings. |
| Habtu (1999)(75) | Ethiopia | Africa | Cross Sectional | Insulin treated | Six month | 100 patients | The correct prescribed dose of |
|  |  |  |  | diabetic patients from the Diabetic Clinic at the Mekelle Hospital in rural Tigray, Northern Ethiopia- the center of the severe Ethiopian famine of the mid-1980s | period in 1997 |  | insulin was only being taken by 50% of patients and the correct syringe by only 12%. Insulin treatment had been interrupted in 48% of cases due to lack of supply. Low BMI(mean of 15.8), young age, and resistance to diabetic ketoacidosis(DKA) amongst study participants were consistent with previous descriptions of malnutrition related diabetes mellitus(MRDM). |
| Hult (2010)(40) | Nigeria | Africa | Retrospective Cohort | 40 year old Nigerians with fetal exposure to famine in Biafra, Nigeria during the Nigerian civil war (1967–1970) | June 2009– July 2009 | 1,339 study participants | Fetal and infant undernutrition was associated with significantly increased risk of impaired glucose tolerance in 40 year old Nigerians. However, early childhood exposure was not associated with increased risk. |
| Kallab (2015)(19) | Country of Asylum: Lebanon<br>Country of Origin: Syria | EMRO | Program implementation reflection | Syrian refugees and vulnerable Lebanese host communities over the age of 40 | November 2014- May 2015 | 1825 patients | DM accounted for 54% of patient cases, with 27% of patients affected by both DM and HTN. Principal barriers to providing diabetic management in active conflict included insecurity, the fluid movement of refugees, limited opening hours of the centers, transportation costs, and medication shortages. |
| Karrouri (2014)(69) | Country of Asylum: Tunisia<br>Country of Origin: Libya | EMRO | Case report | Case of a 10-year-old Libyan boy | -- | One patient | Report of a 10 year old without personal or familial diabetes mellitus history who developed type 1 diabetes appeared immediately following severe psychological trauma. |
| Khader (2012)(81) | Country of Asylum: Jordan<br>Country of Origin: Palestinian Territories | EMRO | Retrospective cohort | Persons with DM at Nuzha PHC Clinic | October 2009- March 2012 | 2,851 patients | A directly observed therapy(DOTS) cohort monitoring system can be successfully adapted and used to monitor and report on Palestinian refugees with DM in Jordan. A sizeable proportion of DM patients of the clinic failed to have postprandial blood glucose measurements, and BP measurements in those with comorbid HTN. |
| Khader (2013)(78) | Country of Asylum: Jordan<br>Country of Origin: Palestinian Territories | EMRO | Retrospective cohort | Palestine refugees living in Jordan | October 2009- June 2013 | 12,549 total patients | High burden of disease due to DM amongst Palestinian refugees at UNRWA primary health care clinics in Jordan. Cohort analysis using e-Health is a successful tool for to assess management and follow-up of DM patients. Complications, including myocardial infarction and end-stage renal disease were significantly more common in males. Females were more likely to be obese. |
| Khader (2014)(79) | Country of Asylum: Jordan<br>Country of | EMRO | Retrospective cohort | Palestinian refugees living in Jordan with DM attending Nunzha Clinic | 2012 | 2,974 DM patients | E-Health systems are useful for monitoring patients, since over half who miss their quarterly appointment fail to return. Suggests a need for monitoring |

|  | **Country/**<br>**Territory**<br>**of Interest** | **WHO**<br>**region** | **Type of study** | **Target**<br>**Population** | **Years of**<br>**observation** | **Number of**<br>**study**<br>**participants** | **Major findings** |
| --- | --- | --- | --- | --- | --- | --- | --- |
|  | Origin:<br>Palestinian<br>Territories |  |  |  |  |  | and active follow-up |
| Khader (2014)(80) | Country of<br>Asylum:<br>Jordan<br>Country of<br>Origin:<br>Palestinian<br>Territories | EMRO | Retrospective<br>cohort | Palestinian<br>refugees living in<br>Jordan with DM<br>attending<br>Nunzha Clinic | 2010-2013 | 119 DM<br>patients | E-health systems are useful for<br>monitoring patients. An<br>increasing number of patients had<br>complications despite no change in<br>obesity rates indicating places<br>where more resources may be<br>useful. |
| Li (2010)(73) | China | Western<br>Pacific | Retrospective<br>cohort | Rural Chinese<br>exposed to the<br>Chinese<br>famine(1959-1961) during<br>fetal life and<br>early childhood | 2002 | 7,874 rural<br>Chinese | In severely affected famine areas,<br>fetal-exposed adults had an<br>increased risk of hyperglycemia<br>compared with nonexposed<br>subjects. Differences were not<br>significant for the early and mid<br>childhood-exposed cohorts. This<br>association appears to be<br>exacerbated by a nutritionally rich<br>environment in later life. |
| Lumey (2015)(74) | Ukraine | Europe | Retrospective<br>cohort | Individuals<br>exposed to the<br>man-made<br>Ukrainian<br>famine of 1932–<br>33 during<br>prenatal<br>development<br>compared with<br>all patients with<br>type 2 diabetes<br>diagnosed at age<br>40 years or older<br>in the Ukraine<br>national diabetes<br>register 2000–08 | 2000-2008 | 43,150<br>patients with<br>diabetes and<br>1,421,024<br>controls | Demonstrates a dose-response<br>relationship between famine<br>severity during prenatal<br>development and odds of type 2<br>diabetes in later life. The<br>associations between type 2<br>diabetes and famine around the<br>time of birth were similar in men<br>and women. |
| Mansour (2008)(76) | Iraq | EMRO | Cross sectional | Diabetic patients<br>in an outpatient<br>clinic in Al-Faiha general<br>hospital in<br>Basrah, South<br>Iraq | January<br>2007 -<br>December<br>2007 | 3,522<br>diabetic<br>patients | The most common reasons for<br>poor glycemic control(HbA1C<br>>7%) listed by patients were drug<br>shortages and drugs and/or<br>laboratory expense(over 50%).<br>30% of diabetic patient with poor<br>glycemic control believed that<br>their poor glycemic control is due<br>to migration after the war. |
| Mateen (2012)(30) | Country of<br>Asylum:<br>Jordan<br>Country of<br>Origin:<br>Iraq | EMRO | Cross sectional | Iraqi refugees<br>receiving health<br>assistance in<br>Jordan as<br>recorded by a<br>UNHCR<br>database | January<br>2010-December<br>2010 | 7642 Iraqi<br>refugees | 11% of refugees presented with<br>type 2 DM. For all refugees the<br>largest number of visits were for<br>essential hypertension (2067<br>visits); visual disturbances (1129);<br>type II diabetes mellitus (1021). |
| Mousa (2010)(29) | Country of<br>Asylum:<br>Jordan,<br>Syria,<br>Lebanon,<br>Gaza, West<br>Bank<br>Country of<br>Origin:<br>Palestinian<br>Territories | EMRO | Cross sectional | UNRWA<br>registered<br>Palestinian<br>refugees<br>attending<br>UNRWA clinics | June 2007 | 7,762<br>refugees | Overall 9.8% of screened refugees<br>had random blood glucose values<br>≥ 126 mg/dL. Being older than 40<br>years, obese or with a positive<br>family history of diabetes or<br>cardiovascular disease increased<br>the risk of presenting with<br>hyperglycemia 3.5, 1.6 and 1.2<br>times respectively. Variations were<br>statistically significant between<br>UNRWA locations and between<br>the sexes. Significant variations<br>were found between fields for females ($\chi^2 = 112.6$, $P < 0.01$) and for males ($\chi^2 = 39.2$, $P < 0.01$), with the highest proportion of cases diagnosed in the Occupied Palestinian Territories and the lowest in Jordan and Syria. |
| Ramachandran (2006)(70) | India | South-East Asia | Retrospective cohort | Tsunami affected population of Chennai(Madras) in Southern India | April 2005-June 2005 | 1,184 tsunami affected subjects, 1,176 controls | Undetected diabetes and impaired glucose tolerance were higher in the tsunami-hit area as compared to controls. Diabetes prevalence was found to be similar in the tsunami affected population and control. Women of both the control and the tsunami affected population had both a higher stress score(using the Harvard trauma questionnaire) than men with a significantly higher stress score in women affected by the tsunami, as well as a higher prevalence of impaired glucose tolerance in the tsunami hit area. |
| Read (2015)(46) | The Philippines | Western Pacific | Cross sectional | Patients treated by an Australian Government deployed surgical team in a field hospital in the city of Tacloban for 4 weeks after Typhoon Haiyan | November 2013 | 131 persons | Sepsis from foot injuries in diabetic patients constituted an unexpected majority of the workload of a foreign collaborative surgical medical team in Tacloban in the aftermath of Typhoon Haiyan. |
| Sengul (2004)(72) | Turkey | Europe | Prospective cohort | Type 1 Diabetic Survivors of the 1999 Marmara Earthquake | 1998-2000 | 88 subjects | HbA1c levels and insulin requirements significantly increased at the 3rd month post earthquake however only increased insulin requirement continued to be significantly increased, one year post earthquake. No significant difference was identified between HbA1c levels pre earthquake and post 1 year earthquake. Results indicated that the Marmara earthquake affected glycemic control of people with type 1 diabetes in the short term but its negative impact did not continue in long term. |
| Sofeh (2004)(83) | Country of Asylum: Pakistan<br>Country of | EMRO | Cross sectional | Adult Afghan Refugees attending Red Cross health care facilities in | ... | 456 patients | The frequency of non-insulin dependent DM was found to be 55.9% amongst Afghan refugees in Peshawar during a two year study period. 17.25% of diabetics |
|  | Origin:<br>Afghanistan |  |  | Peshawar,<br>Pakistan |  |  | had concomitant hyperlipidemia. Gender was not identified as a risk factor for higher fasting blood glucose levels. |
| Strong (2015)(21) | Country of<br>Asylum:<br>Lebanon<br>Country of<br>Origin:<br>Syria | EMRO | Cross sectional | Syrian refugees<br>over age 60<br>residing in<br>Lebanon and<br>registered with<br>either Caritas<br>Lebanon Migrant<br>Center (CLMC)<br>or the<br>Palestinian<br>Women's<br>Humanitarian<br>Organization<br>(PALWHO) | March 2011<br>- March<br>2013 | 210 refugees | 47% of older refugees had DM. The number of days older refugees reporting eating bread only and nothing else corresponded to their reported financial status. Financial difficulties were given as the primary reason for not seeking care by 79% of older refugees with only 1.5% stating they had no difficulties in obtaining care when needed. |
| Wagner (2016)(123) | Cambodia | Western<br>Pacific | Uncontrolled<br>before and after | Unpaid<br>Cambodian<br>village health<br>guide volunteers<br>were trained in<br>DM prevention<br>teaching<br>behaviors | .. | 185 guides<br>were trained<br>to instruct at<br>10 health<br>centers | Knowledge of community health workers on DM prevention techniques increased significantly from pre-test to posttest after 6 months of follow-up. 159 guides (85 %) completed at least one monthly checklist. |
| Yaghi (2012)(94) | Lebanon | EMRO | Cross sectional | Cases of<br>amputations in<br>Lebanon | January<br>2007-December<br>2007 | 661<br>amputations | Diabetes and vascular indications were not only more common than trauma-related amputation, but both were associated with more major surgery and longer hospital stay including conflict afflicted southern Lebanon where trauma, diabetes and vascular disease amputations all occurred at more than twice the national rate. |
| Yusef (2000)(22) | Country of<br>Asylum:<br>Lebanon<br>Country of<br>Origin:<br>Palestinian<br>Territories | EMRO | Cross sectional | Diabetic and<br>hypertensive<br>patients<br>attending<br>UNRWA<br>primary health<br>care facilities in<br>Lebanon | 1997 | 2,202<br>records | Presence of both DM and HTN increased the risk for late-stage complications. Only 18.2% of diabetic patients and only 17.7% of DM patients with HTN were managed by lifestyle modification. About 50% of type 2 and 66% of type 1 patients who were on insulin were well controlled. |

Multiple studies point to the relationship between stress and personal loss incurred in natural disasters and conflict, and a subsequent rise in occurrences of impaired fasting glucose (IFG) and diabetes mellitus (DM) among survivors (34, 47, 68-70). One such retrospective cohort study by An et al. (68) investigated the long-term impact of stresses from the 1976 Tangshan earthquake on the occurrences of impaired IFG and DM among survivors and found that the incidences of IFG and DM for the exposure groups were significantly higher than that for the control group (P = 0.043 for IFG; P = 0.042 for diabetes), with those who had lost relatives exhibiting a higher diabetes incidence than those who had not lost relatives. This effect was only statistically significant in women earthquake survivors (p=0.009) (68). In addition, refugees with diabetes were found to have strongly reduced quality of life (HRQOL) as compared to age-matched non-diabetic controls as identified by Eljedi et al. using the World Health Organization Quality of Life questionnaire (WHOQOL-BREF), with particularly severe effects noted among females (p < 0.05 in all four domains) (71).

Additionally, several studies addressed food insecurity, and identified it as a primary contributing factor affecting diabetes management (21, 29, 47, 72). A study focusing on older Palestinian refugees (21) found that participants practiced reduced meal portion sizes, skipping a meal, or foregoing a full day’s meals due to food shortage at a significantly higher rate than an age matched host population in Syria (reducing portion sizes p <0.001; skipping a meal p<0.001; not eating at all p <0.001). Factors associated with skipped meals or reduced portion sizes included low economic status, larger household size, and type of residence (financial status p=0.009; household size p <0.001; type of residence p <0.001). The number of days older refugees reported eating only bread and nothing else corresponded to reported financial status (p=0.036). The authors theorized that food insecurity may result in challenges in the management of diabetes.

Further studies specifically addressed effects of fetal exposure to malnutrition and impaired glucose tolerance or diabetes later in life (40, 73-75). Hult et al. (40) examined the accumulated risk for glucose intolerance 40 years following fetal exposure to famine in Biafra, Nigeria during the Nigerian civil war. The crude odds ratios for both impaired glucose tolerance and diabetes diagnoses were significantly higher for the group exposed to fetal or infant famine in comparison to controls (40).

Consistent findings were identified by a retrospective cohort study from China by Li et al. (73), who also identified a relationship between the severity of famine for fetal exposed subjects and risk of hyperglycemia later in life (OR = 3.92; 95% CI: 1.64–9.39; *P* = 0.002). Similarly, in a region of Northern Ethiopia recently affected by severe famine, clinical features of 100 insulin-treated diabetic patients were consistent with previous descriptions of malnutrition-related diabetes mellitus (MRDM): young age of onset (70% < 30 yrs), low BMI(mean 15.8), and resistance to ketosis (only 4% admitted with diabetic ketoacidosis despite 48% reporting insulin treatment interruption) (75).

Additional barriers to glycemic control in patients affected by conflict were: migration after war, lack of self-monitoring glucose strips, lack of access and cost of medications, failure to adequately screen for diabetes, inability to travel to a heath facility, lack of education regarding diabetes complications and management, food availability, and difficulty following patients over time (21, 22, 29, 41, 44, 47, 75-81). One cross sectional study (76) which aimed to identify barriers to glycemic control from the patient perspective in a diabetic clinic in the south of Iraq, found that lack of drug supply from a primary health care center or drug shortage is a barrier for 50.8% of patients, while drug and/or laboratory expenses were a barrier for 50.2% of patients. 30.7% of patients said that they were not aware of possible diabetic complications and 30% thought that their failure to control their diabetes was due to migration after the war. Lack of electricity, lack of access to blood glucose monitoring devices, and illiteracy as a cause were cited by 15%, 10.8% and 9.9% respectively (76). In Mali and Ethiopia, insulin was not widely available and access was limited by cost (US$ 11 per vial in Mali)(44, 75). Multiple studies noted that syringes and self-monitoring blood glucose devices were not readily available and posed a financial burden to those who required access to them (44, 75, 77).

One study investigated complementary and alternative medicine (CAM) use among Palestinian diabetic patients and found the use of CAM differed significantly between residents of refugee camps as compared to residents of urban or rural areas (p=0.034)(82). Those who were on CAM reported they were using it to slow down the progression of disease or relieve symptoms and 68% of patients interviewed reported not disclosing CAM use to their physician or pharmacist.

While no study specifically aimed to focus on gender in their primary research objectives, we found a relationship between gender and prevalence or access to resources for diabetes, emerged as a recurring theme (22, 32, 68, 70, 71, 75, 83). These findings will be presented in a separate publication. Other common risk factors associated with diabetes type 2 included age, having a higher BMI, being divorced/widowed/separated, having never attended school, illiteracy, comorbid hypertension, hyperlipidemia, family history, sedentary lifestyle, history of traumatic exposure, and refugee status (21, 22, 29, 32, 69, 83, 84).

Several studies also took a health systems approach and found that reliance on tertiary care for diabetes management fostered unequal access by socioeconomic status, geographic location, and escalating healthcare costs overall (44, 77, 85, 86). One study from Georgia (77),which sought to identify the extent to which the Georgian health system provides for effective diabetes control post-independence, identified a systems level concern that only tertiary-level endocrinologists were able to modify treatment regimens and prescribe insulin whereas even endocrinologists who worked in polyclinics were unable to determine insulin regimens or prescribe insulin. Three studies from Syria (85), Tunisia(86), and Mali (44) identified a similar shift of diabetes care to the tertiary level prior to the emergence of conflict in these countries due to an emerging private sector(85, 86) and lack of specialists(44), respectively. In Mali, the lack of specialists was augmented by a lack of available guidelines, treatment protocols, and training for primary care level providers which prevented a transition of care to primary or general practitioners(44). The authors theorized that this shift of diabetes care to the tertiary level contributed to reduced care access during active conflict in these countries(44, 85, 86).

### Other NCDs

Studies investigating other NCDs centered on musculoskeletal and joint disorders (26, 30, 34, 65) epilepsy and other neuropsychiatric disorders (42, 48, 87), ophthalmic diseases (30, 42, 88), nephropathies and urologic complaints (30, 42, 65) (see Table 5). Two studies measured mortality rates (20, 34) and two also studied quality of life (89, 90). The effects of disability were briefly touched on by Leeuw et al (90), with Amini et al (89) further identifying hearing loss, and tinnitus as having negative impacts on quality of life among blind survivors from the Iranian War (p=0.005, p<0.0001) as compared to non-afflicted counterparts. We found that the majority of the studies on other NCDs did not refer to specific diseases or illnesses (26, 30, 65, 91), but rather represented epidemiological studies referring to conditions more broadly such as in the case of Mateen et al (30) referring to “joint disorders”, and Hung and Redwood-Campbell describing “musculoskeletal”, “respiratory complaints,” and “gastrointestinal complaints” of unclear etiology (26, 65).

**Table 5:**
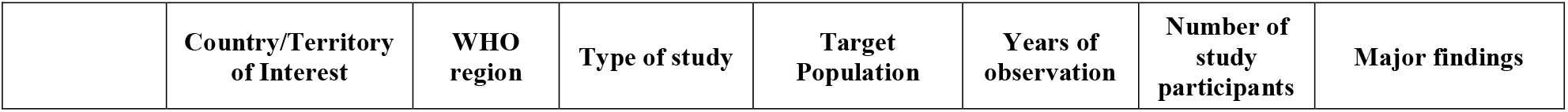
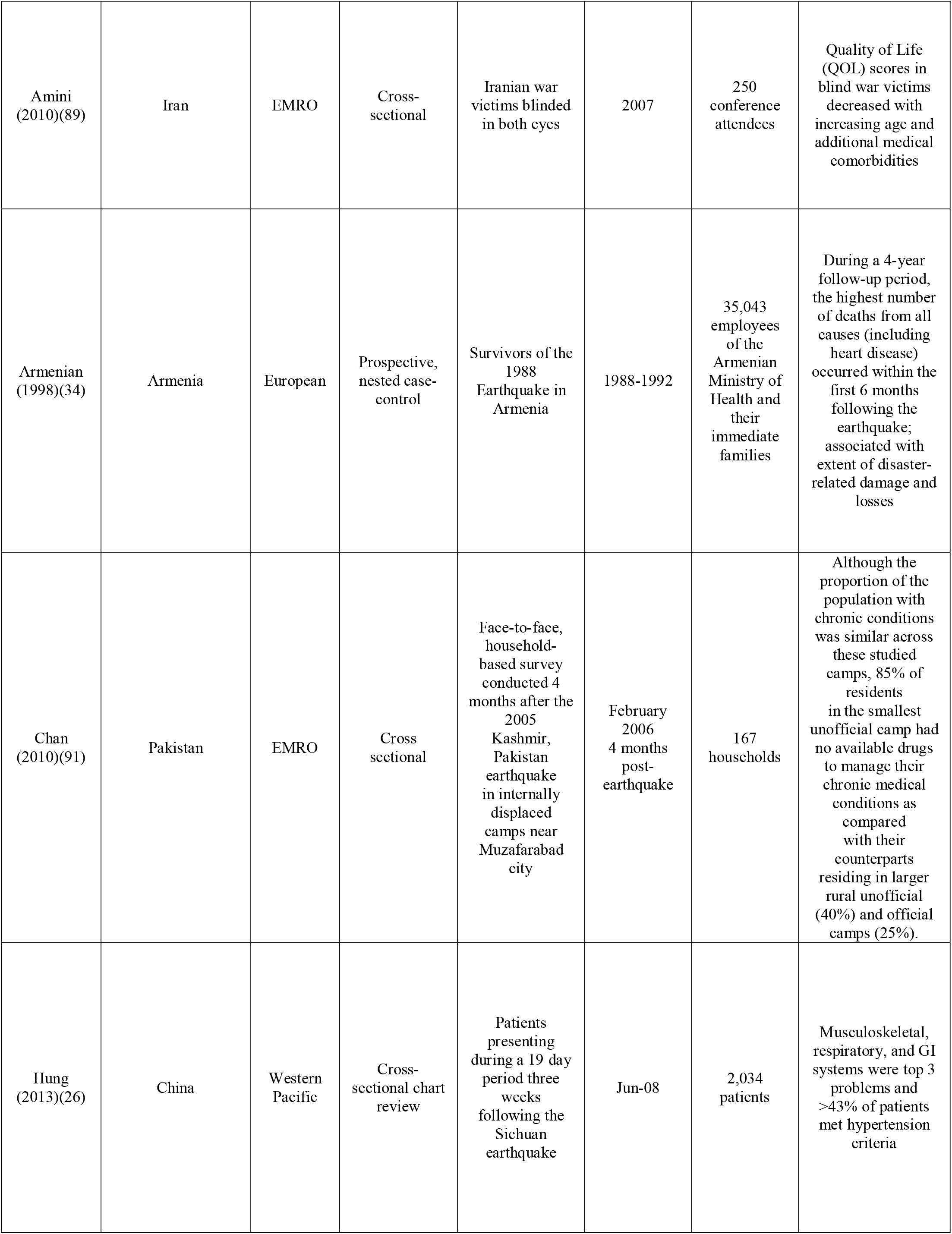
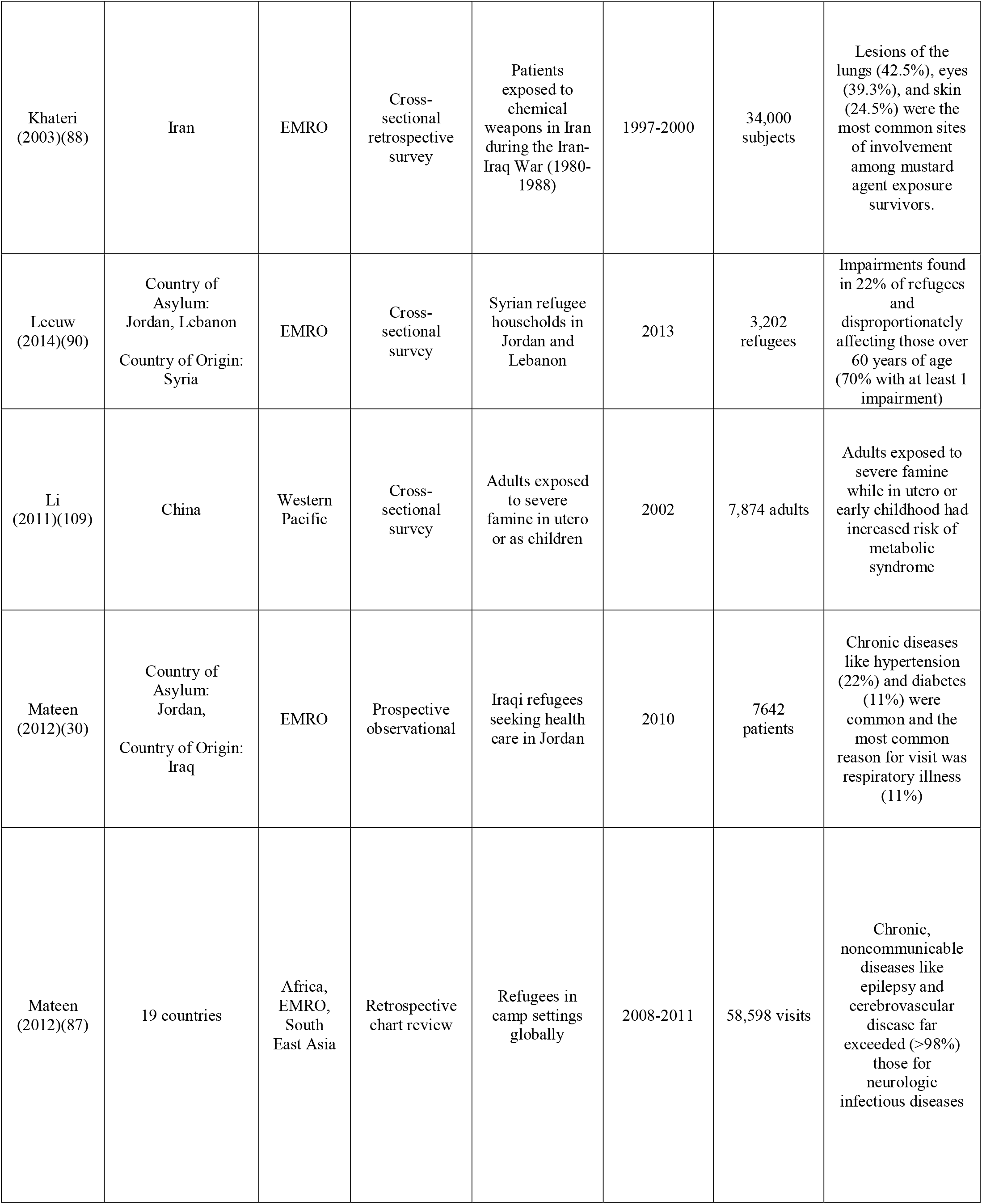
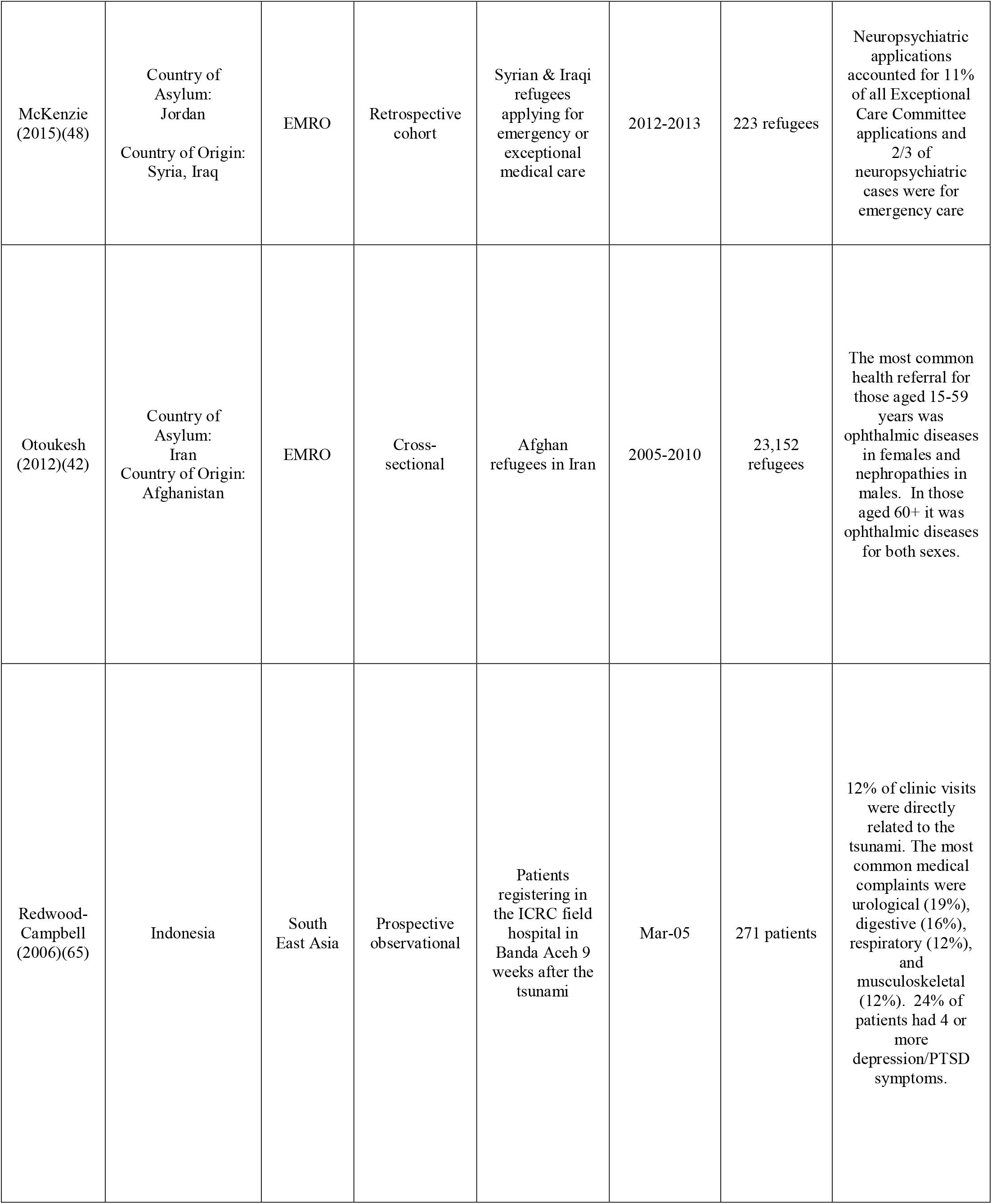
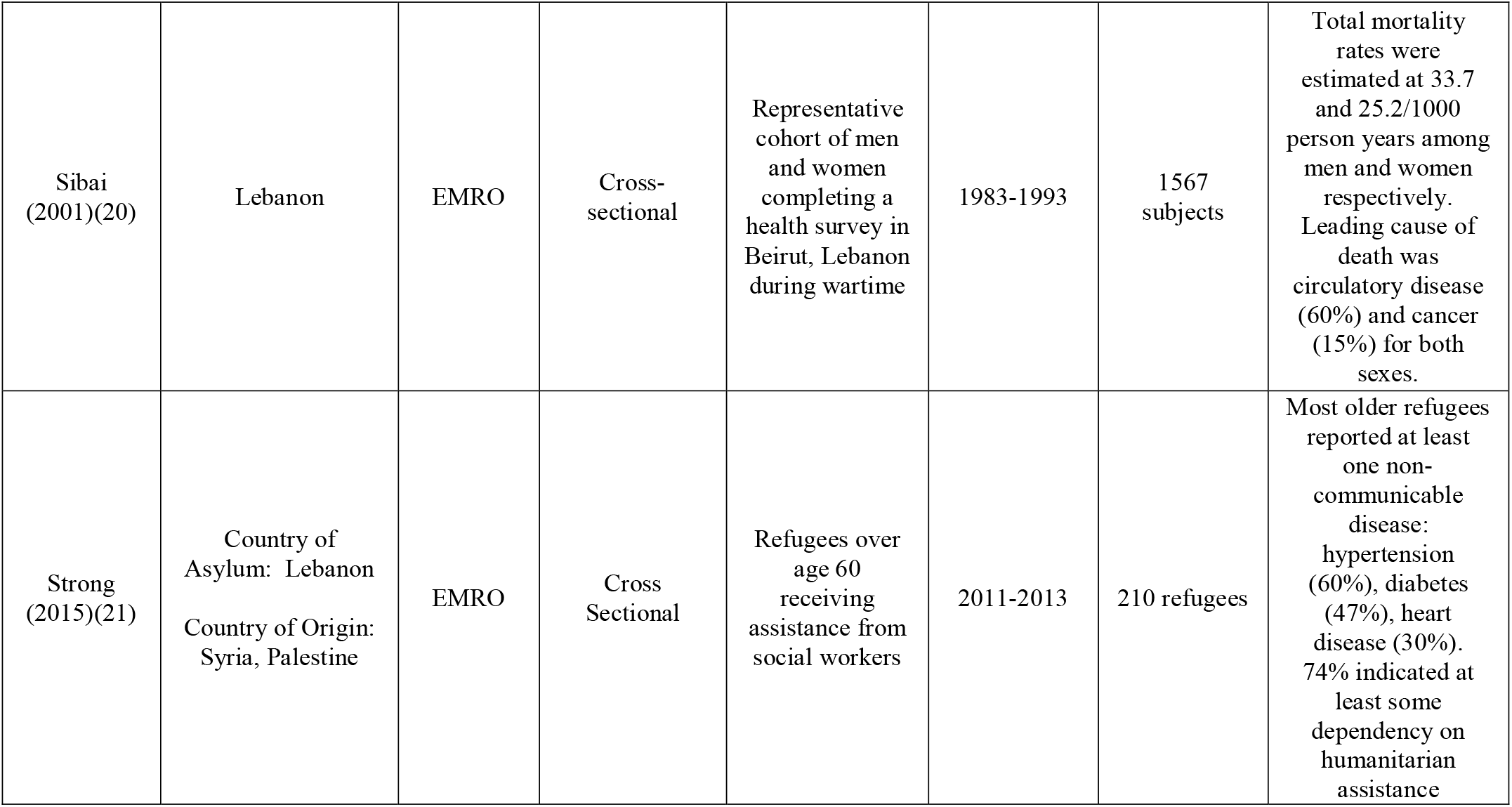
Characteristics of included publications by disease type: Other Non-Communicable Diseases

|  | Country/Territory<br>of Interest | WHO<br>region | Type of study | Target<br>Population | Years of<br>observation | Number of<br>study<br>participants | Major findings |
| --- | --- | --- | --- | --- | --- | --- | --- |
| Amini<br>(2010)(89) | Iran | EMRO | Cross-sectional | Iranian war victims blinded in both eyes | 2007 | 250 conference attendees | Quality of Life (QOL) scores in blind war victims decreased with increasing age and additional medical comorbidities |
| Armenian<br>(1998)(34) | Armenia | European | Prospective, nested case-control | Survivors of the 1988 Earthquake in Armenia | 1988-1992 | 35,043 employees of the Armenian Ministry of Health and their immediate families | During a 4-year follow-up period, the highest number of deaths from all causes (including heart disease) occurred within the first 6 months following the earthquake; associated with extent of disaster-related damage and losses |
| Chan<br>(2010)(91) | Pakistan | EMRO | Cross sectional | Face-to-face, household-based survey conducted 4 months after the 2005 Kashmir, Pakistan earthquake in internally displaced camps near Muzafarabad city | February 2006<br>4 months post-earthquake | 167 households | Although the proportion of the population with chronic conditions was similar across these studied camps, 85% of residents in the smallest unofficial camp had no available drugs to manage their chronic medical conditions as compared with their counterparts residing in larger rural unofficial (40%) and official camps (25%). |
| Hung<br>(2013)(26) | China | Western Pacific | Cross-sectional chart review | Patients presenting during a 19 day period three weeks following the Sichuan earthquake | Jun-08 | 2,034 patients | Musculoskeletal, respiratory, and GI systems were top 3 problems and >43% of patients met hypertension criteria |
| Khateri<br>(2003)(88) | Iran | EMRO | Cross-sectional retrospective survey | Patients exposed to chemical weapons in Iran during the Iran-Iraq War (1980-1988) | 1997-2000 | 34,000 subjects | Lesions of the lungs (42.5%), eyes (39.3%), and skin (24.5%) were the most common sites of involvement among mustard agent exposure survivors. |
| Leeuw<br>(2014)(90) | Country of Asylum:<br>Jordan, Lebanon<br>Country of Origin:<br>Syria | EMRO | Cross-sectional survey | Syrian refugee households in Jordan and Lebanon | 2013 | 3,202 refugees | Impairments found in 22% of refugees and disproportionately affecting those over 60 years of age (70% with at least 1 impairment) |
| Li<br>(2011)(109) | China | Western Pacific | Cross-sectional survey | Adults exposed to severe famine in utero or as children | 2002 | 7,874 adults | Adults exposed to severe famine while in utero or early childhood had increased risk of metabolic syndrome |
| Mateen<br>(2012)(30) | Country of Asylum:<br>Jordan,<br>Country of Origin:<br>Iraq | EMRO | Prospective observational | Iraqi refugees seeking health care in Jordan | 2010 | 7642 patients | Chronic diseases like hypertension (22%) and diabetes (11%) were common and the most common reason for visit was respiratory illness (11%) |
| Mateen<br>(2012)(87) | 19 countries | Africa, EMRO, South East Asia | Retrospective chart review | Refugees in camp settings globally | 2008-2011 | 58,598 visits | Chronic, noncommunicable diseases like epilepsy and cerebrovascular disease far exceeded (>98%) those for neurologic infectious diseases |
| McKenzie<br>(2015)(48) | Country of<br>Asylum:<br>Jordan<br>Country of Origin:<br>Syria, Iraq | EMRO | Retrospective<br>cohort | Syrian & Iraqi<br>refugees<br>applying for<br>emergency or<br>exceptional<br>medical care | 2012-2013 | 223 refugees | Neuropsychiatric<br>applications<br>accounted for 11%<br>of all Exceptional<br>Care Committee<br>applications and<br>2/3 of<br>neuropsychiatric<br>cases were for<br>emergency care |
| Otoukesh<br>(2012)(42) | Country of<br>Asylum:<br>Iran<br>Country of Origin:<br>Afghanistan | EMRO | Cross-sectional | Afghan<br>refugees in Iran | 2005-2010 | 23,152<br>refugees | The most common<br>health referral for<br>those aged 15-59<br>years was<br>ophthalmic diseases<br>in females and<br>nephropathies in<br>males. In those<br>aged 60+ it was<br>ophthalmic diseases<br>for both sexes. |
| Redwood-Campbell<br>(2006)(65) | Indonesia | South<br>East Asia | Prospective<br>observational | Patients<br>registering in<br>the ICRC field<br>hospital in<br>Banda Aceh 9<br>weeks after the<br>tsunami | Mar-05 | 271 patients | 12% of clinic visits<br>were directly<br>related to the<br>tsunami. The most<br>common medical<br>complaints were<br>urological (19%),<br>digestive (16%),<br>respiratory (12%),<br>and<br>musculoskeletal<br>(12%). 24% of<br>patients had 4 or<br>more<br>depression/PTSD<br>symptoms. |
| Sibai<br>(2001)(20) | Lebanon | EMRO | Cross-sectional | Representative cohort of men and women completing a health survey in Beirut, Lebanon during wartime | 1983-1993 | 1567 subjects | Total mortality rates were estimated at 33.7 and 25.2/1000 person years among men and women respectively. Leading cause of death was circulatory disease (60%) and cancer (15%) for both sexes. |
| Strong<br>(2015)(21) | Country of Asylum: Lebanon<br>Country of Origin: Syria, Palestine | EMRO | Cross Sectional | Refugees over age 60 receiving assistance from social workers | 2011-2013 | 210 refugees | Most older refugees reported at least one non-communicable disease: hypertension (60%), diabetes (47%), heart disease (30%). 74% indicated at least some dependency on humanitarian assistance |

Hung et al described musculoskeletal complaints constituting 30.4% of presentations among those visiting a Hong Kong Red Cross clinic in rural China following the 2008 Sichuan earthquake (26). Mateen et al. conducted a far-reaching study of refugees in 127 camp settings across 19 countries and found that reportable neurologic diseases accounted for 59,598 visits over a 4-year period (87). Nearly 90% of these cases were for epilepsy, which they highlight far outweighed the prevalence of neurological diagnoses of an infectious nature. Another study investigated neuropsychiatric disorders among Syrian and Iraqi refugees in Jordan via retrospective review of applications to the Jordanian Exceptional Care Committee, and found stroke to be the most common neuropsychiatric diagnosis (n = 41 applications, 16% of neuropsychiatric applications; median age 64 years) (48).

Specific ophthalmic diseases identified by Mateen et al include cataracts (1.44 visits per refugee) and glaucoma (1.46 visits per refugee), which were exceeded only by cerebrovascular disease (1.46 visits per refugee) among Iraqi refugees in Jordan (30). Of note, more than half of the refugees received concomitant diagnoses in one visit. Otoukesh describes ophthalmic disease as the most common health referral (13.65%) for those aged 15-59 among Afghan refugees in Iran(42). Amini et al(89) measured Quality of Life (QOL) scores in Iranian survivors totally blinded during the Iran-Iraq War, the effects of which were mitigated among those with higher levels of education (p=0.006). Urologic complaints were identified as predominant in the ICRC hospital in Banda Aceh, Indonesia with 19% of complaints(65); specific examples of urologic disorders from Mateen et al among Iraqi refugees constituting a significant amount of morbidity were prostatic hypertrophy and nephrolithiasis(30). Hematologic disorders were described by Otoukesh, and the type of disorder varied by ethnicity, with referrals for the Baluch being the highest at 25% (42).

### Concomitant affliction with NCDs

Finally, co-affliction with multiple NCDs was a recurrent issue in our findings. This was demonstrated by Strong et al among Palestinian refugees in Lebanon, with an average of 4 NCDs per person; Syrian refugees in the same study had an average of 2.5 NCDs per person (21). Three or more risk factors were also seen in displaced persons in Croatia, a statistically significant difference in prevalence when compared to age-matched controls who were not displaced (25). Clustering of risk factors was also evident in a populations being served by UNRWA in Jordan, Syria, Lebanon, West Bank, and the Gaza Strip, and the risk of having CVD was 2.7 times higher in individuals with 4 risk factors as compared to those with only 1 risk factor (29). Concomitant affliction also conferred worse outcomes among Palestine refugees in Jordan with CVD (myocardial infarction, congestive heart failure, stroke and blindness) among those with hypertension and diabetes, when compared to those with hypertension alone in the same cohort (p<0.01) (41). Yusef et al also demonstrated that having concomitant risk factors (such as diabetes and hypertension) resulted in a higher likelihood of presentation with late complications of NCDs at a UNRWA primary healthcare field site in Lebanon (22).

## Discussion

NCDs represented a significant burden for populations affected by humanitarian crises and natural disasters for all regions (20, 39, 42, 44, 45, 48, 49, 53, 67, 92-94, 124), and even conferred increased mortality and morbidity when compared to infectious diseases in one study (95). Individuals with NCDs may present for continued management of chronic conditions or with an acute exacerbation of a chronic NCD triggered by the acute disaster state (64, 65). Furthermore, several studies demonstrated that NCDs adversely affected morbidity of populations in humanitarian crises with women and older populations disproportionately affected (60, 71, 89, 96). Late stage complications of cardiovascular diseases and diabetes including stroke (24, 48, 78, 87, 95), diabetic foot amputations (22, 46, 78, 94), and myocardial infarctions (31, 39) were described in all regions (22, 78). However, pulmonary diseases such as asthma, COPD and lung cancer were noticeably lacking. Similarly, scant studies focused on pediatric populations (59, 63, 66, 67). Strengthening and broadening the spectrum of NCD diagnoses included in disaster management planning is key, and particular focus on children and adolescents is critical as these age groups present key opportunities for interventions to mitigate future NCD morbidity (97, 98). Finally, several studies identified challenges and epidemiologic factors specific to refugee populations or subgroups of refugee populations (42, 45, 51, 53, 55), and they also highlight that refugee populations are heterogenous in their disease burden. Understanding these populations is key in guiding the medical equipment, personnel including specialists, and potential screening programs that should be considered in future humanitarian efforts.

### Disparities in evidence by region

There is little information (19, 44, 94) for all regions and disease categories about the experience of the affected population during acute crises, active conflict, or as internally displaced persons (IDPs) (42, 45, 48, 53, 55) (see Tables 8-10). As far as regional focus of studies assessed, both Sub-Saharan Africa and the Americas were poorly represented in the literature on NCDs in humanitarian crises (40, 44, 75, 95) (see Tables 6-7). This is in spite of the fact that these regions experience a marked dual burden of armed conflict (99, 100) and natural disasters (101, 102), and represent a significant portion of the global NCD burden (103, 104). 32.4%

**Table 6:** Characteristics of included publications by region: Africa

|  | Country/Territory of Interest | Target Population | Type of Study | NCD Studied | Years of Observation | Number of study participants | Major Findings |
| --- | --- | --- | --- | --- | --- | --- | --- |
| Besancon<br>(2015)(44) | Mali | Mali diabetic population following a March 2012 Coup in Bamako | Case Study | Diabetes | Spring 2012 following the March 2012 coup | -- | Diabetics are a vulnerable population in humanitarian crisis due to their continuous need for health care and medicines and the financial burden this may place on them. In an emergency setting sub-populations of diabetics must be taken into account for humanitarian response planning; including people still in active conflict regions, IDPs, refugees, and the host population which houses IDPs. |
| Habtu<br>(1999)(75) | Ethiopia | Insulin treated diabetic patients from the Diabetic Clinic at the Mekelle Hospital in rural Tigray, Northern Ethiopia- the center of the severe Ethiopian famine of the mid-1980s | Cross-sectional | Diabetes | Six month period in 1997 | 100 patients | The correct prescribed dose of insulin was only being administered in 50% of DM patients in rural Tigray, Ethiopia and the correct syringe by only 12% of patients. Insulin treatment had been interrupted in 48% of cases due to lack of supply. Low BMI(mean of 15.8), young age, and resistance to diabetic ketoacidosis(DKA) amongst study participants were consistent with previous descriptions of malnutrition related diabetes mellitus(MRDM). |
| Huerga<br>(2009)(95) | Liberia | Patients of the medical and pediatric wards of Mamba Point Hospital, Monrovia, Liberia, one year after the end of the Liberian civil war | Cross-sectional | Multiple NCDs including CVD (stroke, CHF, and HTN) | January 2005 - July 2005 | 1,034 adult patients<br>1,509 children | Of 1034 adult hospitalized patients in post-war Liberia, 529 (51%) were diagnosed with a noninfectious disease. Among the 241 deaths recorded, the cause was non-infectious disease in 134 (56%) patients. The fatality rate for infectious diseases (19.7%; 92 deaths/465 cases) was lower ($P = 0.04$) than for non-infectious diseases (25.3%; 134 deaths/529 cases). Cardiovascular diseases caused half of deaths due to non-infectious diseases: 25% stroke, 18% heart failure and 10% severe hypertension. No cases of ischemic heart disease were identified.<br>-----<br>Among hospitalized children, 229 (15%) were diagnosed with a noninfectious disease. NCDs represented 34% of all deaths. The fatality rate for infectious diseases (18.6%; 197 deaths/1189 cases) was lower ($P < 0.01$) than for non-infectious diseases (28.8%; 66 deaths/229 cases). |
| Hult<br>(2010)(40) | Nigeria | 40 year old Nigerians with fetal exposure to famine in Biafra, Nigeria during the Nigerian civil war (1967–1970) | Retrospective cohort | Diabetes and HTN | June 2009– July 2009 | 1,339 study participants | Fetal and infant undernutrition was associated with significantly increased risk of hypertension(adjusted OR 2.87; 95% CI 1.90-4.34), and impaired glucose tolerance (OR 1.65; 95% CI 1.02-2.69) in 40 year old Nigerians. However, early childhood exposure was not associated with increased risk. |

**Table 7:** Characteristics of included publications by region: Region of the Americas

|  | Country/Territory of Interest | Target Population | Type of Study | NCD Studied | Years of Observation | Number of study participants | Major Findings |
| --- | --- | --- | --- | --- | --- | --- | --- |
| Daniels (2009)(92) | Peru | Displaced as well as nondisplaced populations in rural to urban settings | Stratified cluster survey | Multiple NCDs | 2007 (six months post-earthquake) | 672 households | Displaced populations sought care more, people with injury or NCD sought care more. People who did not seek care cited cost as a barrier. |
| Naumova(2007)(63) | Ecuador | Pediatric ER visits with acute upper and lower respiratory conditions and asthma related conditions | Retrospective review of medical records | Chronic respiratory disease including asthma | January – 2000<br>December 2000 | 5,169 emergency department records | Rates of ED visits for pediatric patients increased significantly during period of volcanic activity. Youngest patients (4 and under) were most affected. |

**Table 8:** Characteristics of included publications by region: Eastern Mediterranean Region

|  | Country/ Territory of Interest | Target Population | Type of study | NCD studied | Years of observation | Number of study participants | Major findings |
| --- | --- | --- | --- | --- | --- | --- | --- |
| Abukhdeir (2013)(32) | Palestinian Territories- Gaza/ West Bank | Palestinian households in the West Bank and Gaza Strip | Cross-sectional nationally representative household survey | Diabetes, hypertension, cardiovascular disease (CVD) and cancer | 2013 | 4,456 households in the West Bank and 2118 in the Gaza Strip. The response rates for the 2 regions were 84.1% and 96.9% respectively | The authors emphasized that even though previous studies have combined Palestinians as one group, they live in different areas and are subject to different health systems which can result in different health outcomes. Being a refugee was a significant risk factor for diabetes and CVD while being married/engaged or divorced/ separated widowed was a risk factor for diabetes and hypertension. Non-refugees were 33% less likely to have diabetes and 46% less likely to have CVD than refugees. |
| Abul<br>(2001)(57) | Kuwait | Patients admitted to hospitals in Kuwait with asthma for six years (1987-1989 and 1992-1994) | Retrospective cross-sectional study | Asthma | 2001 | 12,113 asthma patients during the pre-Gulf War period compared with 9,771 patients during the post-Gulf War period | During the war, a lot of oil wells were burned, giving suspicion to the potential for increase in asthma. No statistically significant difference in hospital admissions for to death rates attributable to asthma in the pre- and post-Gulf War periods in Kuwait. Notably, the war was 1990/1991, and no data is available for those years, so the immediate effect isn't known. |
| Ahmad<br>(2015)(85) | Syria | Syrian national health system | Situational analysis using document analysis, key informant interviews, and direct clinic observation | Diabetes and cardiovascular disease (CVD) | October 2009 - August 2010 | 53 semi-structured interviews | The rebuilding of a post-conflict health care system in Syria may benefit from insights into the structural problems of the pre-crisis system. Weaknesses that existed before the crisis are compounded by the current conflict. The authors suggest an over reliance on secondary and tertiary care for DM patients with withdrawal of the Syrian government from the public health clinics, which led to escalating healthcare costs and fostered increasingly unequal access. |
| Alabed<br>(2014)(105) | Country of Asylum: Syria<br>Country of Origin: Palestinian Territories | Palestinian refugees living in Damascus attending three UNRWA health clinics | Cross sectional | Diabetes | August 2008 - September 2008 | 154 DM patients | UNRWA clinic inspections highlighted shortages in drug stocks with 47.3% of patients reporting problems accessing prescribed medications and 67.7% reporting having to buy medications at their own expense at least once since their diagnosis. Patients' knowledge of their condition was limited, Patients were generally unaware of the importance of good glucose control and disease management. Women were more likely to attend the clinic than men, with 71% of patients being female. |
| Ali-Shtayeh (2012)(82) | Palestinian Territories-West Bank | Patients attending outpatient departments at Governmental Hospitals in 7 towns in the Palestinian territories (Jenin, Nablus, Tulkarm, Qalqilia, Tubas, Ramalla, and Hebron) | Cross-sectional survey | Diabetes | August 2010 - May 2011 | 1,883 DM patients | The use of CAM differed significantly between residents of refugee camps versus residents of urban or rural areas ($p = 0.034$). Those who were on CAM reported they were using it to slow down the progression of the disease or relieve symptoms. All patients with DM who used CAM were also on conventional therapies. |
| AlKasseh (2014)(84) | Palestinian Territories-Gaza | Patients at UNRWA clinics within Gaza | Retrospective case-control study | Gestational diabetes (GDM) | March 2011 - June 2011 | 189 postnatal GDM women with 189 matched controls by age and place of residency | The present study showed that history of miscarriage more than once, being overweight before pregnancy, history of stillbirth, history of caesarean birth and positive family history of diabetes mellitus were strongly correlated with developing GDM. The WHO criteria for screening for GDM remains a good instrument to identify GDM in refugee populations in war-torn countries (like the Gaza Strip). |
| Amini (2010)(89) | Iran | Completely blind Iranian survivors of the Iran-Iraq War. | Cross-sectional study | Multiple NCDs including hypertension, Hypercholesterolemia, and erectile dysfunction | 2010 | 250 Iran-Iraq war survivors | As blind war survivors' age, they will present with a greater set of burdens despite their relatively better quality of life (QOL) in the physical component scale when compared with lower limb amputees. Risk factors of cardiovascular attack such as high blood pressure and hypercholesterolemia were present: High systolic and diastolic blood pressure, hearing loss, and tinnitus had negative individual correlations to (QOL) ( $p = 0.016$ , $0.016$ , $0.005$ , $p < 0.0001$ ). Hypercholesterolemia showed significant correlation to QOL ( $p = 0.021$ ). |
| Bijani (2002)(58) | Iran | Iranians injured by chemical weapons during the Iraq–Iran war who are under services of the Mostazafan and Janbazan Foundations of Babol, Iran | Cross-sectional | Chronic respiratory diseases | 1994 - 1998 | 220 patients | The clinical evaluations, radiography, and PFTs revealed that the most prevalent effects of chemical weapons on respiratory tract were chronic obstructive lung disease. Victims of sulphorous gas had demonstrated involvement of airways during acute and chronic phases of injury, however over time clinical manifestations, radiography, and PFT gradually became normal. Most patients reported mustard gas exposure.. Chest X-Ray was not reliable to diagnose lung injury in these patients. Diagnosis was completed most accurately by PFTs. |
| Ben Romdhane (2015)(86) | Tunisia | Tunisian national health system | Situational analysis | Cardiovascular disease and diabetes | 2010 | 12 key informants were interviewed and eight documents were reviewed | Weaknesses that existed before the 2011 Revolution(Arab Spring) were compounded during the revolution. This study was conducted prior to political conflict but written post-conflict. Growth of the private sector fostered unequal access by socioeconomic status and reduced coordination and preparedness of the health system. |
| Chan (2009)(124) | Pakistan | Patients $\geq 45$ years who attended two different types of post- | Comparative descriptive study | Multiple NCDs | February 2006<br>4 months post-earthquake | 30,000 patients in a rural site, and 382 | The greatest gap in health services post-earthquake in both sites was non-communicable |
|  |  | earthquake relief clinics during a 17-day field health needs assessment in response to the 2005 Kashmir earthquake |  |  |  | IDPs in a urban site | disease management. Clinical records reviewed in all study locations showed a systematic absence of documentation of common NCDs. In rural areas, older women were less likely to receive medical services while older men were less likely to access psychological services in both sites. During days when solely male doctors provided clinical services in the rural site, medical services utilization decreased by 30%. |
| Chan (2010)(91) | Pakistan | Face-to-face, household-based survey conducted 4 months after the 2005 Kashmir, Pakistan earthquake in internally displaced camps near Muzafarabad city | Cross sectional | Multiple NCDs | February 2006<br>4 months post-earthquake | 167 households | Although the proportion of the population with chronic conditions was similar across these studied camps, 85% of residents in the smallest unofficial camp had no available drugs to manage their chronic medical conditions as compared with their counterparts residing in larger rural unofficial (40%) and official camps (25%). |
| Doocy (2013)(115) | Country of Asylum: Jordan/<br>Syria<br>Country of Origin: Iraq | Iraqi populations displaced in Jordan and Syria | Cross-sectional | Disability and multiple NCDs including hypertension, arthritis, diabetes, chronic respiratory diseases, and cardiovascular disease | October 2008-March 2009 | 1200 and 813 Iraqi households in Jordan and Syria, respectively | Chronic disease prevalence among adults was 51.5% in Syria and 41.0% in Jordan, with hypertension and musculoskeletal problems most common. Overall disability rates were 7.1% in Syria and 3.4% in Jordan, with the majority of disability attributed to conflict and depression the leading cause of mental health disability. |
| Doocy (2015)(114) | Country of Asylum: Jordan<br>Country of Origin: Syria | Syrian refugees in non-camp settings in Jordan | Cross-sectional survey | Multiple NCDs including hypertension, arthritis, diabetes, chronic respiratory diseases, and cardiovascular disease | 1994 - 1998 | 1,550 refugees | More than half of Syrian refugee households in Jordan reported a member with an NCD. Among adults, hypertension prevalence was the highest (9.7 %, CI: 8.8–10.6). While care-seeking was high (85%) among those reporting a NCD, among those who did not seek care, cost was the primary reason. |
| Ebrahimi (2014)(37) | Iran | Patients with cardiovascular and respiratory diseases who received medical services from the Center for Disaster and Emergency Medicine in Sanandaj, Iran during dust event days | Ecological study | Cardiovascular and respiratory diseases | March 2009 - June 2010 | -- | The authors demonstrated a statistically significant increase in emergency admissions for cardiovascular diseases during dust storm episodes in Sanandaj, Iran (r 0.48, p<0.05). The correlation between respiratory diseases and dust storm events were statistically insignificant (0.19). |
| Eljedi (2006)(71) | Palestinian Territories- Gaza | Diabetic patients who were recruited from three refugee camps in the Gaza strip with age- and sex- matched controls living in the same camps | Cross sectional | Diabetes | November 2003 - December 2004 | 197 patients | Using the World Health Organization Quality of Life questionnaire (WHOQOL-BREF) four domains were strongly reduced in diabetic patients as compared to controls, with stronger effects in physical health (36.7 vs. 75.9 points of the 0–100 score) and psychological domains (34.8 vs. 70.0) and weaker effects in social relationships (52.4 vs. 71.4) and environment domains (23.4 vs. 36.2). The impact of diabetes on health-related quality of life was especially severe among females and older subjects. |
| El-Sharif (2002)(66) | Palestinian Territories- West Bank | Schoolchildren aged 6–12 years attending 12 schools in the Ramallah District of the Palestinian West Bank | Cross-sectional | Asthma | Autumn of 2000 | 3,382 children | Children from refugee camps were at a higher risk of asthma and asthma symptoms than children from neighboring villages or cities. Physician-diagnosed asthma was almost double in refugee camps than other places (15.6% versus 8.1% in villages and 7.3% in cities, pv0.001). |
| Forouzan<br>(2014)(61) | Iran | Patients<br>presenting<br>with asthma<br>or<br>bronchospasm<br>in western<br>Iran | Prospective<br>observational | Asthma | November<br>2013 | 2,000<br>patients | Many patients<br>presented with<br>bronchospasm after<br>a thunderstorm. |
| Kallab<br>(2015)(19) | Country of<br>Asylum:<br>Lebanon<br>Country of<br>Origin:<br>Syria | Syrian<br>refugees and<br>vulnerable<br>Lebanese<br>being treated<br>in 8 health<br>facilities run<br>by Amel<br>Association<br>International | Program<br>implementatio<br>n reflection | Diabetes and<br>hypertension | November<br>2014- May<br>2015 | 1,825<br>patients | Of the 1,825 patients<br>enrolled in the<br>program<br>hypertension and<br>diabetes accounted<br>for 46% and 27% of<br>cases respectively,<br>with the remaining<br>27% of patients<br>presenting with both<br>diseases. The<br>program addressed<br>two main problems<br>in Lebanon: lack of<br>access to NCD<br>services and lack of<br>proper management<br>of NCDs. Major<br>challenges included<br>insecurity in the<br>country, patient<br>transportation cost,<br>and high workload<br>for providers. |
| Karrouri<br>(2014)(69) | Country of<br>Asylum:<br>Tunisia<br>Country of<br>Origin:<br>Libya | Case of a 10-year-old<br>Libyan boy | Case report | Diabetes | -- | 1 patient | Report of a 10 year<br>old without personal<br>or familial diabetes<br>mellitus history who<br>developed type 1<br>diabetes appeared<br>immediately<br>following severe<br>psychological<br>trauma. |
| Khader<br>(2012)(81) | Country of<br>Asylum:<br>Jordan<br>Country of<br>Origin:<br>Palestinian<br>Territories | Persons with<br>DM at Nuzha<br>PHC Clinic | Retrospective<br>descriptive<br>study of the<br>cohort<br>reporting<br>framework to<br>monitor<br>burden of<br>disease and<br>management | Diabetes | October<br>2009-March<br>2012 | 2851<br>patients | A directly observed<br>therapy (DOTS)<br>cohort monitoring<br>system can be<br>successfully adapted<br>and used to monitor<br>and report on<br>Palestinian refugees<br>with DM in Jordan.<br>A sizeable<br>proportion of DM<br>patients of the clinic<br>failed to have<br>postprandial blood<br>glucose<br>measurements, and<br>BP measurements in<br>those with comorbid<br>HTN. The study<br>demonstrated to the<br>clinic that they were<br>either not<br>performing or not<br>recording disease-<br>specific procedures<br>that should be done<br>at the investigated<br>visits - can now<br>improve on these in the future and monitor thanks to e-Health system |
| Khader (2013)(78) | Country of Asylum: Jordan<br>Country of Origin: Palestinian Territories | Palestine refugees living in Jordan | Descriptive cohort study using routine data collected through e-Health | Diabetes | October 2009- June 2013 | 12,549 total patients | High burden of disease with predicted annual additional caseload is over 1,000 patients with DM. Many indicated risk factors: smoking, physically inactive, and obesity. Those who came had relatively good disease control.<br>Points to the importance of using e-Health systems to monitor and evaluate and use for strategic planning.<br>Complications, including myocardial infarction and end-stage renal disease were significantly more common in males. Females were more likely to be obese. |
| Khader (2014)(41) | Country of Asylum: Jordan<br>Country of Origin: Palestinian Territories | Palestine refugees living in Jordan | Retrospective cohort study with program and outcome data collected and analyzed using E-Health | Hypertension | October 2009- June 2013 | 18,881 patients | Endorses the use of E-Health and cohort analysis for monitoring and managing patients with HTN and DM. High case load from HTN and comorbid HTN and DM(40-50%) amongst Palestinian refugees being treated at UNRWA primary health care clinics in Jordan. Most common risk factors included smoking, physical inactivity, and obesity. 33% of males smoked, while more than 50% of the women were physically inactive. 75% of women were obese. |
| Khader (2014)(80) | Country of Asylum: Jordan<br>Country of Origin: Palestinian Territories | Palestinian refugees living in Jordan with DM attending Nunzha Clinic | Retrospective cohort | Diabetes | 2010-2013 | 119 DM patients | The E-health system was successful in monitoring annual outcomes, measures of disease control, and development of complications in a cohort of patients with DM. Three major findings were: a progressive loss of<br>patients attending the clinic, mainly lost to follow-up; routine measurements were always performed, and there was a progressive increase in late-stage complications, predominately due to cardiovascular disease and stroke. |
| Khader (2014)(79) | Country of Asylum: Jordan<br>Country of Origin: Palestinian Territories | Palestinian refugees living in Jordan with DM attending Nunzha Clinic | Retrospective cohort study | Diabetes | 2012 | 2,974 DM patients | E-Health systems are useful for monitoring patients, since over half of patients who fail to attend a scheduled quarterly appointment are declared lost to follow-up 1 year later. This suggests a need for monitoring and active follow-up |
| Khan (1997)(55) | Country of Asylum: Pakistan<br>Country of Origin: Afghanistan | Patients from North West Pakistan and Afghan refugees attending the Institute of Radiotherapy and Nuclear Medicine, Peshwar | Cross-sectional | Cancer | 1990 - 1994 | 13,359 patients | In male Afghan refugees, esophageal cancer represented 16.6% of the cases, compared to only 4.6% of the cases in Pakistani residents. Similar patterns in women (13.1% vs. 4.1%) |
| Khateri (2003)(88) | Iran | Individuals with confirmed exposure to mustard agent during the Iran-Iraq war of 1980-1988 and who were evaluated for exposure to mustard agent by medical authorities | Retrospective Cohort | Chronic pulmonary, ocular, and cutaneous lesions | 1997-2000 | 34,000 cases | Among patients, there was a high degree of pulmonary disease: 42.5% of the exposed population exhibiting chronic lung lesions and associated symptoms. Ocular damage, which is observed to be present in 39.3% of mustard exposed Iranians, is another major consequence of exposure to these agents as a result of their ease of absorption through the unprotected eye. |
| Lari (2014)(60) | Iran | Patients exposed to sulfur mustard gas | Cross sectional | Chronic obstructive pulmonary disease (COPD) | March 2010 - April 2011 | 82 patients | The COPD Assessment Test (CAT) was found to be a valid tool for assessment of health-related quality of life in chemical warfare patients with COPD. |
| Leeuw (2014)(90) | Country of Asylum: Jordan, | Syrian refugee households in Jordan and | Cross sectional | Multiple NCDs | 2013 | 3,202 refugees | Impairments found in 22% of refugees and |
|  | Lebanon<br>Country of Origin: Syria | Lebanon |  |  |  |  | disproportionately affecting those over 60 years of age (70% with at least 1 impairment) |
| Mansour (2008)(76) | Iraq | Patients struggling with diabetic control | Cross sectional | Diabetes | January 2007-December 2007 | 3,522 patients | Patient opinion for not achieving good glycemic control included the following: 50.8% cases reported no drug supply or drug shortage, while 50.2% reported high drugs and/or laboratory expenses. 30.7% percent of patients said that they were unaware of diabetic complications and 20.9% think that diabetes is an untreatable disease. 30% think that non-control of their diabetes is due to migration after the war. No electricity or erratic electricity, self-monitoring of blood glucose is not available, or strips were not available or could not be used, and illiteracy as a cause was seen in 15%, 10.8% and 9.9% respectively. |
| Mateen (2012)(30) | Country of Asylum: Jordan<br>Country of Origin: Iraq | Iraqi refugees receiving UNHCR health assistance in Jordan | Cross sectional | Multiple NCDs including hypertension, visual disturbances, diabetes, and joint disorders | January 2010-December 2010 | 7,642 registered Iraqi refugees | Among adults 18 years or older, 22% had hypertension; 11% had type II diabetes mellitus; 4% had type I diabetes mellitus; 10% had visual disturbances; 10% had disorders of lipoprotein metabolism and other lipidemias; 9% had other joint disorders and 7% had chronic ischemic heart disease. Cancer care was required by 2% of refugees. For all refugees as a group, the largest number of visits were for essential hypertension (2067 visits); visual disturbances (1129); type II diabetes mellitus (1021); other joint disorders (969), and acute upper respiratory infections (952). |
| McKenzie (2015)(48) | Country of Asylum: Jordan<br>Country of Origin: Iraq, Syria | Iraqi/Syrian refugees residing in Jordan | Retrospective cohort | Neuro-psychiatric disorders | 2012–2013 | 223 refugees | Among neuropsychiatric applications, stroke was the most common diagnosis, accounting for 16%. Brain tumors accounted for 13% of neuropsychiatric applications and was the most expensive diagnosis overall and per applicant. The ECC denied six applications for reasons of eligibility, cost, and/or prognosis. Of the 20 approved applications, 15% (n = 3) were approved for less than the requested amount, receiving on average 39% of requested funds. |
| Mirsadraee (2011) (59) | Iran | Patients whose parents were exposed to chemical warfare | Case control | Asthma | -- | 409 children | The prevalence of asthma was not significantly different in the offspring of chemical warfare victims. |
| Mousa (2010)(29) | Country of Asylum: Jordan, Lebanon, Syria, West Bank/Gaza<br>Country of Origin: Palestinian Territories | Refugees registered by the United Nations Relief and Works Agency for Palestine Refugees in the Near East (UNRWA) | Case series | Diabetes and hypertension | June 2007 | 7,762 refugees | A total of 9% of those screened were diagnosed with hypertension or diabetes. Being older than 40 years, obese or with a positive family history of diabetes or cardiovascular disease increased the risk of presenting with hypertension and/or hyperglycemia 3.5, 1.6 and 1.2 times respectively. Risk factors were very common (obesity and smoking). |
| Otoukesh (2012)(42) | Country of Asylum: Iran<br>Country of Origin: Afghanistan | Afghan refugees in Iran | Retrospective cross sectional | Multiple NCDs including ophthalmic diseases, neoplasm, nephropathies, ischemic heart disease, and perinatal disorders | 2005 -2010 | 23,152 refugees | The Afghan refugees who received referrals for care represented a higher number of women, age 15- 59 years old, for ophthalmic diseases, neoplasms, and nephropathies. |
| Shamseddine (2004)(49) | Lebanon | Lebanese population following the 1975 -1990 Lebanese Civil War | Nationwide, Population-Based Prevalence Study | Cancer | 1998 | 4,388 cases | Among males, the most frequently reported cancer was bladder (18.5%), followed by prostate (14.2%), and lung<br>cancer (14.1%)<br>Among females, breast cancer alone constituted around one third of the total cancer caseload in the country, followed by colon cancer (5.8%), and cancer of the corpus uteri (4.8%). One limitation of the study is that the last and only census undertaken in Lebanon was in 1932, and the population estimates and projections may have been subject to minor inaccuracies. |
| Sibai (2001)(20) | Lebanon | Lebanese aged 50 years and over residing in Beirut, Lebanon in 1983 | Retrospective cohort study | Multiple NCDs including cancer, cardiovascular disease, cancer, and nephropathies. | 1983-1993 | 1,567 cases | The most important causes were non-communicable diseases, mainly circulatory disease (60%); and cancer (15%). Among circulatory diseases, ischaemic heart disease accounted for the majority of the mortality burden (68%) followed by cerebrovascular diseases (21%). In countries that lack reliable sources of mortality data, the utility of verbal autopsy can be viably extended to cohort studies for assessing causes of death. |
| Sibai (2007)(122) | Lebanon | Lebanese aged 50 years and over residing in Beirut, Lebanon | Retrospective cohort study | Cardiovascular disease | 1984-1994 | 1,567 cases | Most important causes of death were CVD and Cancer. High adjusted risk of CVD mortality associated with being single (never-married) versus married among men and women |
| Sofeh (2004)(83) | Country of Asylum: Peshawar, Pakistan<br>Country of Origin: Afghanistan | Afghan refugees attending Red Cross dispensaries and hospitals in Peshawar Pakistan | Cross-sectional | Multiple NCDs including diabetes mellitus | -- | 456 patients | Out of 456 patients examined during the study, 255 patients suffered from DM, 80 with hepatitis, 69 with nephritis, and 52 with hyperlipidemia. |
| Strong (2015)(21) | Country of Asylum: Lebanon | Syrian refugees over age 60 | Cross-sectional | Multiple NCDs including hypertension, | March 2011 - March | 210 refugees | Older refugees reported a high burden of chronic |
|  | Country of Origin: Syria | residing in Lebanon and registered with either Caritas Lebanon Migrant Center (CLMC) or the Palestinian Women's Humanitarian Organization (PALWHO) |  | diabetes, heart disease, hyperlipidemia, arthritis, and ocular diseases. | 2013 |  | illnesses and disabilities. Hypertension was most common (60%), followed by diabetes mellitus (47%), and heart disease (30%). The burden from these diseases was significantly higher in older Palestinians compared to older Syrians, even when controlling for the effects of sex and age. Financial difficulties were given as the primary reason for not seeking care by 79% of older refugees. |
| Wright (2010)(56) | Kuwait | Kuwaiti nationals ages 50-69 exposed to the 1990 Iraqi invasion | Cross-sectional | Asthma and PTSD | December 2003 - January 2005. | 5,028 subjects | War-related stressors were associated with elevated risk of incident asthma in elderly Kuwaiti civilians exposed to 1990 Iraqi invasion. Study suggested that those who reported highest stress exposure in the invasion were more than twice as likely to report asthma. Suggestive of correlation between war trauma and asthma. |
| Yaghi (2012)(94) | Lebanon | Cases of amputations in Lebanon | Cross-sectional | Diabetes | January 2007 - December 2007 | 661 amputations | Diabetes and vascular indications were not only more common than trauma-related amputation, but both were associated with more major surgery and longer hospital stay including conflict afflicted southern Lebanon where trauma, diabetes and vascular disease amputations all occurred at more than twice the national rate. |
| Yusef (2000)(22) | Country of Asylum: Lebanon<br>Country of Origin: Palestinian Territories | Diabetic and hypertensive patients attending UNRWA primary health care facilities in Lebanon | Cross-sectional | Diabetes and hypertension | 1997 | 2,202 records | Presence of both diabetes and hypertension increased the risk for late-stage complications. Only 18.2% of diabetic patients and 17.7% of diabetic patients with hypertension were managed by lifestyle modification. About 50% of type 2 and 66% of type 1 patients who were on insulin were well controlled. Medication shortages may drive medication choices for hypertension. |
| Zubaid (2006)(39) | Kuwait | Catchment area of Mubarak Al Kabeer Hospital | Ecological | Acute myocardial infarction (AMI) | March 2003 | 1 Missile Attack Period (MAP) and 4 control periods | The number of admissions for AMI was highest during MAP, 21 cases compared to 14-16 cases in the four control periods, with a trend towards increase during MAP (incidence rate ratio=1.59; 95% CI 0.95 to 2.66, p<0.07). The number of admissions for AMI during the first 5 days of MAP was significantly higher compared to the first 5 days of the four control periods (incidence rate ratio=2.43; 95% CI 1.23 to 4.26, p<0.01). This indicates missile attacks were associated with an increase in the incidence of AMI. |

**Table 9:** Characteristics of included publications by region: Western Pacific

|  | Country/<br>Territory<br>of Interest | Target<br>Population | Type of<br>Study | NCD<br>Studied | Years of<br>Observation | Number of<br>study<br>participants | Major Findings |
| --- | --- | --- | --- | --- | --- | --- | --- |
| An (2014)(68) | China | Adults who had lived through the 1976 Tangshan earthquake (vs control Age 37-60) | Cross-sectional | Diabetes | Sept 2013 – Dec 2013 | 1551 adults | Earthquake stress linked to higher incidence of DM<br>Women more likely to have diabetes after experiencing earthquake stressors compared to men. |
| Chan (2011)(125) | China | Evacuees from the 2008 Sichuan earthquake | Descriptive, cross sectional study | Multiple NCDs including diabetes, hypertension, heart failure | May 2008 | 132 adults | Chronic health needs constituted a significant proportion of emergency care during the acute phase of the earthquake. Disaster responders must consider NCDs as well as trauma. |
| Chen (2009)(35) | China | Adults who were in the West China Hospital on the day of the 2008 Sichuan (Wenchuan) earthquake | Case series | Hypertension | May 2008 | 11 patients | Mean blood pressure and heart rate increased immediately after the earthquake, regardless of gender or pre-existing hypertension. BP gradually declined within 6 hours after the earthquake and increased again during aftershocks. |
| Hung<br>(2013)(26) | China | Patients treated by Hong Kong Red Cross three weeks after the 2008 Sichuan earthquake | Cross sectional chart review | Multiple NCDs including hypertension, stroke | June 2008 | 2,034 patient encounters | There was a high prevalence of chronic disease after the earthquake, especially hypertension |
| Huynh<br>(2004)(53) | Vietnam | Women hospitalized with invasive cervical squamous cell carcinoma (subjects) and other extrauterine cancers (controls) | Case control | Cancer | June 1996 – Sept 1996 | 145 women in S. Vietnam, 80 women in N Vietnam | The development of invasive cervical cancer was significantly associated with military service by husbands during the 2<sup>nd</sup> Indochinese War and with parity status. Geographic and temporal variation in cervical cancer rates among Vietnamese women was associated with the movement of soldiers. |
| Li<br>(2010)(118) | China | Rural adults born between 1954 and 1964 in selected communities from the 2002 China National Nutrition and Health Survey | Cross-sectional | Diabetes | 2002 | 7,874 adults | Fetal exposure to severe famine increases the risk of hyperglycemia in childhood. The association is exacerbated by a nutritionally rich environment later in life |
| Li<br>(2011)(109) | China | Rural adults born between 1954 and 1964 in selected communities from the 2002 China National Nutrition and Health Survey | Cross-sectional | Diabetes | 2002 | 7,874 adults | Fetal or infant exposure to famine increases the risk of metabolic syndrome in adulthood. The association is exacerbated by a western dietary pattern or being overweight in adulthood |
| Li<br>(2012)(117) | China | People exposed to the 1959-1961 Chinese famine and those not exposed | Retrospective cohort | Stomach cancer | 1970-2009 | Population level data: (2.4million) | Prolonged malnutrition during early life may increase the risk of stomach cancer later in life. |
| Marom<br>(2014)(45) | Philippines | People who survived the 2013 earthquake and typhoon (Haiyan) in the city of Bogo (northern Cebu island) | Case series | Head and neck tumors | 2013 | 1,844 adults treated at the field hospital – 5% (85) had H&N tumors | Surgical interventions in pts with H&N tumors in a relief mission can be performed for therapeutic, palliative and diagnostic purposes. |
| Read<br>(2015)(46) | Philippines | Patients treated by an Australian govt deployed surgical team in a field hosp in Tacloban for 4 wks after Typhoon Haiyan | Cross sectional | Diabetes | Nov 2013 | 131 people | Sepsis from foot injuries in diabetic patients constituted an unexpected majority of the workload of a foreign collaborative surgical medical team in Tacloban in the aftermath of Typhoon Haiyan |
| Sun<br>(2013)(27) | China | Survivors of Wenchuan earthquake staying in a temporary shelter for more than 1 year | Cross sectional | Hypertension | March-May 2009 | 3,230 adults | Age, family history of hypertension, sleep quality, waist-to-hip ratio, BMI and blood glucose levels are risk factors for earthquake-induced hypertension. Mental stress was not a risk factor. |
| Wagner<br>(2016)(123) | Cambodia | Community health care workers in Siam Reap province | Cross sectional | Type 2 Diabetes | .. | 185 | Community health workers were able to effectively learn diabetes prevention curriculum suggesting that they would be effective at disseminating the information |
| Wu<br>(2015)(96) | China | Senior over age 60 living in 8 villages in China during a 2011 flood | Cross sectional survey | Multiple NCDs including hypertension, diabetes | February 2012 | 1,183 elderly patients | There was a marked decline in health status of the elderly after a flood. There were greater detrimental impacts on women and single elderly |

**Table 10:**
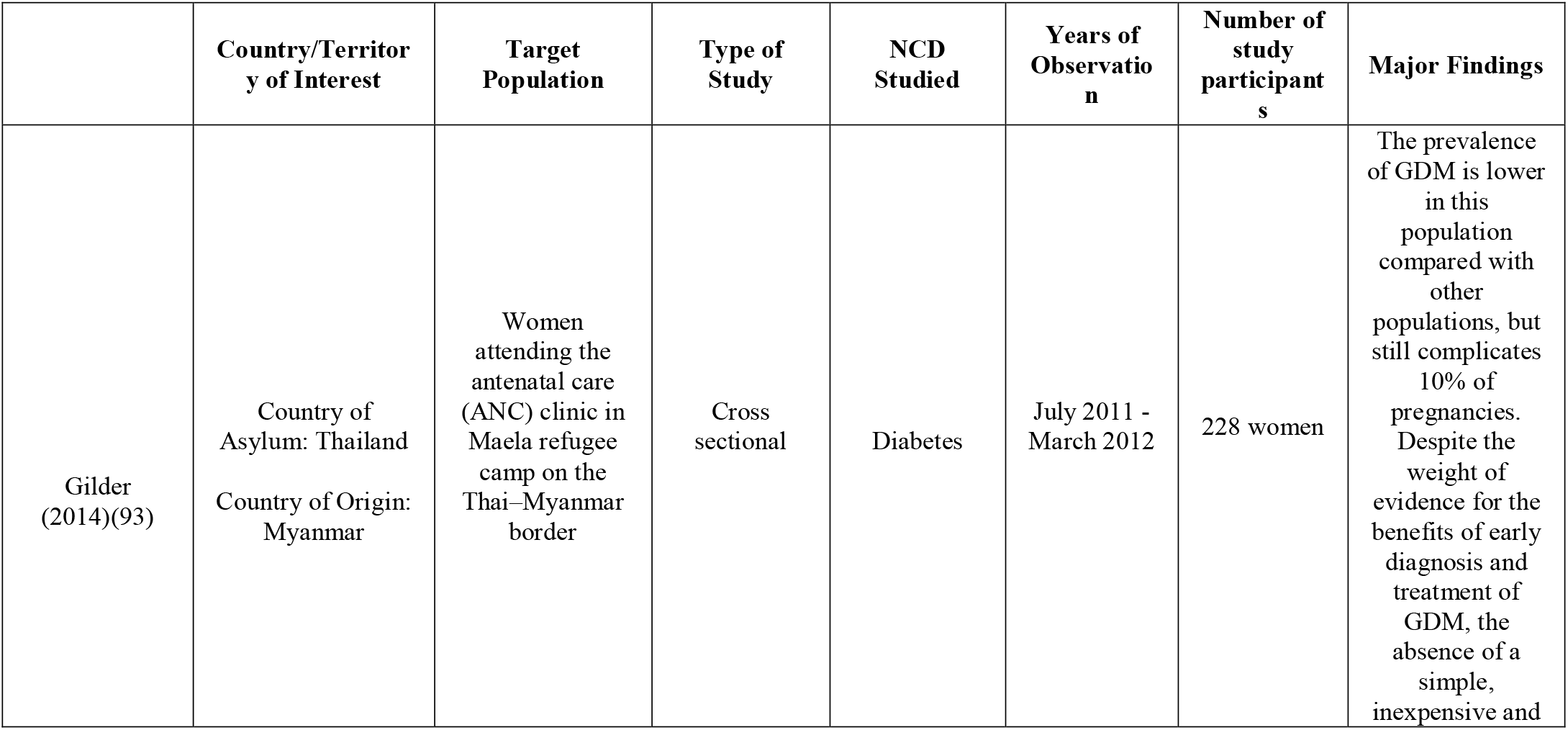
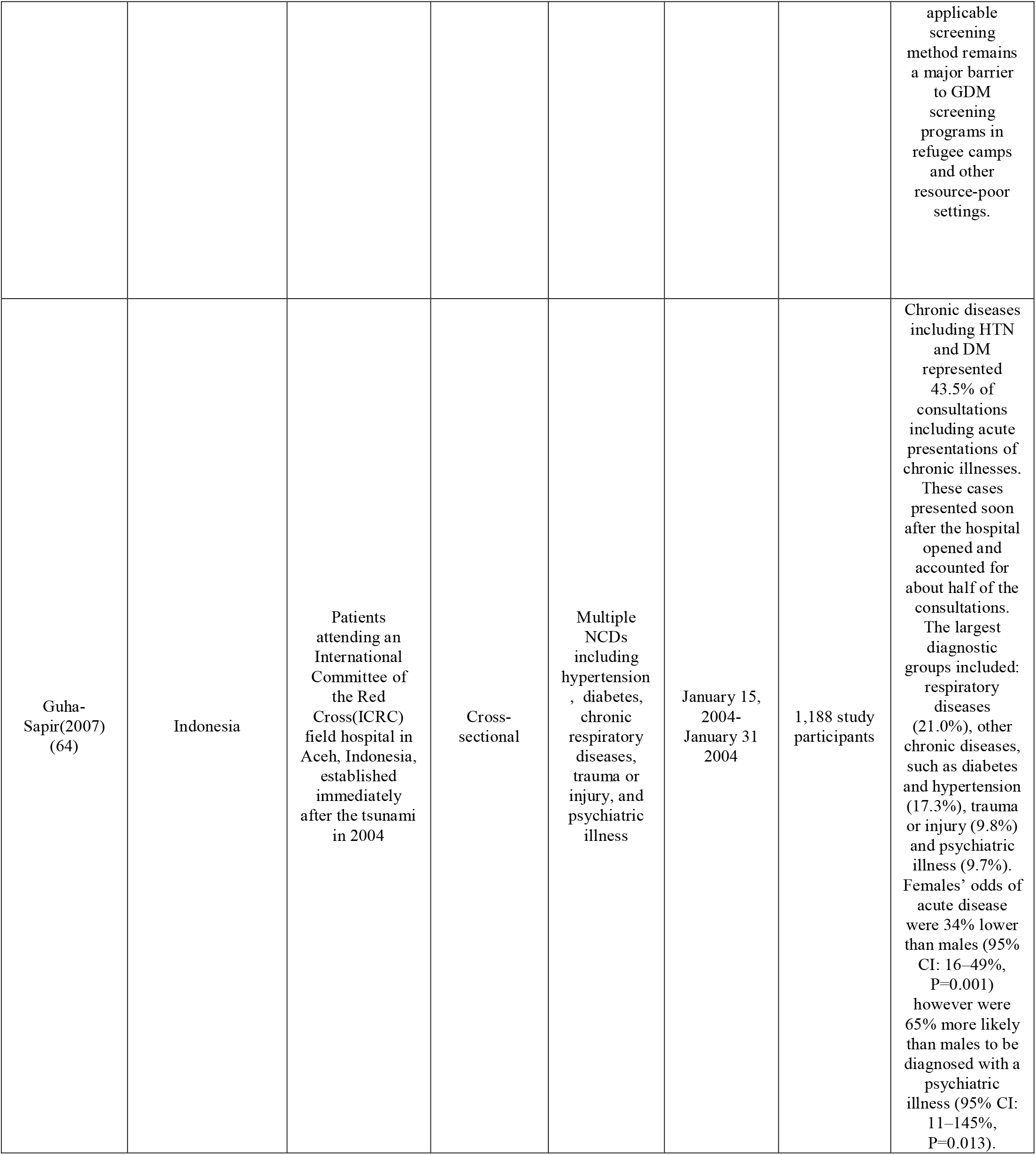
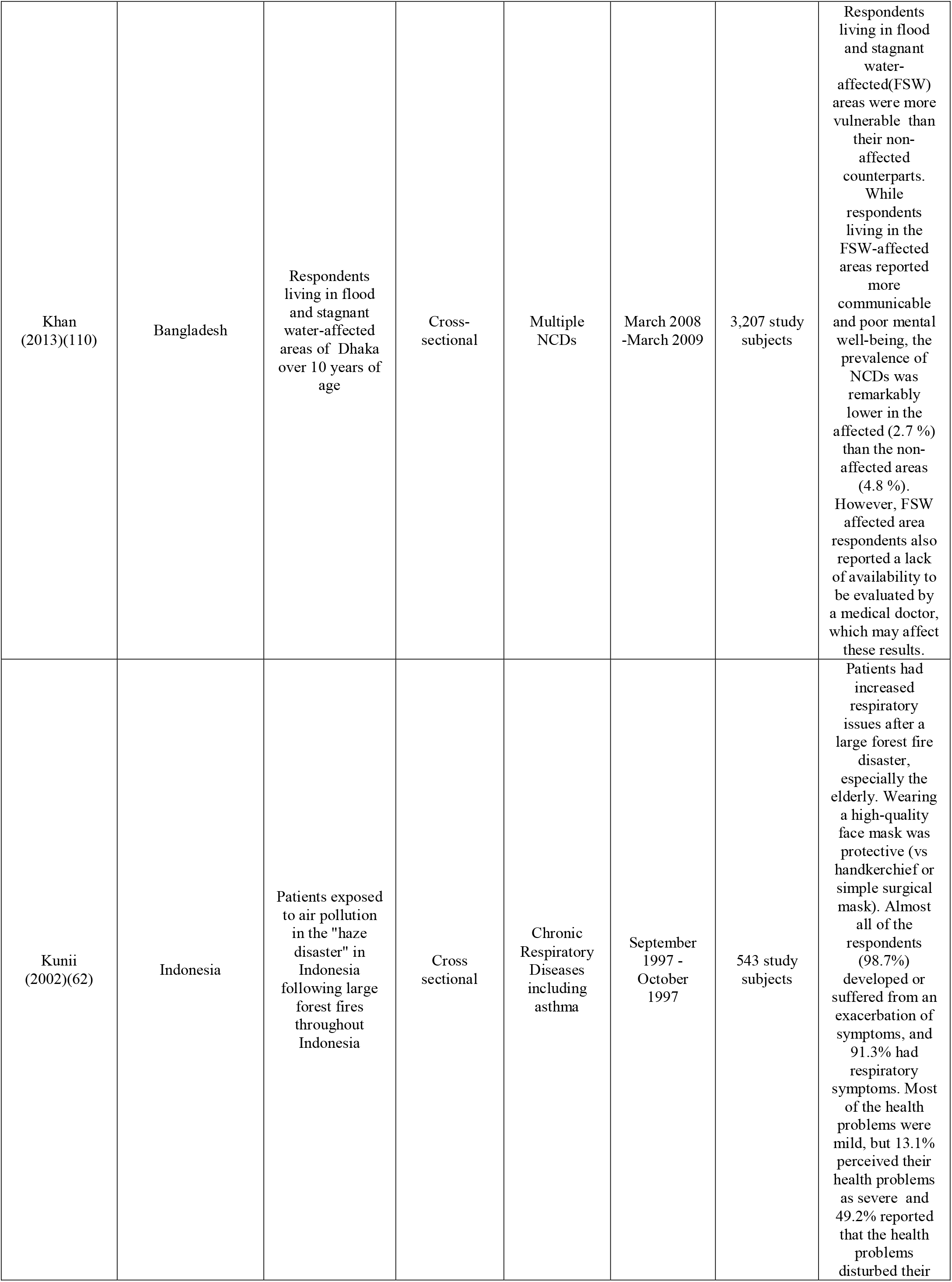
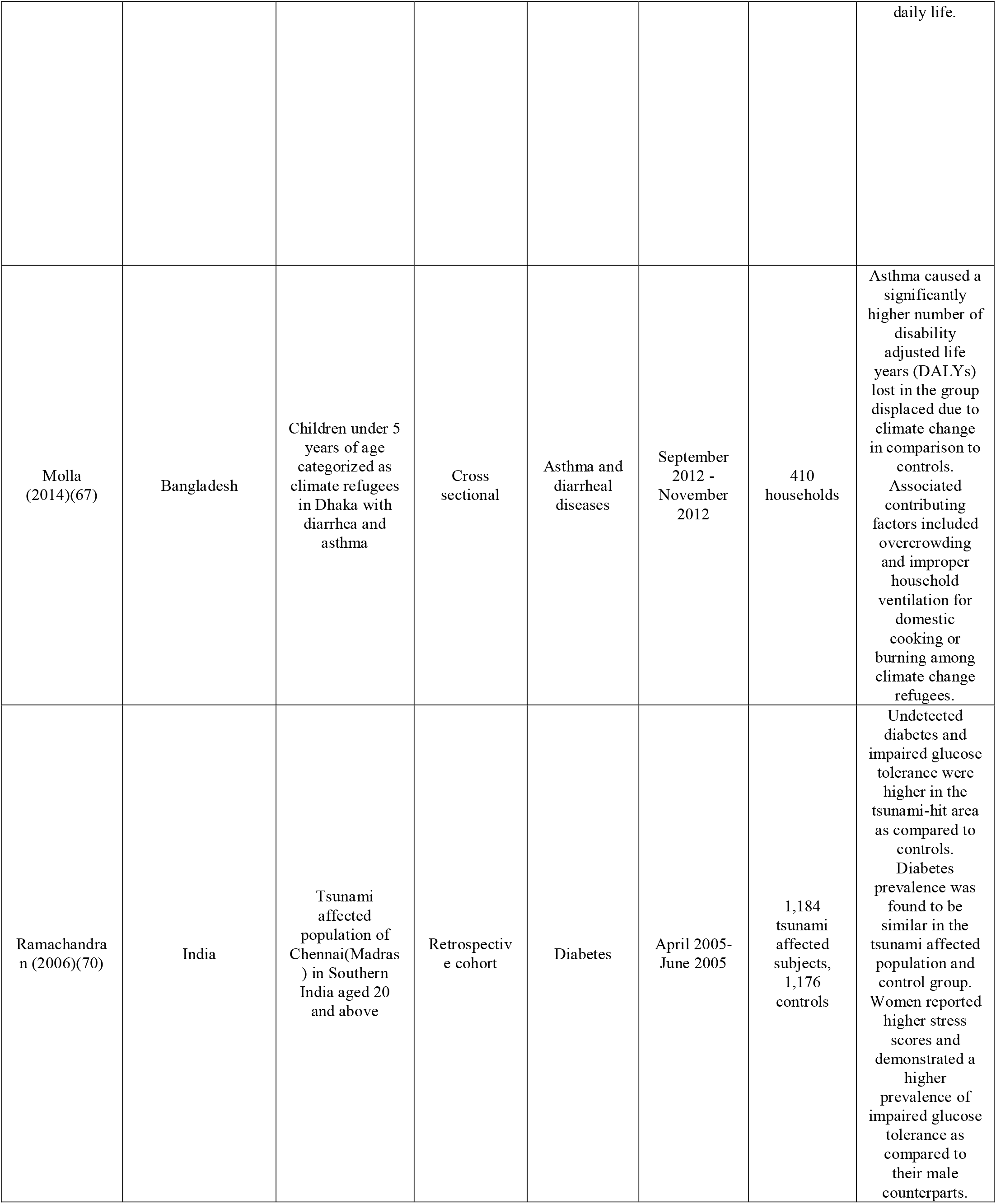
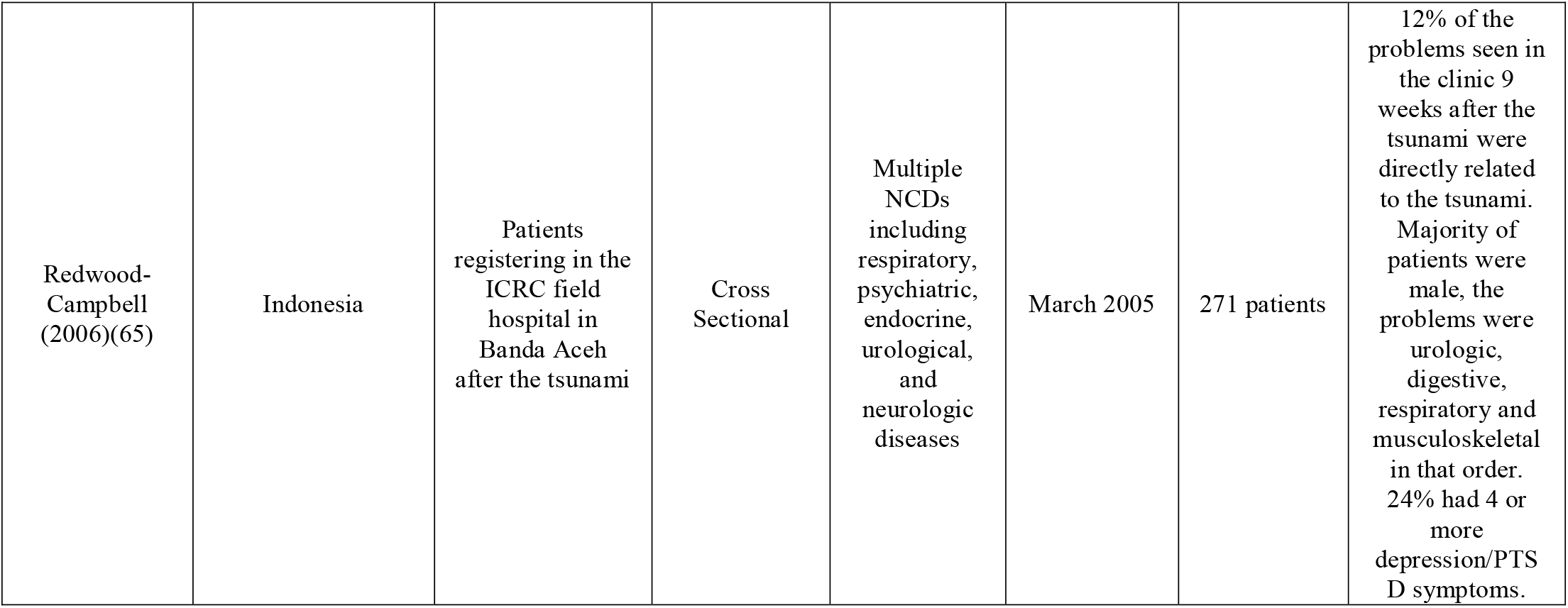
Characteristics of included publications by region: Southeast Asia

In Asia, the Western Pacific, and the Americas there was a specific focus on natural disasters (63, 64), whereas the Eastern Mediterranean Region (EMRO) and Africa regions focused primarily on armed conflict (39, 44, 94, 95). Of note, most studies either reported on the consequences of conflict after the fact (95) or when the population of interest had relocated to a refugee camp or host country (42, 48, 55, 81).

Certain regions or populations were more heavily studied than others as was the case for DM where 57.6% of studies reviewed were conducted in the EMRO region. 32.4% of DM studies focused on the Palestinian (22, 29, 32, 71, 78-82, 84, 105) population alone, higher than Africa, the Americas, Western Pacific (WP), and South East Asia (SEA) combined. The high prevalence of articles conducted in the EMRO may reflect the higher prevalence of diabetes there (106). However, with a rise of diabetes in all regions including Sub-Saharan Africa, where the largest percentage increase in the incidence of diabetes is projected in the coming decade, this represents a significant gap in the available literature (107).

Increased research is needed on NCDs in multiple settings, particularly outside of the EMRO region, and better understanding of the effects of diverse crises, rather than just armed conflict is also key. The effects of climate change (11), and subsequent increasing natural disasters, highlight our need to identify NCD burden in order to guide appropriate responses during these events.

### Methodology

The predominant study design consisted of retrospective chart reviews with a minority of cross-sectional studies (42, 49, 52, 55). We observed that several studies either did not include a comparison group in their study design, or they used a time period across which the comparison was made that was arbitrary in nature. While challenging to conduct given the context of the studies, this limits validity of findings in many studies (42, 45, 48, 53, 55). In addition, it was noted that publications were clustered by research group or author (31, 41, 73, 78-81, 108, 109), which speaks to the need for increased academic outputs in LMICs, and Africa in particular. Finally, several articles that were included in our results included NCDs as a peripheral focus, rather than as primary outcomes (65, 110). There is a need to prioritize NCDs in humanitarian crises, assessing both epidemiology, but also effective interventions to target disease. Consideration should be given to include comparison or control groups in study design, for example individuals in neighboring regions, non-refugee counterparts, or matched sample populations not afflicted by the disease (34, 53) to be able to better assess and thus target the effects of the humanitarian crises itself on disease outcomes. Additionally, long-term cohorts and registries (105) would be ideal to better understand the diversity of diseases and contributory factors in even greater depth. Of all the studies included, none referred to the Sphere guidelines(111), WHO Noncommunicable Diseases in Emergencies brief (112), or WHO PEN package of essential NCD interventions(113) as markers for study design, which we propose be included in future research.

### Concomitant affliction with NCDs and NCD risk factors

We found that the populations studied were commonly afflicted with multiple NCD risk factors (21, 29, 32) and multiple NCDs (21, 22, 29, 41), which supports the need for consolidated care for NCDs as co-affliction confers higher risk of complications (22). Many commonly cited risk factors in HICs such as age, family history, higher BMI, comorbid hypertension, smoking, hyperlipidemia, family history, sedentary lifestyle for DM, cancer, cardiovascular disease were cited (21, 22, 29, 32, 49, 69, 83, 84). However, a lack of association between NCDs and family history as well as other traditional risk factors was also found, and this may result in under-recognition and subsequent under-diagnosis in these settings (93, 110). Greater attention to screening and allocation of resources are needed in disaster-prone settings to prepare, in addition to medical relief efforts deployed during crises specifically prepared to address NCD care.

### Barriers to NCD care

The most commonly cited barriers to healthcare access in all phases of disasters and major disease diagnoses studied, included personal attributes: low levels of education (75, 76, 114), financial difficulties (21, 76, 92), displacement (44, 76), and illiteracy (29, 32, 76). The most commonly cited systems level concerns were lack of access to medications, and affordability of medications (21, 22, 44, 76, 115). Multiple DM specific studies noted that syringes and self-monitoring blood glucose devices were not readily available and posed a financial burden to those who required access to them (44, 75, 77). Several studies also noted shifting of medications from the clinically indicated medication to cheaper or more available options, which may lead to worse outcomes (22, 77). Such challenges may be magnified more for migratory refugees as compared to those who are more established in refugee camps, as demonstrated by Yusef et al (22).

Greater attention to screening and allocation of resources to treat NCDs including acute cardiovascular events such as acute myocardial infarction and stroke are needed in disaster-prone settings, outside of other medical relief efforts. This should include palliative interventions that aim to reduce excess morbidity and suffering from NCDs (116). A health system situational analysis in Tunisia demonstrates the effectiveness of a robustly developing primary health care system, which falls short in this setting without sufficient human resources, reimbursement for public sector, consensus around guidelines for management, and the absence of ancillary providers such as nutritionists or specialists for referral, when needed (86). In post-war Liberia, with majority of CVD deaths occurring within 24 hours of admission, optimization of emergency care which is the first point of contact, was also highlighted (95). Hung et al, the only researchers focusing on the pre-hospital setting, also enforce the importance of raising awareness among first responders of the associated increased burden of NCDs during crisis and propose guidelines adapted to this (26). Finally, decentralization of care to community-based settings, such as for eye care, was presented (30).

### Disaster related stressors and NCD development and morbidity

Multiple studies identified disaster-related psychologic and physical stressors as significant risk factors for NCDs (25, 35, 40, 47, 51, 52, 56, 68-70, 73, 74), as well as described subsequent increased NCD related morbidity as a result of disaster stressors (25, 34, 36, 38, 39, 56, 72, 94). Bereavement, injuries in the family (34, 38, 68), displacement (92), temporal/ geographical proximity (32, 36, 39, 72), and war-related physical and psychological trauma (34, 56, 70) were some of the independent predictors of diagnosis, and increased NCD morbidity (34, 35, 38, 39, 56, 68). Refugee status was independently identified both as a risk factor for diagnosis with an NCD (25, 29, 32, 47, 66), and conferring worse morbidity as indicated by Disability Adjusted Life Years (DALYs) lost (67).

Malnutrition and food insecurity during disaster were commonly cited risk factors for increased NCD morbidity. Notably, fetal exposure to severe famine was associated with an increased risk of cancer (117), DM/impaired glucose tolerance (40, 73-75), metabolic syndrome later in life (109), and the unique phenomenon of Malnutrition Related Diabetes Mellitus (MRDM) (75). The risk of MRDM was exacerbated by a nutritionally rich environment later in life (74, 118). Another hypothesis for the higher prevalence of DM was lack of ability to monitor and control dietary intake and blood sugar during a crisis (21, 29, 47, 72).

Finally, environmental exposures from natural disasters (26, 37, 61-63) and war related toxins (58, 59, 88) contribute to NCD burden for these populations particularly for respiratory and cardiovascular diseases. Natural disasters impacting chronic respiratory illness include thunderstorms, earthquakes, forest fires, volcanic eruptions, and tsunami. Dust storms were a notable exception in one study (37), for which a link to increased pulmonary illness was not shown, while in contrast there was evidence of an effect on cardiovascular disease.

In sum, disaster settings confer higher incidence of NCDs and associated comorbidity Furthermore, attention to refugee status in disaster settings is key given a disparate disease burden. Refugee populations have greater burden of disease and worsened outcomes when compared to host populations. Distribution of disease within a refugee population may be unique, and further divergent by ethnic group even among refugee populations. This is critical information for humanitarian intervention design and implementation.

### Lack of infrastructure for NCDs confers poorer responsiveness during crisis

In many countries affected by humanitarian emergencies, there is scarce data on NCD surveillance, epidemiology, and outcomes in populations at risk in the pre-disaster setting, which creates challenges for disaster mitigation efforts (91, 93). Moreover, poorly functioning systems for delivery of NCD care (44, 85), and underdiagnosis of NCDs (93, 110) in the pre-disaster setting are compounded by new challenges resulting from widespread destruction of the health system (119). Centralized care of NCDs in tertiary health facilities pre-disaster was commonly noted to hinder NCD care access during the relief phase and integration of NCD care into the health system at all levels was supported. (44, 65, 85, 86). Several diseases including leading cancer diagnoses are amenable to prevention, screening, and early detection such as breast cancer (49, 55), cervical cancer (51, 53), and other cancers associated with tobacco use (49, 55). Cervical cancer, for example, is amenable both to primary prevention strategies (HPV immunization and barrier protection during sexual intercourse) as well as secondary prevention (pap smears), and was identified as an opportunity for targeting by several studies with high prevalence including in Vietnam (53) and Croatia (51). Decentralization of primary care provision to community-based settings, such as for eye care, was advocated to address the loss of healthcare infrastructure (65), and may reduce stress on facilities providing emergent care (30, 65). Reinforcement of the public health sector’s capacity for NCD management benefits both the relief phase of disaster response as well as well as post-disaster rehabilitation and reconstruction (119). Overall, increased preparedness (44, 46, 77, 85) and responsiveness by aid providers, health providers, and local governments to NCDs in disasters (44, 46) would help improve disaster mitigation assessments. Validated tools such as the WHO Stepwise approach to Surveillance (120) or Demographic Health Surveys(121) could be used for surveillance or to develop registries in countries.

## Conclusion

An increased focus on the effects of, and mitigating factors for, NCDs occurring in disaster-afflicted LMICs is direly needed. While majority of studies included in our review presented epidemiologic evidence for the burden of disease, research is needed to address contributing factors, and means of managing disease in these extremely resource-variable settings. Regions particularly lacking evidence on LMICs in our study were Africa and the Americas; majority of evidence was from the EMRO region. Among the four lead NCDs, chronic respiratory disease was under-addressed despite evidence that it contributes to high morbidity in crisis. Furthermore, increased evidence on actual diseases such as myocardial infarction and diabetes, rather than simply focusing on risk factors such as hypertension is also needed. Attention to vulnerable populations including women and refugees is also a priority. Refugees have unique exposures that may predispose them to certain illnesses, such as MRDM, and management needs that warrant separate attention from host populations. Given this, we propose that refugee status be considered as an independent risk factor for future studies and interventions. We found only scant interventions designed to address NCDs, which will be presented elsewhere. All in all, screening and prevention for NCDs should be a priority alongside communicable disease programs, such as counseling for smoking cessation, counseling on diet, HPV vaccination, and screening for common cancers like breast and cervical cancer. Studies on implementation for these and other interventions will be key. Additionally, equipping health systems to address NCDs both pre-disaster and during the crisis will enhance these efforts, as well as decentralization of care from tertiary settings that are already overextended during crisis. Finally, the need to address disease in collaboration with other sectors such as agriculture, and urban policy-makers, rather than working in silos, was also supported.

## Supporting information

Appendix: Full Pubmed Search Strategy

## Data Availability

All data generated or analyzed during this study are included in this published article and its supplementary information files. The study is registered at PROSPERO (CRD42018088769).

## Declarations

### Ethics approval and consent to participate

Not applicable.

### Consent for publication

Not applicable.

### Authors’ contributions

CN conceptualized the study, developed the study protocol, led the screening and data extraction processes, and was a major contributor in writing the manuscript. RB conceptualized the study, carried out the literature search and data extraction, and contributed to writing the manuscript. RL conceptualized the study, carried out the data extraction, and was a major contributor in writing the manuscript. DH developed the study protocol and the literature search process, and contributed to writing the manuscript. LW assisted with screening and data extraction. PA and AS contributed to data extraction and contributed to writing the manuscript. AH contributed to study conceptualization, oversaw study protocol development, and contributed to writing the manuscript. All authors read and approved the final manuscript.

## Acknowledgements

Not applicable.

## Authors’ information

CN is an Assistant Professor in the Section of Global Health and International Emergency Medicine in the Department of Emergency Medicine at Yale University. Her research centers on: Non-communicable Diseases (NCDs), barriers to care, and intervention development with a particular focus on East Africa. Her past work includes developing a health linkage to care program for refugees in the lead resettlement state per capita in the US (NE), and serving on the board of the leading refugee resettlement agency in CT. RB holds an MPH in Chronic Disease Epidemiology and has conducted a variety of research projects understanding health impacts of humanitarian emergencies. She is currently a PHI/CDC Surveillance Fellow working at the CDC Zambia country office. RL is a medical student at Ben Gurion University in Beer Sheva, Israel and affiliate researcher in the Department of Emergency Medicine at Yale University, with over seven years of fieldwork with organizations supporting refugee health provision and human rights work, including but not limited to Physicians for Human Rights-Israel, Save a Child’s Heart, the Physicians for Human Rights Student Advisory Board (PHR SAB), and the Integrated Refugee and Immigrant Services(IRIS). PA is the director of Global Health Education in the Department of Emergency Medicine at Yale University, on the Research Committee for the Society for North American Refugee Health Providers, and the President of the Academy for Women in Academic Emergency Medicine, with a specific research focus on refugees and other displaced populations. AS is an Assistant Professor Adjunct of Emergency Medicine at Yale University School of Medicine, having completed the Global Health and International Emergency Medicine Fellowship at Yale University and a Master’s degree in Tropical Medicine & International Health at the London School of Hygiene and Tropical Medicine, and also currently serves as senior editor for the Global Emergency Medicine Literature Review (GEMLR) group. DH has been a medical librarian for 17 years and has experience developing robust and reproduceable search strategies for systematic reviews. AH is an Assistant Professor in the divisions of Education and Global Health in the Department of Emergency Medicine at Brown University and is fellowship trained in disaster medicine and emergency management. She served as co-founder and director for the Uganda Village Project in rural eastern Uganda for 10 years, overseeing public health programs including malaria prevention, family planning, water, sanitation and hygiene.

## List of Abbreviations

AMI: Acute Myocardial Infarction
BMI: Body Mass Index
CAT: COPD Assessment Tool
COPD: Chronic Obstructive Pulmonary Disease CVD: Cardiovascular Disease
DALYs: Disability Adjusted Life Years
DM: Diabetes Mellitus
ECC: Exceptional Care Committee
EMRO: Eastern Mediterranean Region
HICs: High Income Countries
HPV: Human Papillomavirus
HRQOL: Health-Related Quality of Life
HTN: Hypertension
ICRC: International Committee of the Red Cross
IFG: Impaired Fasting Glucose
LDL: Low Density Lipoprotein
LMICs: Low and Middle-Income Countries
NCDs: Non-Communicable Diseases
MRDM: Malnutrition Related Diabetes Mellitus
MeSH: Medical Subject Heading
NGOs: Non-Governmental Organisations
OR: Odds Ratio
SEA: South East Asia
UNHCR: United Nations High Commissioner for Refugees
UNISDR: United Nations Office for Disaster Risk Reduction
UNRWA: United Nations Relief and Works Agency
WHO: World Health Organization
WHOQOL-BREF: World Health Organization Quality of Life Questionnaire
WP: Western Pacific

---

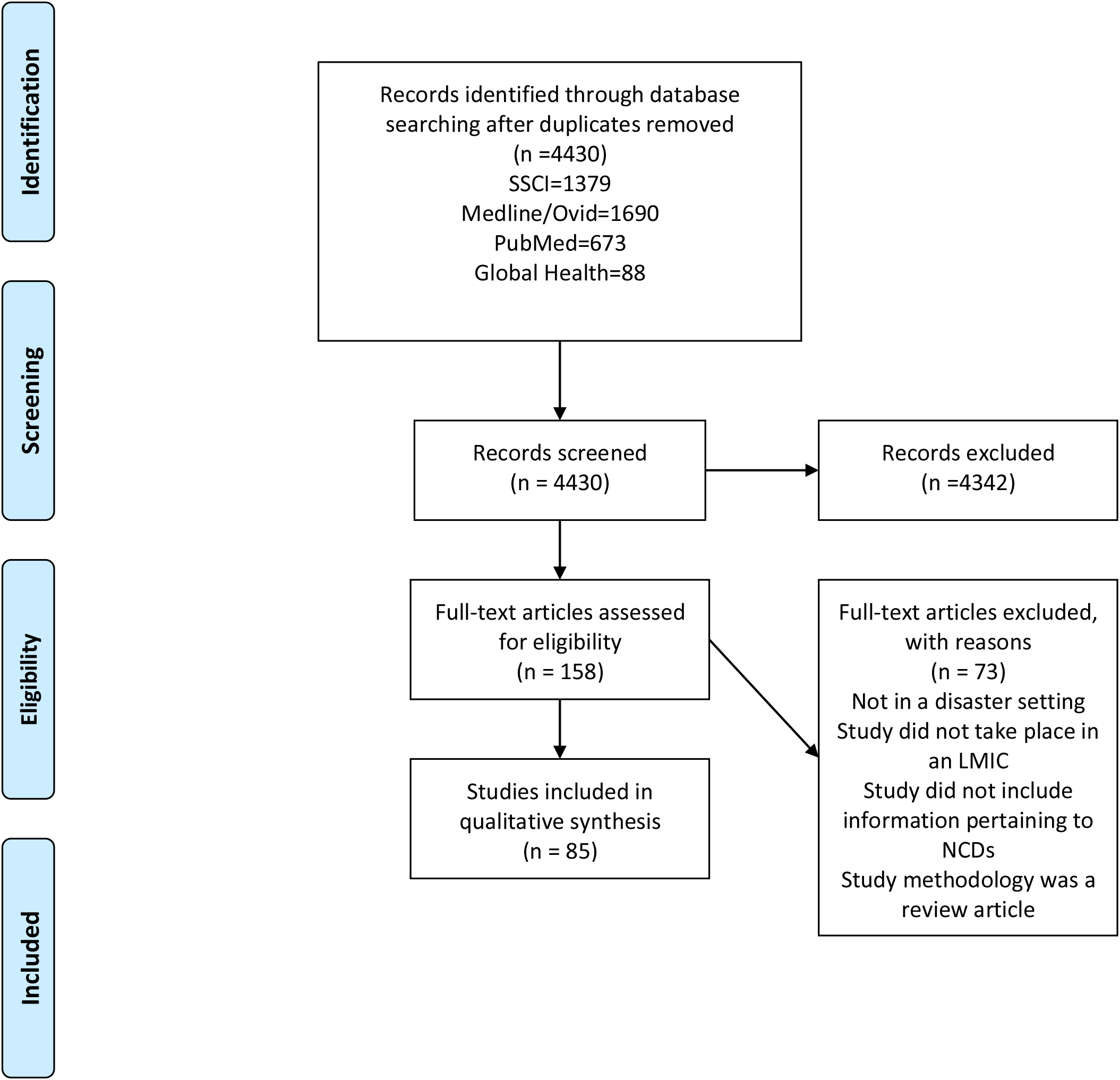
PRISMA 2009 Flow Diagram. From: Moher D, Liberati A, Tetzlaff J, Altman DG, The PRISMA Group (2009). Preferred Reporting Items for Systematic Reviews and Meta-Analyses: The PRISMA Statement. PLoS Med 6(7): e1000097. doi:10.1371/journal.pmed1000097 For more information, visit www.prisma-statement.org.

